# Penetrance and Pleiotropy of Polygenic Risk Scores for Schizophrenia, Bipolar Disorder, and Depression in the VA Health Care System

**DOI:** 10.1101/2022.02.16.22271088

**Authors:** Tim B. Bigdeli, Georgios Voloudakis, Peter B. Barr, Bryan Gorman, Giulio Genovese, Roseann E. Peterson, David E. Burstein, Vlad I. Velicu, Yuli Li, Rishab Gupta, Manuel Mattheisen, Simone Tomasi, Nallakkandi Rajeevan, Frederick Sayward, Krishnan Radhakrishnan, Sundar Natarajan, Anil K. Malhotra, Yunling Shi, Hongyu Zhao, Thomas R. Kosten, John Concato, Timothy J. O’Leary, Ronald Przygodzki, Theresa Gleason, Saiju Pyarajan, Mary Brophy, Cooperative Studies Program (CSP) #572, Million Veteran Program (MVP), Larry J. Siever, Grant D. Huang, Sumitra Muralidhar, J. Michael Gaziano, Mihaela Aslan, Ayman H. Fanous, Philip D. Harvey, Panos Roussos

## Abstract

**Background:** Serious mental illnesses, including schizophrenia, bipolar disorder and depression are heritable, highly multifactorial disorders and major causes of disability worldwide. Polygenic risk scores (PRS) aggregate variants identified from genome-wide association studies (GWAS) into individual-level estimates of liability, and are a promising tool for clinical risk stratification.

**Methods:** By leveraging the VA’s extensive electronic health record (EHR) and a cohort of 9,378 individuals with confirmed diagnoses of schizophrenia or bipolar I disorder, we validated automated case-control assignments based on ICD-9/10 codes, and benchmarked the performance of current PRS for schizophrenia, bipolar disorder, and major depression in 400,000 Million Veteran Program (MVP) participants. We explored broader relationships between PRS and 1,650 disease categories via phenome-wide association studies (PheWAS). Finally, we applied genomic structural equation modeling (gSEM) to derive novel PRS indexing common and disorder-specific latent genetic factors.

**Findings:** Among 3,953 and 5,425 individuals with diagnoses of schizophrenia or bipolar disorder type I that were confirmed by structured clinical interviews, 95% were correctly identified using ICD-9/10 codes (2 or more). Current PRS were robustly associated with case status in European (p<10^−254^) and African (p<10^−5^) participants and were higher among more frequently hospitalized patients (p<10^−4^). PheWAS confirmed previous associations among higher neuropsychiatric PRS and elevated risk for psychiatric and physical health problems and extended these findings to African Americans.

**Interpretation:** Using diagnoses confirmed by in-person structured clinical interviews and current neuropsychiatric PRS, we demonstrated the validity of an EHR-based phenotyping approach in US veterans, highlighting the potential of PRS for disentangling biological and mediated pleiotropy.

**Funding:** Department of Veterans Affairs Cooperative Studies Program (CSP) #572; Million Veteran Program (MVP-000, MVP-006); Office of Research and Development, Department of Veterans Affairs.

## INTRODUCTION

Serious mental illnesses (SMIs) such as schizophrenia (SCZ), bipolar disorder (BIP), and major depression (DEP) are leading causes of disability and public health expenditure, and affected persons suffer disproportionately from increased morbidity and early mortality. Recent years have seen important advances in our understanding of the complex multifactorial underpinnings of SMIs, with genome-wide association studies (GWAS) yielding robust and replicable associations with specific loci— 270, 64, and 44 for SCZ, BIP, and DEP, respectively ^1–3^. However, small effect sizes at individual variants and extreme polygenicity have thwarted the transformative mechanistic insights needed for development of novel therapeutics and prevention strategies.

Polygenic risk scores (PRS) aggregate genetic effects across the genome, including many variants that do not attain genome-wide significance, and can account for more variance in liability than genome-wide significant findings alone, albeit they are typically less predictive than a positive family history ^4^ or certain rare copy number variants ^5^. Ever-increasing GWAS sample sizes have seen the variance in liability captured by PRS climb steadily— from 3% in the first demonstrative application to SCZ ^6^ to upwards of 10% in recent Psychiatric Genomics Consortium (PGC) analyses.^1^ As applied to non-psychiatric traits, the clinical utility of PRS is emergent,^7^ and potential applications in psychiatry are actively being explored,^8^ including risk stratification and predicting treatment response.

With large biobanks now linking the electronic health records (EHRs) of hundreds of thousands of patients to their individual-level genomic data, there is an opportunity to explore the relationships of PRS (or specific variants) with a wide range of clinical phenotypes, i.e. a genotype-to-phenotype or “reverse genetics” paradigm. Also known as a phenome-wide association study (PheWAS),^9^ this unbiased, “disease-agnostic” approach has the potential to uncover hitherto unrecognized relationships between distinct diagnostic entities, and to aid in disentangling complex pleiotropic effects. A recent application of PheWAS from the PsycheMERGE Consortium analyzed PRS for SCZ in more than 100,000 patients from four large health care systems— Geisinger Health System, Mount Sinai Health System, Partners HealthCare System, and Vanderbilt University Medical Center— and uncovered robust associations with both psychiatric and non-psychiatric diagnoses.^10^

The Veterans Health Administration (VHA) is the largest integrated health care system in the United States, with 171 medical centers and 1 112 outpatient clinics serving over 9 million veterans. Launched in 2010, the Million Veteran Program (MVP) is a landmark endeavor that links genomic laboratory testing, survey-based self-report data, and EHRs spanning decades, with the goal of creating a “mega-biobank” and novel evidence base for precision medicine initiatives. ^11^ Demographically and clinically, the 850,000 enrolled participants reflect the population that utilizes the VHA, with over-representation of older and male individuals, as well as higher rates of multiple, chronic conditions compared to the general population ^12,13^, despite better access to healthcare.

Cooperative Studies Program (CSP) #572 ^14^ is a cohort of ∼9,300 veterans with SCZ or bipolar disorder type I (BIP1) that received detailed in-person assessments of clinical diagnosis, functioning, and symptomatology. Within this companion study to the MVP, we evaluated the sensitivity of ICD-9/10 billing codes for SCZ, BIP, and DEP, applying case-control definitions of varying stringency and breadth of clinical phenotype (e.g. SCZ versus any psychosis). We benchmarked penetrance for current neuropsychiatric PRS in 400,000 ancestrally diverse MVP participants and explored the broad relationships of PRS with physical and mental health conditions via PheWAS. Finally, recognizing the considerable shared genetic basis of these disorders, we applied genomic structural equation modeling (gSEM) to derive common and disorder-specific latent genetic factors for comparative genomic analyses, and explored pleiotropic associations of these latent factors with PheWAS.

## METHODS

### Study Participants

This study was approved by the VA Central Institutional Review Board (IRB), and all patients provided written informed consent. Additional details of study ascertainment and assessment are described elsewhere.^14^

#### CSP #572

Participants were recruited through their clinicians, posted notices at participating VA hospitals, and through word of mouth. A total of 27 VA sites participated in this study (with a steady-state of 25 sites). All patients met lifetime DSM-IV criteria for SCZ or BIP1. Patients with major neurologic illnesses, or medical problems that could interfere with central nervous system function, were excluded. Information from medical charts, patients’ clinicians, or other informants were used, if needed, to confirm diagnoses— with all participants receiving the Structured Clinical Interview for the DSM (SCID).^15^ Diagnosed substance abuse was not an exclusion criterion, given the high level of co-occurrence in the population and concerns about representativeness.

#### MVP

Participants are active users of the VHA healthcare system, and were recruited through invitational mailings or by MVP staff while receiving clinical care. Informed consent and authorization per the Health Insurance Portability and Accountability Act were the only other inclusion criteria. All participants completed a Baseline Survey which includes information on demographic factors, family pedigree, health status, lifestyle habits, military experiences, medical history, family history of specific illnesses and physical features; and many also completed an optional Lifestyle Survey ^11^. At the time of writing, 4,697 individuals (∼50% of CSP #572; ∼0.7% of MVP) were dually enrolled in both CSP #572 and MVP.

CSP #572 and MVP participants were genotyped on the MVP 1.0 Axiom array ^16^. Genetic ancestry of participants was classified using the harmonized ancestry and race/ethnicity (HARE) method ^17^, which combines information on genetic ancestry with self-identified race and ethnicity.

### Electronic health records (EHRs)

We compiled a list of commonly prescribed antipsychotics, mood stabilizers, and antidepressants from the VA national formulary (Supplemental Table 1) and extracted these from prescription records. We selected control participants by leveraging the EHR, after exclusion of individuals with any EHR-recorded history of psychotic or affective disorders or past treatment with neuroleptic, mood-stabilizing, or antidepressant medications, or who self-endorsed a personal or family history of any of these conditions on the MVP core survey.

### PRS Profiling

We constructed PRS from published and current GWAS results (the “training” datasets), testing individual-level scores for association with case-control status in MVP/CSP #572 (the “target” dataset). Variants that met quality control filtering in both the training and target datasets were clumped in the appropriate 1000 Genomes Project (1000G) Phase 3 population (*r*^2^>0.1; 500kb window). For varying *p*-value thresholds in the training dataset, scores were constructed by summing the number of tested alleles weighted by their effect estimates (i.e., the log of the allelic odds ratio). We also compared results based on a recently-developed Bayesian framework that applies continuous shrinkage to test statistics, PRS-CS^18^, using the same LD reference panel (1000G EUR) used in Zheutlin et al.^10^. We tested for case-control differences with logistic regression using age, sex, and the first six ancestry principal components (PCs). To compare results with the civilian cohorts, we employed a similar approach to Zheutlin et al. ^10^. Details of sample-level and variant-level quality control are given in the supplemental methods.

### Genomic SEM

We used Genomic Structural Equation Modeling (gSEM),^19^ a recently developed statistical method, to model the genetic covariance structure underlying SCZ, BIP, and DEP. Briefly, gSEM models the multivariate genetic architecture of complex traits by estimating individual SNP associations on latent constructs, is robust to sample overlap and sample-size imbalance, and does not require individual-level genetic data. In the current analysis, we estimated the SNP associations on a common factor (shared between SCZ, BIP, and DEP), as well as SNP associations specific to each disorder. Our analyses were limited to high quality, common SNPs (minor allele frequency >0.05, imputation information>0.9) available across all three of the input GWAS and the 1000G Phase 3 reference panel, resulting in a final set of 5,929,464 SNPs.

### PheWAS

We employed phenome-wide association studies (PheWAS) to summarize associations between neuropsychiatric PRS and “phecodes” representing groupings of related ICD-9/10 billing codes.^9^ Our primary analyses required cases and controls to have two or more and zero ICD-9/10 codes, respectively; we tested phecodes for association with genomic factors within European ancestry (EA) and African ancestry (AA) individuals, using logistic regression and covarying for age, sex, and six ancestry principal components (PCs). To enable comparisons of our results with published ones, we used scaled PRS (mean = 0; SD = 1); logistic regressions were adjusted for age (age and age^2^), sex, and ancestry PCs.

We performed a series of sensitivity analyses, co-varying for selected diagnoses or treatment with antipsychotics, mood-stabilizers, and antidepressants, or removing individuals with any lifetime diagnosis of psychotic, mood, or substance disorders.

## RESULTS

### Validation of EHR-Derived Phenotypes

Table 1 includes descriptives for CSP #572 and MVP and displays the number of participants meeting various case inclusion criteria based on phecodes and medication treatment. CSP #572 recruited a total of 3,953 and 5,425 patients with SCZ and BIP1 diagnosis, respectively, with diagnoses based on the SCID. Phecodes related to SCZ (295.1, 295.2, and 295.3), BIP (296.1) and DEP (296.2), inpatient admission and drug treatment with antipsychotics, mood stabilizers and antidepressants were derived from EHRs, which are available for both CSP #572 and MVP participants through the VHA medical system. We first sought to evaluate the precision and accuracy of EHR-derived phenotypes to capture “caseness” based on the SCID assessment in the CSP #572 participants. Among patients with SCID-confirmed diagnoses of SCZ or BIP1, more than 95% were correctly assigned on the basis of having two or more relevant Phecodes.

**Table 1.** CSP #572 and MVP participants meeting varying EHR-based criteria for SCZ, BIP, and DEP.

|  |  | Cooperative Studies Program (CSP) #572 |  |  |  |  |  | Million Veteran Program (MVP)* |  |  |
| --- | --- | --- | --- | --- | --- | --- | --- | --- | --- | --- |
|  |  | Schizophrenia |  |  | Bipolar I disorder |  |  |  |  |  |
| No. Participants |  | 3,953 |  |  | 5,425 |  |  | 697,921 |  |  |
| Median age (sd) |  | 64 (10.1) |  |  | 61 (11.5) |  |  | 61 (14.2) |  |  |
| Female (%) |  | 289 (7.3) |  |  | 1,005 (18.5) |  |  | 62,749 (9.0) |  |  |
| Diagnosis | Phecode | No. Participants (%) |  |  |  |  |  |  |  |  |
|  |  | ≥1 ICD9/10 | ≥2 ICD9/10 | Inpatient | ≥1 ICD9/10 | ≥2 ICD9/10 | Inpatient | ≥1 ICD9/10 | ≥2 ICD9/10 | Inpatient |
| Schizophrenia | 295.1 | 3,803 (96.2) | **3,770 (95.4)** | 2,978 (75.3) | 1,821 (33.6) | 1,368 (25.2) | 848 (15.6) | 24,698 (3.5) | 18,023 (2.6) | 11,532 (1.7) |
| *Paranoid* | 295.2 | +8 (0.2) | +11 (0.3) | +9 (0.2) | +132 (2.4) | +99 (1.8) | +60 (1.1) | +4,787 (0.69) | +2,806 (0.4) | +1,388 (0.2) |
| *Psychosis* | 295.3 | +26 (0.66) | +39 (1.0) | +64 (1.6) | +524 (9.7) | +402 (7.4) | +208 (3.8) | +19,905 (2.9) | +11,200 (1.6) | +5,402 (0.8) |
|  |  | 3,837 (97.1) | 3,820 (96.6) | 3,051 (77.2) | 2,477 (45.7) | 1,869 (34.5) | 1,116 (20.6) | 49,390 (7.1) | 32,029 (4.6) | 18,322 (2.6) |
| Bipolar disorder (mania) | 296.1 | 610 (15.4) | 462 (11.7) | 189 (4.8) | 3,381 (62.3) | 3,056 (56.3) | 1,134 (20.9) | 16,819 (2.4) | 10,432 (1.5) | 3,919 (0.6) |
| *Bipolar disorder (any)* | 296.1 | +1,063 (26.9) | +691 (17.5) | +613 (15.5) | +1,858 (34.2) | +2,136 (39.4) | +2,660 (49.0) | +62,394 (8.9) | +43,343 (6.2) | +20,228 (2.9) |
|  |  | 1,673 (42.3) | 1,153 (29.2) | 802 (20.3) | 5,239 (96.6) | **5 192 (95.7)** | 3,794 (69.9) | 79,213 (11.3) | 53,775 (7.7) | 24,147 (3.5) |
| Depression | 296.2 | 2,997 (75.8) | 2,506 (63.4) | 1,529 (38.7) | 4,490 (82.8) | 4,048 (74.6) | 2,139 (39.4) | 332,104 (47.6) | 288,561 (41.3) | 120,713 (17.3) |
| Treatment |  | ≥1 Rx | ≥2 trials |  | ≥1 Rx | ≥2 trials |  | ≥1 Rx | ≥2 trials |  |
| antipsychotics |  | 3,784 (95.7) | 3,413 (86.3) |  | 4,793 (88.4) | 3,816 (70.3) |  | 121,339 (17.4) | 56,276 (8.1) |  |
| mood stabilizers |  | 1,834 (46.4) | 706 (17.9) |  | 4,854 (89.5) | 3,319 (61.2) |  | 102,498 (14.7) | 33,340 (4.8) |  |
| antidepressants |  | 2,928 (74.1) | 1,921 (48.6) |  | 4,529 (83.5) | 3,446 (63.5) |  | 357,795 (51.3) | 218,864 (31.4) |  |

\*Participants dually enrolled in CSP #572 and MVP were excluded.

Nearly a third of confirmed SCZ patients had two or more BIP-related Phecodes; and 25% of confirmed BPI patients had multiple ICD-9/10 codes for SCZ. For patients with ICD-9/10 codes for both disorders, taking the prevailing diagnosis (see methods) resulted in fewer than 3% of SCZ cases being misclassified, compared to 10% for BPI. For most misclassified BPI cases, the prevailing diagnosis was schizoaffective disorder (53% specified as schizoaffective bipolar type). In 7% of confirmed BPI cases, the prevailing diagnosis was schizoaffective disorder, with AA veterans overrepresented in this group (38% versus 24% overall).

Comparing receiver operating characteristic (ROC) curves for predictive models based on the varying criteria displayed in Table 1, we concluded that a minimum of two phecodes for SCZ (295.1) or BIP (296.1) offered the best overall balance of sensitivity versus specificity (Figure 1; Supplemental Table 2).

**Figure 1.**
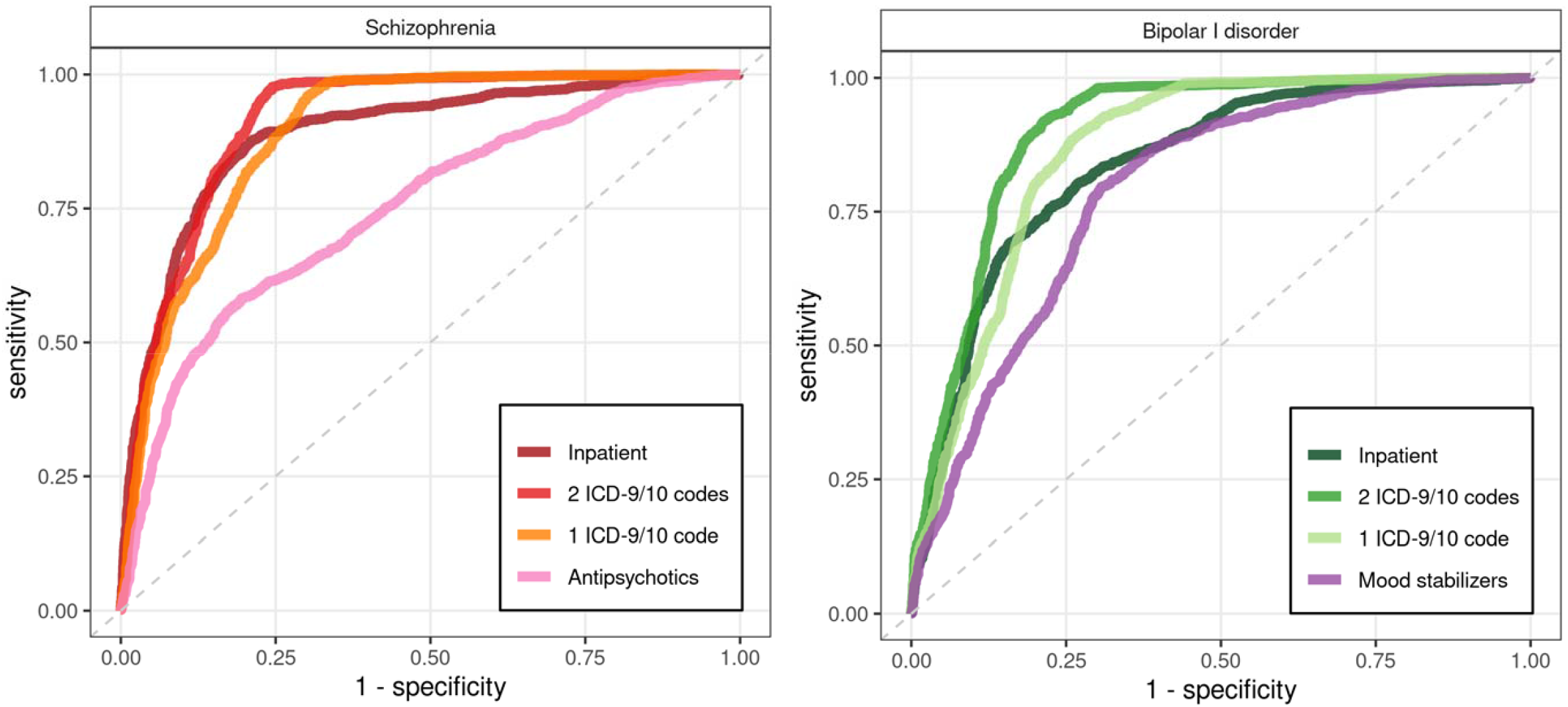
Prediction of SCID-confirmed diagnoses from EHR-based criteria. For varying case criteria displayed in Table 1, sensitivity and specificity estimates for a split-half cross-validation experiment are displayed. The dashed line indicates a random (50/50) prediction.

### Penetrance of neuropsychiatric PRS in the VA Healthcare System

Case prevalence estimates for each decile of PRS are displayed in Figure 2. As expected, the prevalence of SMI was higher among veterans treated at VHA facilities than in the general population ^20,21,22^.

**Figure 2.**
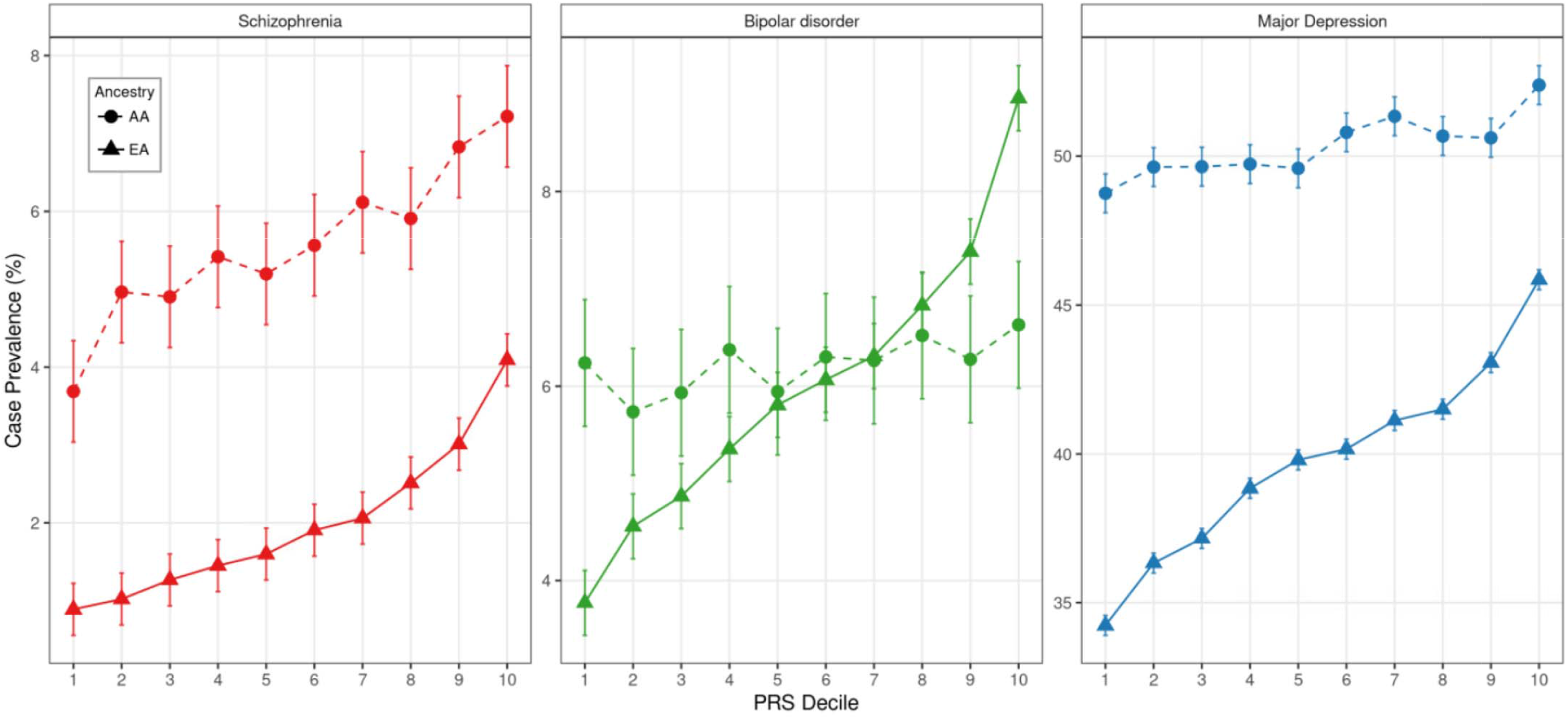
Case prevalence by neuropsychiatric PRS decile in MVP. For PRS constructed from current PGC summary statistics for SCZ, BIP, and DEP, the estimated prevalence of that diagnosis in each PRS decile is displayed separately for AA and EA populations. Ascertained patients enrolled in CSP #572 were excluded.

EA individuals in the uppermost decile of SCZ PRS had ∼2.4-fold higher odds of being diagnosed with SCZ than those below the 90% percentile (95% CI=2.26, 2.55; p<10^−183^); and 4.8-fold higher odds than those in the bottommost decile (95% CI=4.22, 5.43; p<10^−131^). Corresponding estimates for psychosis (Phecode 295.3) were 2-fold (95% CI=1.85, 2.09; p<10^−101^) and 3.2-fold (95% CI=2.89, 3.63; p<10^−90^), respectively; 1.6-fold (95% CI=1.53, 1.66; p<10^−106^) and 2.4-fold (95% CI=2.24, 2.57; p<10^−131^) for BIP; and 1.2-fold (95% CI=1.15, 1.21; p<10^−35^) and 1.4-fold (95% CI=1.33, 1.43; p<10^−70^) for DEP.

Odds of being diagnosed with BIP were 1.7-fold higher for the top decile of BIP PRS compared to the bottom 90% (95% CI=1.61, 1.75; p<10^−126^); and 2.5-fold greater when comparing the top and bottom deciles (95% CI=2.34, 2.71; p<10^−135^). We observed comparable risks of being diagnosed with SCZ or any psychosis, and similarly increased odds of DEP as seen for SCZ PRS.

Comparing individuals in the top decile of DEP PRS with the remaining 90%, odds of DEP were ∼1.4-fold higher (95% CI=1.26, 1.45; p<10^−16^); and 1.7-fold higher for the top versus bottom decile (95% CI=1.55, 1.94; p<10^−21^), with similarly increased odds of SCZ, psychosis, and BIP.

### Cross-ancestry portability of neuropsychiatric PRS

The higher prevalence of SCZ and DEP among AA individuals was largely unrelated to individuals’ risk strata (Figure 2).

AA individuals in the top decile of SCZ PRS had ∼1.4-fold higher risk of diagnoses of SCZ than those below the 90% percentile (95% CI=1.34, 1.57; p<10^−18^) and 2.2-fold higher risk than those in bottom decile (95% CI=1.89, 2.65; p<10^−19^); with comparisons of psychosis risk yielding nearly identical estimates. At extremes, odds of BIP and DEP were increased 1.6-fold (95% CI=1.33, 1.84; p<10^−7^) and 1.3-fold (95% CI=1.17, 1.39; p<10^−7^), respectively.

### Current PRS for SCZ have equivalent penetrance in civilian and veteran health care systems

The demographics of the US veteran population differs from cohorts recruited from civilian health care systems. For instance, the VA population comprises mostly males (∼90%) and has higher prevalence for neuropsychiatric disorders when compared to civilian cohorts ^22^. Using the same training GWAS and Bayesian PRS ^18^ approach as recently employed by the PsycheMERGE consortium^10^, we see a robust association of SCZ PRS with SCZ diagnosis (OR per SD unit increase, 1.56, 95% CI=1.52,1.61; *p*=2.7×10^−222^) in the EA subset of our cohort, which is within the confidence interval (OR = 1.55, 95% CI=1.39,1.72) estimated from four civilian health care systems ^10^.

### Higher loadings of neuropsychiatric PRS in more chronic illness

We observed a trend of increased polygenic loading in more chronic or severe variants of illness ^1,23^. Among EA patients who received inpatient SCZ treatment, we observed a positive association between SCZ PRS and the number of inpatient codes (β=0.06, 95% CI=0.03, 0.09; p=8.65×10^−5^). We observed a similar pattern of results for BIP (β=0.03, 95% CI=0.02, 0.05; p=3.97×10^−4^) and DEP (β=0.02; 95% CI=0.01, 0.02; p=2.95×10^−5^).

Among AA participants, this association was only found for SCZ (β=0.05, 95% CI=0.01, 0.08; p=2.01×10^−5^).

### Pleiotropic influences of neuropsychiatric PRS

Higher polygenic loading for SCZ, BIP, and DEP increased the odds for numerous psychiatric diagnoses and physical health conditions (Figure 3; Supplemental Tables 3-8). Associations between SCZ and respiratory symptoms and infections; and between DEP and cardiovascular disease, hypertension, diabetes mellitus, somatic symptoms, and respiratory problems were robust among veterans without a lifetime diagnosis of psychotic, mood, or substance-use disorders, or who received relevant pharmacological treatment. Comparing PheWAS results, we observed relative enrichments of SCZ PRS for mental health problems, and of DEP PRS for circulatory, respiratory, and endocrine problems, among others.

**Figure 3.**
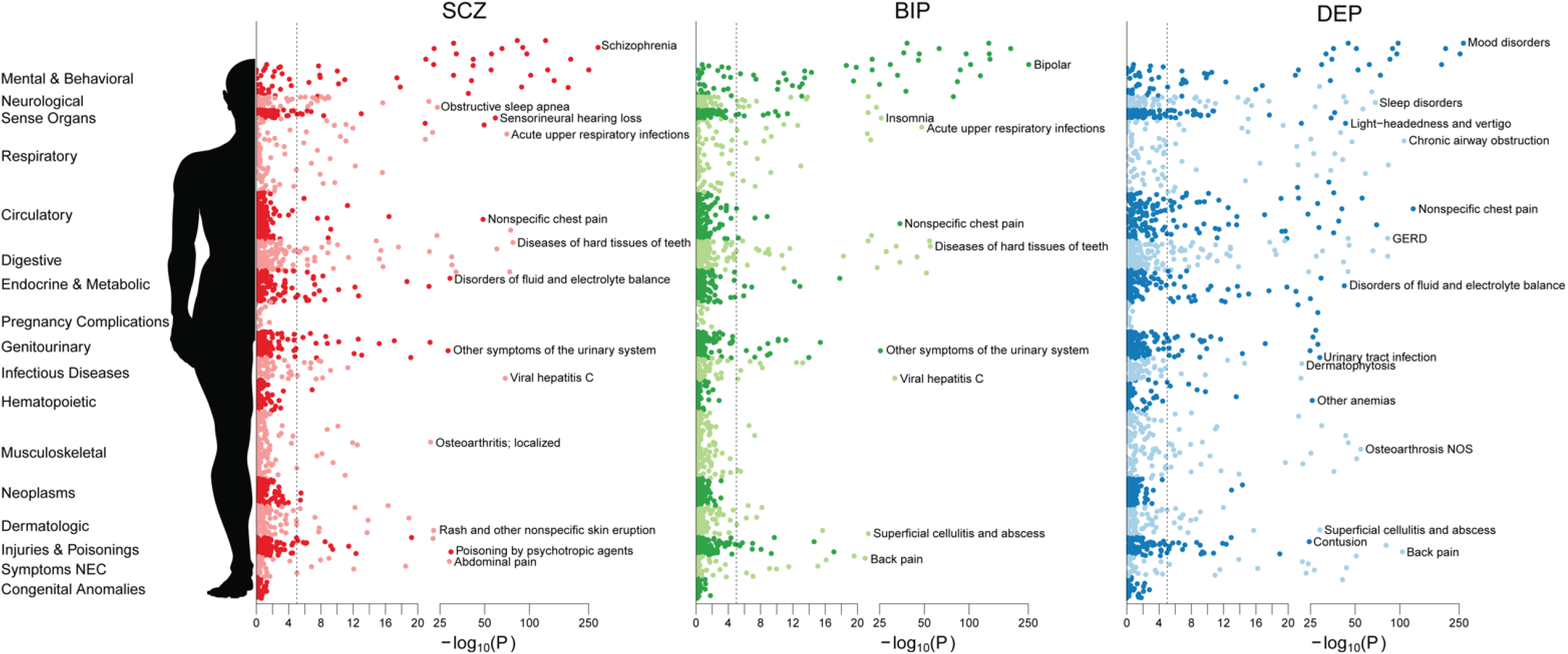
PheWAS results for neuropsychiatric PRS. *(top)* PheWAS of SCZ, BIP, and DEP PRS across 1,650 disease categories are displayed. The dotted line indicates an approximate Bonferroni adjusted *p*-value threshold for the number of Phecodes tested. *(bottom)* For significant results in PheWAS of SCZ and DEP PRS, the distribution of effect sizes within each disease category is displayed as a boxplot.

**Figure 3.**
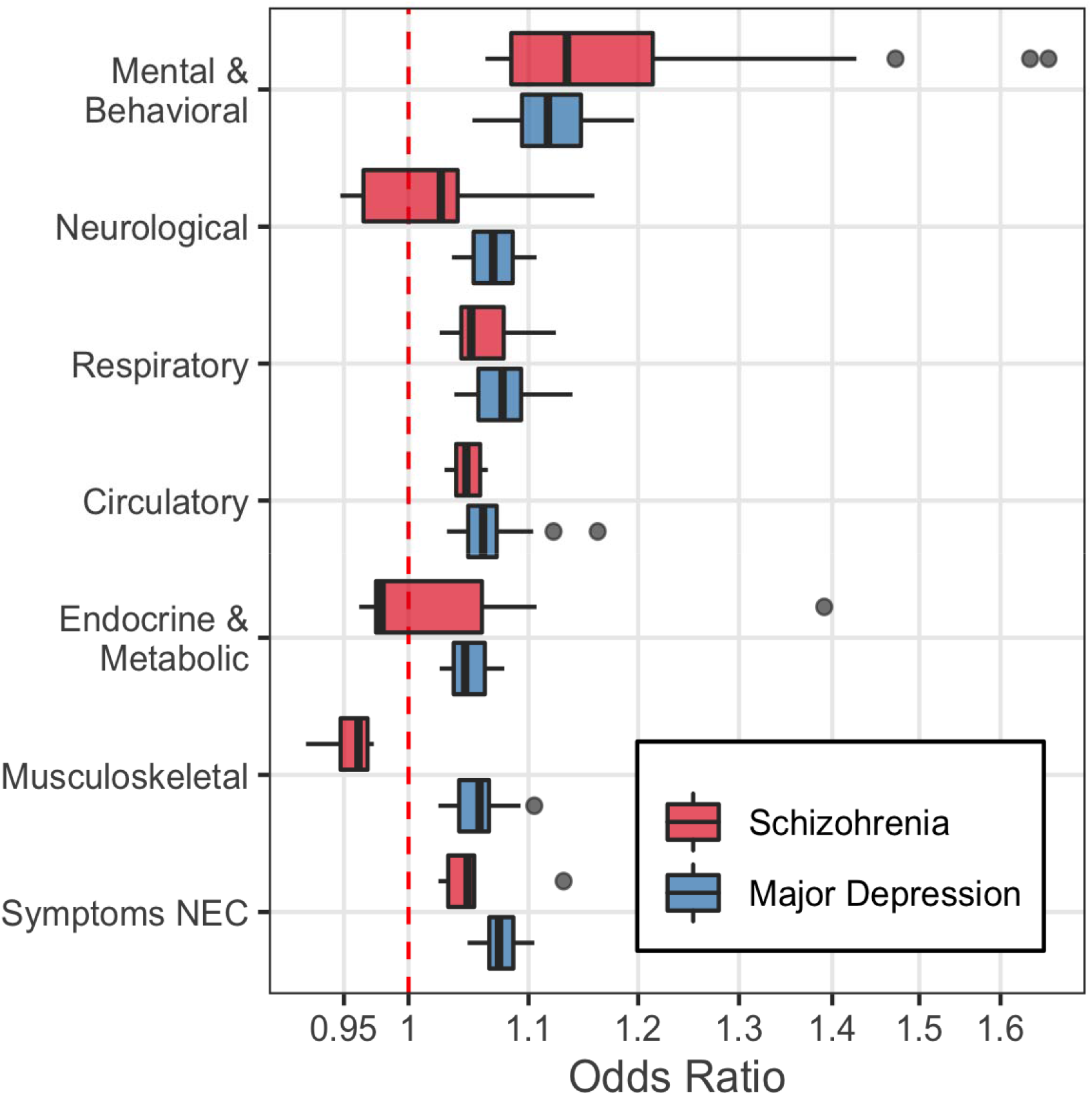
PheWAS results for neuropsychiatric PRS. *(top)* PheWAS of SCZ, BIP, and DEP PRS across 1,650 disease categories are displayed. The dotted line indicates an approximate Bonferroni adjusted *p*-value threshold for the number of Phecodes tested. *(bottom)* For significant results in PheWAS of SCZ and DEP PRS, the distribution of effect sizes within each disease category is displayed as a boxplot.

Relationships between SCZ PRS and dental problems, respiratory symptoms, skin infections, substance use disorders, and suicide were replicable among AA veterans, and remained significant after adjusting for lifetime diagnoses and medications.

### *Genomic SEM* and *latent factor PRS*

Comparing our primary results to those based on latent genomic factors, we found that both SCZ-specific and common-factor liability increased odds of psychosis-spectrum diagnoses.

BIP-specific risk was weakly associated with increased odds of receiving said diagnosis (OR=1.09, 95% CI=1.04,1.14; p=4.64×10^−04^); however, adjusting for treatment with antipsychotics partially restored the associations with BIP (OR=1.16, 95% CI=1.10,1.23; p=3.72×10^−8^).

Observed protective effects of SCZ PRS for sleep apnea, osteoarthritis, and hearing loss appear to be driven by SCZ-specific influences (Supplemental Tables 9-10).

The majority of relationships between SMI PRS and broader psychiatric diagnoses and physical health problems were driven by a shared genetic liability (Supplemental Tables 11-15).

### Polygenic validation of the psychosis-affective spectrum

We further explored the trans-diagnostic spectrum concept via hierarchical assignments of participants to SCZ, BIP, DEP, or related diagnoses; schizoaffective disorders, bipolar II, cyclothymia, and dysthymia were considered as “intermediate” categories of illness and were included in analyses given adequate sample sizes. Figure 4 displays estimated effects of primary and latent factor PRS across disorders, comparing each group to a common set of screened controls.

**Figure 4.**
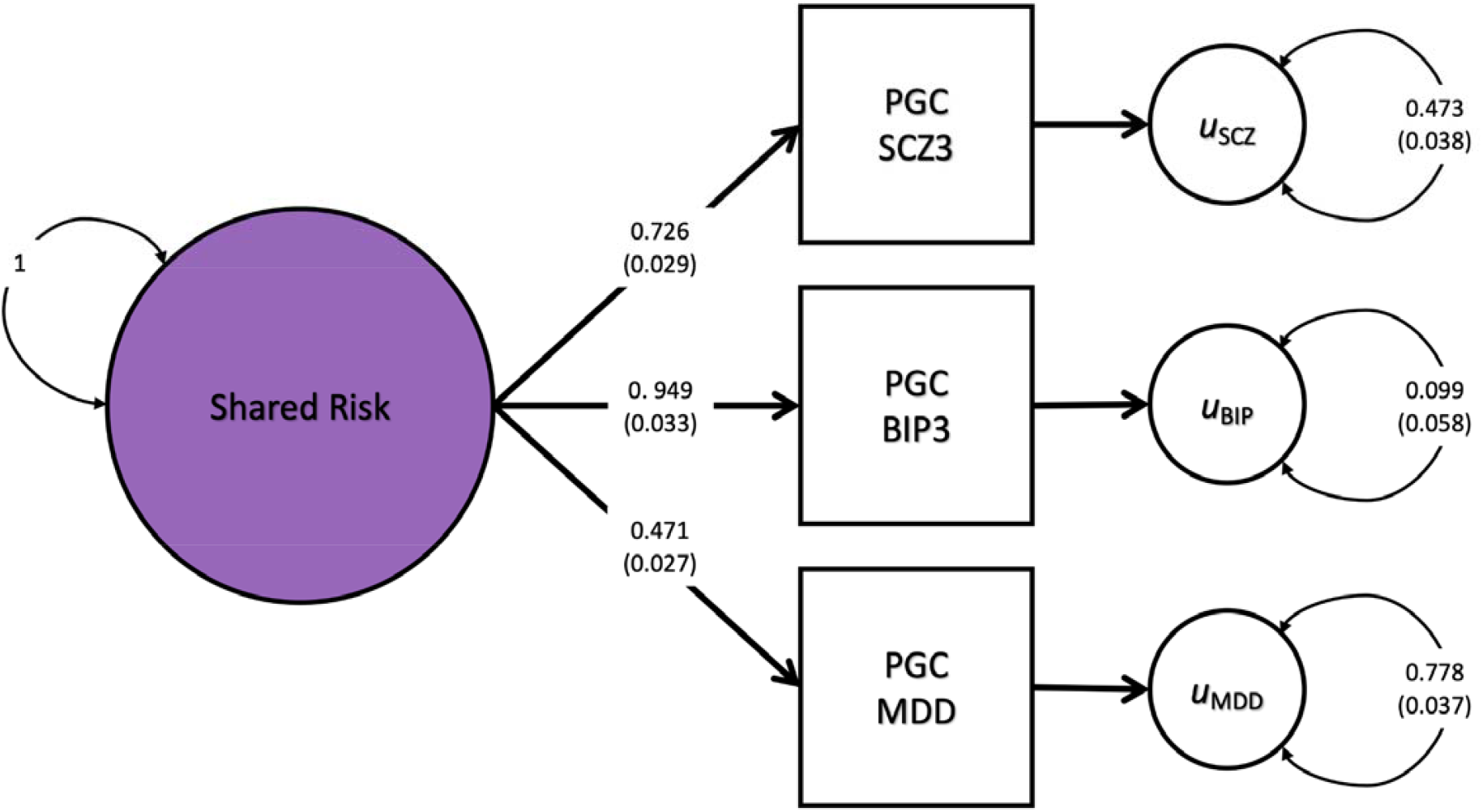
Association of neuropsychiatric PRS with psychotic and affective diagnoses. *(top)* Loadings of a common factor (“p”) on SCZ, BIP, and DEP results, and residual variances corresponding to disorder-specific effects. *(bottom)* Odds ratios (OR) per SD unit increase in PRS for selected diagnoses compared against a common set of controls; analogous results based on latent, disorder-specific PRS appear are plotted in lighter hues. Case assignments were hierarchic al and non-overlapping.

**Figure 4.**
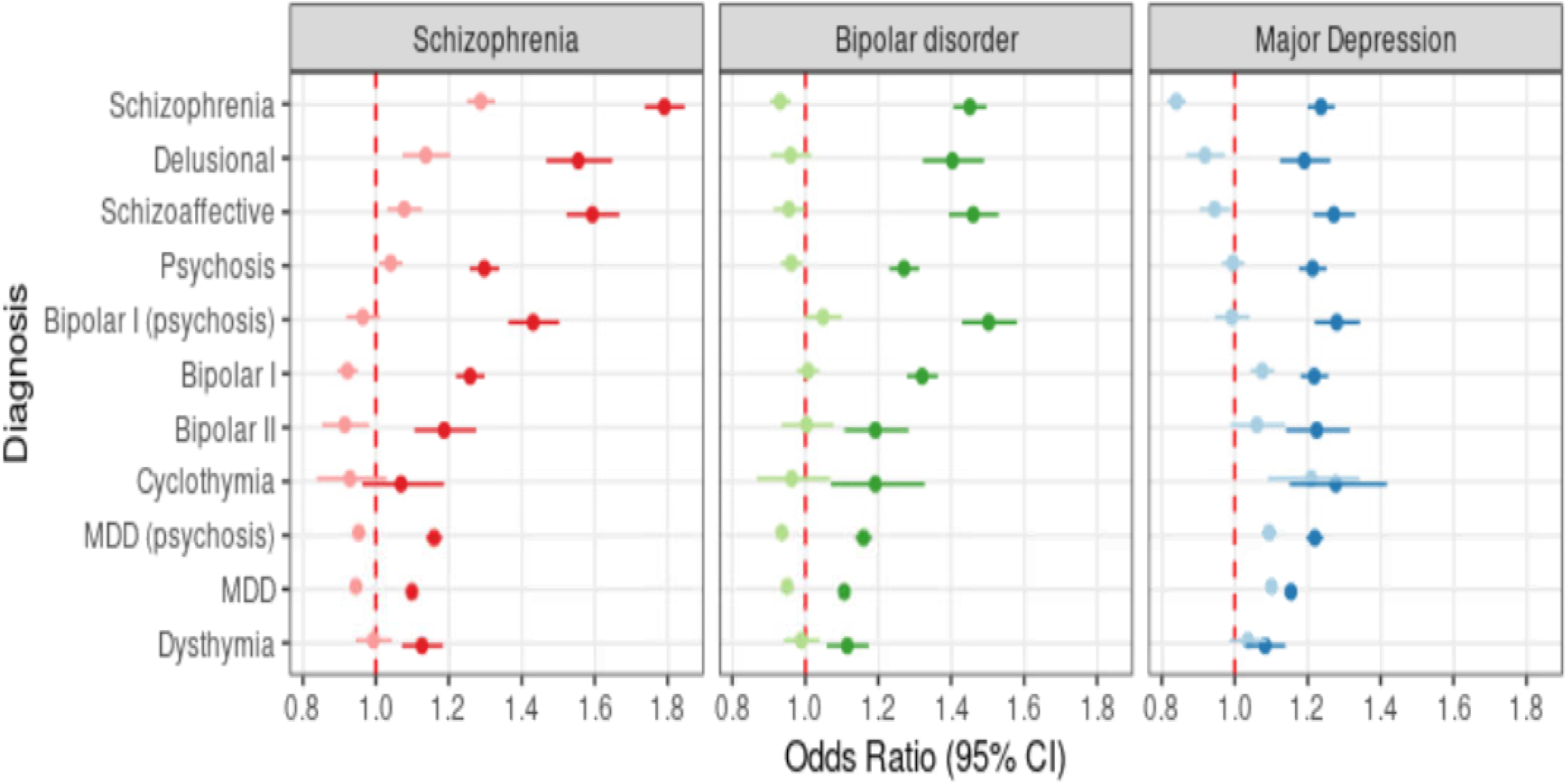
Association of neuropsychiatric PRS with psychotic and affective diagnoses. *(top)* Loadings of a common factor (“p”) on SCZ, BIP, and DEP results, and residual variances corresponding to disorder-specific effects. *(bottom)* Odds ratios (OR) per SD unit increase in PRS for selected diagnoses compared against a common set of controls; analogous results based on latent, disorder-specific PRS appear are plotted in lighter hues. Case assignments were hierarchical and non-overlapping.

## DISCUSSION

Building on our previous reports that published GWAS results are robustly generalizable to the US Veteran population ^24^, we have demonstrated that the penetrance of current neuropsychiatric PRS is equivalent across VA and civilian healthcare systems, despite marked differences in prevalence ^10^. Leveraging the VA’s extensive EHR, we confirm and extend reported associations between neuropsychiatric PRS and broad disease categories in ∼400,000 individuals. We derived novel, latent factors indexing disorder-specific and shared cross-disorder risk and attempted to disentangle widespread pleiotropy from confounding through extensive, secondary modeling.

We first validated an EHR-based phenotyping approach in an embedded, well-characterized cohort of 9,300 patients with confirmed diagnoses of SCZ or BPI and found that a simple approach, ^10^ requiring at least two ICD-9/10 billing codes, correctly identified ∼95% of confirmed cases. Applying this phenotyping strategy to the full MVP cohort, we detected prevalences of SCZ, BIP, and DEP that are several-fold higher than in the general population, albeit representative of the US veteran population at large ^22^. Rates of SCZ and DEP among AA veterans were markedly elevated compared to EA veterans, which may be suggestive of implicit bias in diagnosis ^25^, self-selection bias for VHA utilization, limited alternatives for healthcare, or other structural issues.

Critically, despite markedly higher prevalence of SCZ, BIP, and DEP in the VA healthcare system, we did not find predictive values to be meaningfully attenuated; SCZ PRS yielded effect sizes for EA participants that were virtually identical to those reported by the PsychEMERGE consortium across four US civilian healthcare systems.

Current neuropsychiatric PRS were robustly associated with diagnosed illness, and a range of other psychiatric problems. Higher SCZ PRS also increased risk of suicide, OCD and personality disorders, anxiety, and substance-use behaviors, as well as a host of physical and somatic symptoms, recapitulating recent findings based on four US civilian healthcare systems ^10^. Notably, increased risks of certain infections and dental problems were detectable even when adjusting for psychotic and affective diagnoses and treatment, and excluding diagnosed substance-use disorders, suggesting that neuropsychiatric liability may be penetrant even in individuals who lack a formal diagnosis. Other relationships, such as those observed with erectile dysfunction and polydipsia, were explained by relevant diagnoses and side-effects of prescribed medications ^26^.

We observed protective effects of SCZ PRS on hearing loss, osteoarthritis, obesity and diabetes mellitus, and sleep apnea, in contrast to widely documented, risk-increasing iatrogenic effects of second-generation antipsychotics ^27^. In contrast, DEP PRS were strongly associated with increased risk of obesity and diabetes.

Latent, disorder-specific influences on SCZ were primarily associated with diagnoses of SCZ, paranoid disorders, psychosis, and schizoid personality disorder, but only modestly with BIP, evincing some fidelity of published GWAS to Kraepelin’s dichotomy. In contrast, BIP-specific PRS showed attenuated effects on their BIP risk, but which were recoverable by adjusting for lifetime diagnoses of psychosis-spectrum diagnoses, or treatment with antipsychotics.

Strikingly, the higher prevalence of SCZ and DEP among AA veterans were largely unrelated to individuals’ risk strata; for example, only EA veterans in the uppermost SCZ PRS decile had absolute risk comparable with AA veterans in the lowest decile. Despite lower cross-population generalizability of BIP and DEP PRS, comparisons of AA individuals at extremes of PRS yielded significant associations.

## Limitations

We did not attempt to model environmental or experiential differences related to participants’ military service, which may partially explain increased rates of some illnesses. We did not specifically investigate the implications of predominantly male ascertainment in MVP; we simply adjusted for the effects of biological sex. CSP #572 participants largely served in the period between the Vietnam and Gulf war conflicts, while MVP participants’ service eras were more broadly distributed.

Because available EHR data is restricted to treatment received at VA facilities, any relevant medical history outside the VHA health system, including before or during participants’ military service, is limited to self-report.

## Conclusions

Application of current neuropsychiatric PRS to the MVP yielded results consistent with multiple, continuous liability distributions underlying SCZ, BIP, and DEP ^28,29^, underscoring the advantages of multivariate and trans-diagnostic approaches for studying these complex, heterogeneous clinical presentations.

## Supporting information

Supplemental methods, tables, figures, and acknowledgements

## Data Availability

All summary results produced in the present study are available upon reasonable request to the authors.

## ACKNOWLEDGEMENTS

This research was supported by the Department of Veterans Affairs CSP #572 and the Million Veteran Program (MVP-000 and MVP-006). The Million Veteran Program is supported by the Office of Research and Development, Department of Veterans Affairs. The contents do not represent the views of the U.S. Department of Veterans Affairs or the United States Government. Complete acknowledgements for CSP #572 and MVP are given in the Supplementary Materials.

Dr. Bigdeli is the recipient of a 2019 NARSAD Young Investigator Grant (#28276).

Dr. Voloudakis is supported by the National Institutes of Health (NIH) under award number K08MH122911 and is the recipient of a 2020 NARSAD Young Investigator Grant (#29350).

We are grateful to Drs. R. Karlsson Linnér and T.T. Mallard for sharing their script for plotting PheWAS results, which served as the basis of the figure presented herein. Dr. Harvey has served as a consultant to multiple pharmaceutical companies and device manufacturers on phase 2 or 3 treatment development; this consulting work has been determined to be unrelated to the content of the paper.

No other authors report any relevant conflicts of interest.

## Notes

### Author Declarations

This study was approved by the VA Central Institutional Review Board (IRB), and all patients provided written informed consent.

