## Supplemental methods, tables, figures, and acknowledgements for "Penetrance and Pleiotropy of Polygenic Risk Scores for Schizophrenia, Bipolar Disorder, and Depression in the VA Health Care System"

SUPPLEMENTARY METHODS

*Quality Control*

Samples were required to have call rate >98.5%, heterozygosity which deviates 3 SD or less from the mean, concordance between imputed genetic sex matches self-reported gender. A “greedy” algorithm was used to remove individuals with kinship coefficient ≥0.0884 detailed in Supplementary Methods. Variants with MAF >0.01, call rate >98%, Hardy-Weinberg equilibrium $p$-value $>10^{-10}$, and imputation $R^2$ >0.9 were retained for analysis.

*Relatedness checking*

A greedy algorithm was used for excluding related individuals: related samples and samples with cryptic relationship were removed with a kinship coefficient cut-off of ≥ 0.0884 estimated by the KING v.2.0 software (PMID: 20926424) as previously described (PMID: 31358989, PMID: 31676865). First samples with multiple relationships (≥1) were removed, then individuals were removed from relationship pairs by keeping the individuals with the lowest variant missingness rates to maximize the quality of the remaining samples.

*Genomic SEM*

The factor loadings for the shared factor are presented in Figure 4. Because this model is just-identified, there is no way to determine overall fit. However each of the phenotypes shows strong-to-modest loading on this latent factor (SCZ = 0.72; BIP = 0.94; DEP = 0.47). Genomic SEM corrects for inflation in test statistics due to population stratification by default (multiplying input GWAS standard errors by the square root of their LDSC intercept), so the inflation of the test-statistics in QQ-plots is indicative of polygenic signals in the resulting multivariate GWAS.

Of the genome wide significant SNPs ($p < 5x10^{-8}$), we identified 83 independent lead SNPS (LD $r^2 < .1$) associated with the shared latent factor, 36 associated with SCZ-specific variance, 29 associated with DEP-specific variance, and 11 associated with BIP-specific variance. Of the SNPs associated with the shared latent factor, only 10 SNPs showed evidence of significant heterogeneity, using a liberal threshold of $p < 6.02x10^{-4}$, and the vast majority of SNPs seem robustly associated with the shared latent factor.

SUPPLEMENTARY TABLES

**Supplemental Table 1. Medications queried from VHA prescription records.**

| Antipsychotics | | Mood stabilizers | Antidepressants | |
|---|---|---|---|---|
| aripiprazole | ziprasidone | carbamazepine | Amitriptyline | Sertraline |
| asenapine | olanzapine | divalproex | Buproprion | Trazadone |
| cariprazine | loxapine | lamotrigine | Citalopram | Venlafaxine |
| chlorpromazine | molindone | lithium | Desipramine | Vortioxetine |
| clozapine | thioridazine | valproate | Desvenlafaxine | Doxepin |
| fluphenazine | trifluoperazine | gabapentin | Duloxetine | Mirtazapine |
| haloperidol | brexpiprazole | lacosamide | Escitalopram | Nortriptyline |
| Iloperidone | zuclopentixol | topiramate | Fluoxetine | Tranylcypromine |
| lurasidone | paliperidone | zonisamide | Fluvoxamine | trimipramine |
| perphenazine | sertindole | riluzole | Imipramine | Bupropion |
| pimozide | mesoridazine | oxcarbazepine | Paroxetine | Nefazodone |
| quetiapine | thiothixene | valproic acid | Phenelzine | Selegiline |
| risperidone | zuclopenthixol | | Selegilene | Trazodone |

**Supplemental Table 2. Predictive models for SCZ or BPI diagnosis based on EHR.** Results are displayed for split-half, training/testing experiment in CSP #572. Results for varying thresholds are reported.

| | | Threshold | | | | | | | | |
|---|---|---|---|---|---|---|---|---|---|---|
| | | P(case)>0.5 | | | P(case)>0.7 | | | P(case)>0.9 | | |
| Outcome | Predictor | Sensitivity | Specificity | AUC | Sensitivity | Specificity | AUC | Sensitivity | Specificity | AUC |
| SCZ/BIP | 1 ICD-9/10 | 0.697 | 0.961 | 0.829 | 0.914 | 0.565 | 0.740 | 1.000 | 0.000 | 0.500 |
| | 2 ICD-9/10 | 0.772 | 0.955 | 0.864 | 0.889 | 0.666 | 0.777 | 1.000 | 0.000 | 0.500 |
| | Inpatient code | 0.868 | 0.766 | 0.817 | 0.913 | 0.651 | 0.782 | 0.997 | 0.051 | 0.524 |
| | Antipsychotics | 0.820 | 0.564 | 0.692 | 0.960 | 0.218 | 0.589 | 1.000 | 0.000 | 0.500 |
| BIP/SCZ | 1 ICD-9/10 | 0.989 | 0.562 | 0.775 | 0.990 | 0.560 | 0.775 | 0.991 | 0.549 | 0.770 |
| | 2 ICD-9/10 | 0.981 | 0.699 | 0.840 | 0.981 | 0.699 | 0.840 | 0.983 | 0.674 | 0.828 |
| | Inpatient code | 0.763 | 0.763 | 0.763 | 0.949 | 0.481 | 0.715 | 1.000 | 0.000 | 0.500 |
| | mood-stabilizers | 0.813 | 0.666 | 0.739 | 0.971 | 0.302 | 0.636 | 0.991 | 0.171 | 0.581 |

**Supplemental Table 3. Significant findings ($p$<$10^{-25}$) in PheWAS of SCZ PRS in EA participants.**

| Group | Phecode | Description | Cont | Case | Base | | Diagnosis-adjusted | | Medication-adjusted | |
|---|---|---|---|---|---|---|---|---|---|---|
| | | | | | OR (95% CI) | *p*-value | OR (95% CI) | *p*-value | OR (95% CI) | *p*-value |
| circulatory system | 418 | Nonspecific chest pain | 178521 | 91397 | 1.06 (1.05,1.07) | 6.8E-50 | 1.03 (1.03,1.04) | 5.5E-16 | 1.04 (1.03,1.04) | 3.0E-17 |
| digestive | 521 | Diseases of hard tissues of teeth | 233809 | 60055 | 1.09 (1.08,1.1) | 6.5E-79 | 1.06 (1.05,1.07) | 1.0E-33 | 1.05 (1.04,1.06) | 5.5E-26 |
| digestive | 521.1 | Dental caries | 236323 | 58432 | 1.09 (1.08,1.1) | 3.3E-76 | 1.06 (1.05,1.07) | 2.8E-32 | 1.05 (1.04,1.06) | 9.4E-25 |
| digestive | 523 | Gingival and periodontal diseases | 246246 | 49998 | 1.09 (1.08,1.1) | 1.4E-61 | 1.05 (1.04,1.07) | 1.8E-25 | 1.05 (1.04,1.06) | 8.8E-20 |
| digestive | 523.3 | Periodontitis (acute or chronic) | 271618 | 25654 | 1.08 (1.07,1.1) | 4.2E-33 | 1.05 (1.04,1.06) | 5.3E-13 | 1.04 (1.03,1.06) | 8.8E-10 |
| digestive | 525 | Other diseases of the teeth and supporting structures | 230287 | 60680 | 1.09 (1.08,1.1) | 4.5E-75 | 1.05 (1.04,1.06) | 4.7E-29 | 1.05 (1.04,1.06) | 9.2E-21 |
| digestive | 525.1 | Loss of teeth or edentulism | 261829 | 38073 | 1.07 (1.05,1.08) | 4.2E-31 | 1.03 (1.02,1.04) | 5.1E-08 | 1.02 (1.01,1.03) | 9.8E-05 |
| digestive | 578 | Gastrointestinal hemorrhage | 250313 | 35111 | 1.07 (1.06,1.08) | 5.4E-31 | 1.05 (1.04,1.06) | 8.5E-16 | 1.04 (1.03,1.06) | 1.2E-13 |
| endocrine/metabolic | 276 | Disorders of fluid, electrolyte, and acid-base balance | 221908 | 54151 | 1.06 (1.05,1.07) | 3.3E-30 | 1.03 (1.02,1.04) | 3.9E-10 | 1.02 (1.01,1.03) | 6.7E-05 |
| genitourinary | 599 | Other symptoms/disorders or the urinary system | 215307 | 59968 | 1.05 (1.04,1.06) | 2.4E-29 | 1.03 (1.02,1.04) | 8.2E-11 | 1.03 (1.02,1.04) | 1.1E-11 |
| infectious diseases | 70.3 | Viral hepatitis C | 294978 | 13935 | 1.17 (1.15,1.19) | 4.2E-70 | 1.14 (1.12,1.16) | 1.8E-50 | 1.13 (1.11,1.15) | 1.6E-42 |
| injuries & poisonings | 969 | Poisoning by psychotropic agents | 306220 | 1944 | 1.3 (1.25,1.36) | 1.2E-30 | 1.25 (1.19,1.31) | 6.5E-22 | 1.18 (1.12,1.23) | 3.2E-12 |
| mental disorders | 290.3 | Other persistent mental disorders due to conditions classified elsewhere | 288225 | 14474 | 1.11 (1.09,1.13) | 6.9E-32 | 1.06 (1.04,1.08) | 2.2E-11 | 1.05 (1.03,1.06) | 3.1E-07 |
| mental disorders | 292 | Neurological disorders | 246745 | 40721 | 1.08 (1.06,1.09) | 7.5E-42 | 1.04 (1.03,1.05) | 3.0E-11 | 1.03 (1.02,1.04) | 1.7E-06 |
| mental disorders | 296 | Mood disorders | 161370 | 132442 | 1.12 (1.11,1.13) | 1.3E-184 | 0.87 (0.84,0.91) | 2.8E-11 | 1.08 (1.07,1.08) | 5.5E-66 |
| mental disorders | 296.1 | Bipolar | 285708 | 20399 | 1.29 (1.27,1.31) | 1.0E-253 | 1.24 (1.23,1.26) | 5.5E-171 | 1.2 (1.18,1.22) | 9.7E-99 |
| mental disorders | 296.2 | Depression | 167868 | 124718 | 1.1 (1.09,1.11) | 1.4E-134 | 0.9 (0.88,0.92) | 2.5E-22 | 1.06 (1.05,1.07) | 1.1E-42 |
| mental disorders | 296.22 | Major depressive disorder | 200712 | 92217 | 1.08 (1.08,1.09) | 2.7E-84 | 0.98 (0.97,1) | 5.4E-03 | 1.04 (1.03,1.05) | 7.6E-16 |
| mental disorders | 297 | Suicidal ideation or attempt | 285894 | 14832 | 1.19 (1.17,1.21) | 9.4E-92 | 1.14 (1.12,1.16) | 8.8E-51 | 1.09 (1.07,1.11) | 3.4E-22 |
| mental disorders | 297.1 | Suicidal ideation | 291719 | 12317 | 1.21 (1.19,1.23) | 4.1E-90 | 1.16 (1.14,1.18) | 6.9E-52 | 1.11 (1.08,1.13) | 5.6E-24 |
| mental disorders | 297.2 | Suicide or self-inflicted injury | 304207 | 3948 | 1.24 (1.2,1.28) | 6.4E-40 | 1.19 (1.15,1.23) | 9.1E-26 | 1.12 (1.08,1.16) | 8.4E-12 |
| mental disorders | 300 | Anxiety disorders | 166912 | 123000 | 1.13 (1.12,1.14) | 8.6E-188 | 1.07 (1.06,1.09) | 1.4E-47 | 1.08 (1.07,1.09) | 5.9E-76 |
| mental disorders | 300.1 | Anxiety disorder | 206217 | 79142 | 1.13 (1.12,1.14) | 1.8E-176 | 1.09 (1.08,1.1) | 9.5E-65 | 1.09 (1.08,1.1) | 5.4E-77 |
| mental disorders | 300.11 | Generalized anxiety disorder | 278052 | 21853 | 1.13 (1.12,1.15) | 3.7E-67 | 1.09 (1.07,1.1) | 6.3E-30 | 1.08 (1.07,1.1) | 3.0E-28 |
| mental disorders | 300.12 | Agorophobia, social phobia, and panic disorder | 292966 | 12188 | 1.14 (1.12,1.16) | 6.8E-43 | 1.09 (1.07,1.11) | 4.9E-21 | 1.08 (1.06,1.1) | 4.9E-15 |
| mental disorders | 300.3 | Obsessive-compulsive disorders | 306240 | 3142 | 1.24 (1.2,1.29) | 3.1E-33 | 1.19 (1.15,1.24) | 4.7E-22 | 1.16 (1.12,1.21) | 1.1E-16 |
| mental disorders | 300.4 | Dysthymic disorder | 274401 | 23556 | 1.08 (1.07,1.1) | 3.7E-30 | 1.02 (1.01,1.04) | 5.4E-04 | 1.03 (1.02,1.04) | 3.1E-05 |
| mental disorders | 300.9 | Posttraumatic stress disorder | 230809 | 69537 | 1.07 (1.07,1.08) | 8.7E-57 | 1.02 (1.01,1.03) | 1.6E-06 | 1.03 (1.02,1.04) | 9.0E-08 |
| mental disorders | 301 | Personality disorders | 290676 | 13909 | 1.21 (1.19,1.24) | 1.3E-103 | 1.17 (1.14,1.19) | 1.8E-62 | 1.12 (1.1,1.14) | 1.1E-32 |
| mental disorders | 301.2 | Antisocial/borderline personality disorder | 303440 | 5191 | 1.22 (1.19,1.25) | 5.7E-43 | 1.17 (1.14,1.21) | 1.2E-27 | 1.11 (1.07,1.14) | 1.8E-11 |
| mental disorders | 304 | Adjustment reaction | 238220 | 47151 | 1.06 (1.05,1.07) | 6.8E-32 | 1.02 (1.01,1.03) | 2.6E-03 | 1.03 (1.02,1.04) | 2.0E-09 |
| mental disorders | 306 | Other mental disorder | 257665 | 23904 | 1.11 (1.09,1.12) | 3.2E-51 | 1.07 (1.05,1.08) | 5.8E-20 | 1.06 (1.04,1.07) | 1.4E-15 |
| mental disorders | 316 | Substance addiction and disorders | 261155 | 36576 | 1.17 (1.15,1.18) | 1.1E-148 | 1.12 (1.11,1.14) | 4.2E-81 | 1.1 (1.09,1.12) | 5.8E-55 |
| mental disorders | 317.1 | Alcoholism | 255939 | 39754 | 1.15 (1.13,1.16) | 2.4E-130 | 1.11 (1.1,1.12) | 7.7E-76 | 1.1 (1.09,1.11) | 6.3E-61 |
| mental disorders | 318 | Tobacco use disorder | 175890 | 103781 | 1.07 (1.06,1.07) | 1.9E-56 | 1.04 (1.03,1.05) | 3.1E-25 | 1.04 (1.03,1.05) | 3.3E-21 |
| respiratory | 465 | Acute upper respiratory infections of multiple or unspecified sites | 204993 | 53253 | 1.09 (1.08,1.1) | 8.5E-72 | 1.07 (1.06,1.08) | 9.4E-38 | 1.07 (1.05,1.08) | 5.5E-36 |
| sense organs | 389 | Hearing loss | 121761 | 165684 | 0.94 (0.93,0.95) | 9.6E-51 | 0.93 (0.92,0.94) | 4.2E-73 | 0.94 (0.93,0.94) | 1.0E-62 |
| sense organs | 389.1 | Sensorineural hearing loss | 143787 | 132938 | 0.93 (0.93,0.94) | 3.3E-60 | 0.92 (0.91,0.93) | 1.1E-83 | 0.93 (0.92,0.93) | 1.1E-72 |
| symptoms | 785 | Abdominal pain | 206085 | 63494 | 1.05 (1.04,1.06) | 6.0E-30 | 1.03 (1.02,1.04) | 1.5E-08 | 1.03 (1.02,1.04) | 6.0E-08 |

**Supplemental Table 4. Significant findings ($p<10^{-25}$) in PheWAS of BIP PRS in EA participants.**

| Group | Phecode | Description | Cont | Case | Base | | Diagnosis-adjusted | | Medication-adjusted | |
|---|---|---|---|---|---|---|---|---|---|---|
| | | | | | OR (95% CI) | *p*-value | OR (95% CI) | *p*-value | OR (95% CI) | *p*-value |
| circulatory system | 418 | Nonspecific chest pain | 178521 | 91397 | 1.01 (1.01,1.01) | 2.9E-35 | 1 (1,1.01) | 8.3E-09 | 1.01 (1.01,1.01) | 3.6E-19 |
| digestive | 521 | Diseases of hard tissues of teeth | 233809 | 60055 | 1.01 (1.01,1.02) | 4.7E-56 | 1.01 (1.01,1.01) | 6.8E-20 | 1.01 (1.01,1.01) | 6.2E-30 |
| digestive | 521.1 | Dental caries | 236323 | 58432 | 1.01 (1.01,1.02) | 2.2E-55 | 1.01 (1.01,1.01) | 1.2E-19 | 1.01 (1.01,1.01) | 2.3E-29 |
| digestive | 522 | Diseases of pulp and periapical tissues | 284618 | 14361 | 1.02 (1.02,1.02) | 1.1E-28 | 1.01 (1.01,1.02) | 3.7E-14 | 1.01 (1.01,1.02) | 8.5E-17 |
| digestive | 523 | Gingival and periodontal diseases | 246246 | 49998 | 1.01 (1.01,1.02) | 1.0E-45 | 1.01 (1.01,1.01) | 5.0E-16 | 1.01 (1.01,1.01) | 3.1E-23 |
| digestive | 523.3 | Periodontitis (acute or chronic) | 271618 | 25654 | 1.02 (1.01,1.02) | 2.4E-33 | 1.01 (1.01,1.01) | 1.7E-13 | 1.01 (1.01,1.01) | 8.3E-19 |
| digestive | 525 | Other diseases of the teeth and supporting structures | 230287 | 60680 | 1.01 (1.01,1.02) | 8.1E-53 | 1.01 (1.01,1.01) | 2.2E-16 | 1.01 (1.01,1.01) | 5.1E-26 |
| digestive | 525.1 | Loss of teeth or edentulism | 261829 | 38073 | 1.01 (1.01,1.02) | 3.1E-35 | 1.01 (1,1.01) | 1.2E-10 | 1.01 (1.01,1.01) | 6.0E-17 |
| genitourinary | 599 | Other symptoms/disorders or the urinary system | 215307 | 59968 | 1.01 (1.01,1.01) | 2.9E-26 | 1.01 (1,1.01) | 7.2E-09 | 1.01 (1.01,1.01) | 2.2E-15 |
| infectious diseases | 70.3 | Viral hepatitis C | 294978 | 13935 | 1.02 (1.02,1.02) | 1.2E-32 | 1.02 (1.01,1.02) | 4.2E-20 | 1.02 (1.01,1.02) | 4.1E-26 |
| mental disorders | 290.3 | Other persistent mental disorders due to conditions classified elsewhere | 288225 | 14474 | 1.02 (1.01,1.02) | 6.5E-26 | 1.01 (1.01,1.01) | 1.7E-08 | 1.01 (1.01,1.02) | 1.2E-12 |
| mental disorders | 292 | Neurological disorders | 246745 | 40721 | 1.01 (1.01,1.02) | 1.8E-40 | 1.01 (1,1.01) | 1.1E-11 | 1.01 (1.01,1.01) | 2.2E-19 |
| mental disorders | 296 | Mood disorders | 161370 | 132442 | 1.02 (1.02,1.02) | 7.0E-193 | 1 (0.99,1.01) | 9.0E-01 | 1.02 (1.02,1.02) | 1.4E-121 |
| mental disorders | 296.1 | Bipolar | 285708 | 20399 | 1.05 (1.05,1.06) | 7.1E-255 | 1.04 (1.04,1.05) | 1.2E-171 | 1.05 (1.04,1.05) | 3.6E-154 |
| mental disorders | 296.2 | Depression | 167868 | 124718 | 1.02 (1.02,1.02) | 1.1E-138 | 0.99 (0.98,0.99) | 5.9E-11 | 1.02 (1.01,1.02) | 7.3E-86 |
| mental disorders | 296.22 | Major depressive disorder | 200712 | 92217 | 1.02 (1.02,1.02) | 4.5E-97 | 1 (1,1) | 9.8E-01 | 1.01 (1.01,1.01) | 9.6E-54 |
| mental disorders | 297 | Suicidal ideation or attempt | 285894 | 14832 | 1.03 (1.03,1.04) | 6.5E-85 | 1.03 (1.02,1.03) | 3.5E-46 | 1.03 (1.02,1.03) | 2.2E-47 |
| mental disorders | 297.1 | Suicidal ideation | 291719 | 12317 | 1.04 (1.03,1.04) | 9.0E-80 | 1.03 (1.02,1.03) | 3.0E-44 | 1.03 (1.02,1.03) | 5.2E-45 |
| mental disorders | 300 | Anxiety disorders | 166912 | 123000 | 1.02 (1.02,1.02) | 1.4E-137 | 1.01 (1.01,1.01) | 4.6E-22 | 1.02 (1.01,1.02) | 5.9E-83 |
| mental disorders | 300.1 | Anxiety disorder | 206217 | 79142 | 1.02 (1.02,1.02) | 1.2E-132 | 1.01 (1.01,1.01) | 2.2E-39 | 1.02 (1.02,1.02) | 4.3E-84 |
| mental disorders | 300.11 | Generalized anxiety disorder | 278052 | 21853 | 1.02 (1.02,1.02) | 2.2E-37 | 1.01 (1.01,1.01) | 1.8E-11 | 1.01 (1.01,1.02) | 3.7E-22 |
| mental disorders | 300.4 | Dysthymic disorder | 274401 | 23556 | 1.02 (1.01,1.02) | 1.7E-30 | 1 (1,1.01) | 2.7E-04 | 1.01 (1.01,1.01) | 5.8E-15 |
| mental disorders | 300.9 | Posttraumatic stress disorder | 230809 | 69537 | 1.01 (1.01,1.01) | 2.5E-47 | 1 (1,1) | 1.0E-03 | 1.01 (1.01,1.01) | 1.2E-19 |
| mental disorders | 301 | Personality disorders | 290676 | 13909 | 1.04 (1.03,1.04) | 8.9E-89 | 1.03 (1.02,1.03) | 2.3E-50 | 1.03 (1.02,1.03) | 2.5E-50 |
| mental disorders | 301.2 | Antisocial/borderline personality disorder | 303440 | 5191 | 1.04 (1.03,1.04) | 1.2E-38 | 1.03 (1.02,1.04) | 1.2E-23 | 1.03 (1.02,1.03) | 2.9E-20 |
| mental disorders | 304 | Adjustment reaction | 238220 | 47151 | 1.01 (1.01,1.02) | 2.6E-39 | 1 (1,1.01) | 7.5E-06 | 1.01 (1.01,1.01) | 9.6E-24 |
| mental disorders | 306 | Other mental disorder | 257665 | 23904 | 1.02 (1.02,1.02) | 8.0E-41 | 1.01 (1.01,1.01) | 5.6E-14 | 1.01 (1.01,1.02) | 2.7E-20 |
| mental disorders | 316 | Substance addiction and disorders | 261155 | 36576 | 1.03 (1.03,1.03) | 6.0E-121 | 1.02 (1.02,1.02) | 6.3E-62 | 1.02 (1.02,1.03) | 3.2E-78 |
| mental disorders | 317.1 | Alcoholism | 255939 | 39754 | 1.02 (1.02,1.02) | 3.4E-64 | 1.01 (1.01,1.01) | 1.3E-28 | 1.02 (1.01,1.02) | 5.2E-42 |
| mental disorders | 318 | Tobacco use disorder | 175890 | 103781 | 1.01 (1.01,1.01) | 4.9E-34 | 1.01 (1,1.01) | 9.6E-12 | 1.01 (1.01,1.01) | 2.2E-21 |
| neurological | 327.4 | Insomnia | 230277 | 51649 | 1.01 (1.01,1.01) | 1.4E-26 | 1 (1,1.01) | 8.2E-04 | 1.01 (1,1.01) | 5.9E-12 |
| respiratory | 465 | Acute upper respiratory infections of multiple or unspecified sites | 204993 | 53253 | 1.01 (1.01,1.02) | 3.5E-49 | 1.01 (1.01,1.01) | 6.1E-22 | 1.01 (1.01,1.01) | 6.0E-30 |

**Supplemental Table 5. Significant findings ($p<10^{-25}$) in PheWAS of DEP PRS in EA participants.**

| Group | Phecode | Description | Cont | Case | Base | | Diagnosis-adjusted | | Medication-adjusted | |
|---|---|---|---|---|---|---|---|---|---|---|
| | | | | | OR (95% CI) | *p*-value | OR (95% CI) | *p*-value | OR (95% CI) | *p*-value |
| circulatory system | 401 | Hypertension | 69818 | 225389 | 1.06 (1.05,1.07) | 7.3E-40 | 1.04 (1.03,1.05) | 2.5E-19 | 1.04 (1.03,1.05) | 5.9E-17 |
| circulatory system | 401.1 | Essential hypertension | 70815 | 224179 | 1.06 (1.05,1.07) | 2.4E-40 | 1.04 (1.03,1.05) | 8.1E-20 | 1.04 (1.03,1.05) | 2.3E-17 |
| circulatory system | 411 | Ischemic Heart Disease | 187667 | 100858 | 1.08 (1.07,1.09) | 4.2E-71 | 1.06 (1.05,1.07) | 2.2E-42 | 1.06 (1.05,1.07) | 3.1E-39 |
| circulatory system | 411.1 | Unstable angina | 287650 | 11889 | 1.12 (1.1,1.14) | 1.8E-33 | 1.09 (1.07,1.11) | 1.6E-20 | 1.09 (1.07,1.11) | 1.9E-19 |
| circulatory system | 411.2 | Myocardial infarction | 270506 | 27694 | 1.08 (1.06,1.09) | 1.5E-29 | 1.06 (1.04,1.07) | 2.5E-17 | 1.05 (1.04,1.07) | 1.4E-16 |
| circulatory system | 411.3 | Angina pectoris | 265965 | 26054 | 1.08 (1.07,1.1) | 2.5E-34 | 1.06 (1.05,1.07) | 3.3E-18 | 1.06 (1.04,1.07) | 2.3E-17 |
| circulatory system | 411.4 | Coronary atherosclerosis | 205059 | 88022 | 1.07 (1.06,1.08) | 6.1E-57 | 1.05 (1.05,1.06) | 1.1E-34 | 1.05 (1.04,1.06) | 5.1E-32 |
| circulatory system | 411.8 | Other chronic ischemic heart disease | 242159 | 53680 | 1.07 (1.06,1.08) | 8.7E-40 | 1.05 (1.04,1.06) | 2.5E-24 | 1.05 (1.04,1.06) | 4.6E-22 |
| circulatory system | 418 | Nonspecific chest pain | 178521 | 91397 | 1.1 (1.09,1.11) | 7.9E-125 | 1.07 (1.06,1.08) | 1.8E-59 | 1.07 (1.06,1.08) | 3.4E-56 |
| circulatory system | 427 | Cardiac dysrhythmias | 168977 | 104053 | 1.05 (1.04,1.06) | 5.6E-37 | 1.03 (1.02,1.04) | 2.2E-15 | 1.03 (1.02,1.04) | 3.2E-14 |
| circulatory system | 428 | Congestive heart failure; nonhypertensive | 254758 | 41526 | 1.08 (1.07,1.09) | 7.9E-42 | 1.05 (1.04,1.07) | 6.8E-22 | 1.05 (1.04,1.06) | 5.4E-21 |
| circulatory system | 433 | Cerebrovascular disease | 239598 | 49889 | 1.06 (1.05,1.07) | 5.8E-29 | 1.03 (1.02,1.04) | 5.3E-11 | 1.03 (1.02,1.04) | 7.0E-11 |
| circulatory system | 458 | Hypotension | 256763 | 27983 | 1.08 (1.07,1.1) | 6.6E-37 | 1.05 (1.03,1.06) | 2.4E-12 | 1.05 (1.03,1.06) | 4.4E-12 |
| circulatory system | 458.9 | Hypotension NOS | 273868 | 16517 | 1.09 (1.08,1.11) | 1.3E-27 | 1.06 (1.04,1.07) | 2.1E-11 | 1.06 (1.04,1.07) | 3.2E-11 |
| dermatologic | 681 | Superficial cellulitis and abscess | 224657 | 52705 | 1.06 (1.05,1.07) | 5.6E-30 | 1.03 (1.02,1.04) | 3.7E-09 | 1.03 (1.02,1.04) | 1.0E-08 |
| digestive | 521 | Diseases of hard tissues of teeth | 233809 | 60055 | 1.07 (1.06,1.08) | 1.1E-46 | 1.03 (1.02,1.04) | 1.3E-10 | 1.03 (1.02,1.04) | 7.9E-11 |
| digestive | 521.1 | Dental caries | 236323 | 58432 | 1.07 (1.06,1.08) | 4.4E-46 | 1.03 (1.02,1.04) | 1.2E-10 | 1.03 (1.02,1.04) | 7.0E-11 |
| digestive | 523 | Gingival and periodontal diseases | 246246 | 49998 | 1.07 (1.06,1.08) | 1.6E-38 | 1.03 (1.02,1.04) | 1.4E-08 | 1.03 (1.02,1.04) | 1.5E-08 |
| digestive | 525 | Other diseases of the teeth | 230287 | 60680 | 1.08 (1.07,1.09) | 2.9E-67 | 1.04 (1.03,1.05) | 3.3E-19 | 1.04 (1.03,1.05) | 8.3E-20 |
| digestive | 525.1 | Loss of teeth or edentulism | 261829 | 38073 | 1.08 (1.07,1.09) | 7.5E-43 | 1.04 (1.03,1.05) | 5.6E-11 | 1.04 (1.03,1.05) | 4.4E-11 |
| digestive | 530 | Diseases of esophagus | 153615 | 131627 | 1.08 (1.07,1.09) | 2.3E-84 | 1.05 (1.04,1.06) | 7.0E-38 | 1.05 (1.04,1.06) | 5.3E-34 |
| digestive | 530.1 | Esophagitis, GERD and related diseases | 156612 | 128556 | 1.08 (1.07,1.09) | 2.7E-84 | 1.05 (1.04,1.06) | 5.4E-38 | 1.05 (1.04,1.06) | 4.3E-34 |
| digestive | 530.11 | GERD | 162469 | 122039 | 1.08 (1.07,1.09) | 3.3E-84 | 1.05 (1.04,1.06) | 2.0E-38 | 1.05 (1.04,1.06) | 2.0E-34 |
| digestive | 532 | Dysphagia | 252740 | 40103 | 1.07 (1.05,1.08) | 3.7E-31 | 1.03 (1.02,1.05) | 4.2E-10 | 1.03 (1.02,1.04) | 2.5E-09 |
| digestive | 561 | Symptoms involving digestive system | 233209 | 41830 | 1.08 (1.07,1.09) | 5.2E-46 | 1.05 (1.03,1.06) | 9.0E-16 | 1.04 (1.03,1.06) | 1.4E-15 |
| digestive | 578 | Gastrointestinal hemorrhage | 250313 | 35111 | 1.07 (1.05,1.08) | 2.2E-28 | 1.04 (1.03,1.05) | 3.4E-12 | 1.04 (1.03,1.05) | 1.7E-11 |
| endocrine/metabolic | 250 | Diabetes mellitus | 189084 | 108545 | 1.05 (1.04,1.05) | 1.8E-30 | 1.03 (1.02,1.04) | 1.9E-15 | 1.03 (1.02,1.04) | 2.8E-13 |
| endocrine/metabolic | 250.2 | Type 2 diabetes | 190540 | 107822 | 1.04 (1.04,1.05) | 4.8E-29 | 1.03 (1.02,1.04) | 1.5E-14 | 1.03 (1.02,1.04) | 1.9E-12 |
| endocrine/metabolic | 250.24 | T2D with neurological manifestations | 259077 | 40530 | 1.06 (1.05,1.08) | 1.1E-30 | 1.04 (1.03,1.05) | 1.6E-13 | 1.04 (1.02,1.05) | 2.2E-10 |
| endocrine/metabolic | 276 | Disorders of fluid, electrolyte, acid-base | 221908 | 54151 | 1.07 (1.06,1.08) | 2.1E-43 | 1.04 (1.03,1.05) | 3.4E-15 | 1.04 (1.03,1.05) | 1.2E-14 |
| endocrine/metabolic | 276.1 | Electrolyte imbalance | 242364 | 39794 | 1.06 (1.05,1.07) | 5.9E-26 | 1.03 (1.02,1.04) | 2.9E-09 | 1.03 (1.02,1.04) | 4.4E-09 |
| genitourinary | 585 | Renal failure | 230047 | 61932 | 1.05 (1.04,1.06) | 7.3E-26 | 1.03 (1.02,1.04) | 1.3E-10 | 1.03 (1.02,1.04) | 1.7E-09 |
| genitourinary | 585.1 | Acute renal failure | 263505 | 29679 | 1.07 (1.06,1.08) | 3.8E-28 | 1.04 (1.03,1.06) | 4.4E-11 | 1.04 (1.03,1.05) | 1.5E-10 |
| genitourinary | 591 | Urinary tract infection | 252052 | 33029 | 1.07 (1.06,1.08) | 6.3E-30 | 1.04 (1.03,1.06) | 6.2E-12 | 1.04 (1.03,1.06) | 8.8E-12 |
| genitourinary | 599 | Disorders or the urinary system | 215307 | 59968 | 1.05 (1.04,1.06) | 4.8E-29 | 1.03 (1.02,1.04) | 7.7E-08 | 1.03 (1.02,1.04) | 8.4E-08 |
| genitourinary | 600 | Hyperplasia of prostate | 189050 | 97473 | 1.05 (1.04,1.06) | 5.4E-27 | 1.03 (1.02,1.04) | 1.9E-10 | 1.03 (1.02,1.04) | 4.4E-10 |
| hematopoietic | 285 | Other anemias | 223129 | 59404 | 1.05 (1.04,1.06) | 1.0E-26 | 1.03 (1.02,1.04) | 7.8E-10 | 1.03 (1.02,1.04) | 4.3E-09 |
| mental disorders | 292 | Neurological disorders | 246745 | 40721 | 1.07 (1.06,1.08) | 6.4E-33 | 1.02 (1.01,1.03) | 2.3E-04 | 1.03 (1.01,1.04) | 3.8E-06 |
| mental disorders | 296 | Mood disorders | 161370 | 132442 | 1.15 (1.14,1.16) | 7.6E-271 | 1.08 (1.03,1.12) | 2.8E-04 | 1.1 (1.09,1.11) | 5.4E-68 |
| mental disorders | 296.1 | Bipolar | 285708 | 20399 | 1.16 (1.15,1.18) | 8.2E-91 | 1.1 (1.08,1.12) | 1.9E-35 | 1.12 (1.1,1.14) | 2.9E-49 |
| mental disorders | 296.2 | Depression | 167868 | 124718 | 1.15 (1.14,1.16) | 3.2E-257 | 1.05 (1.03,1.07) | 1.9E-05 | 1.09 (1.08,1.1) | 1.2E-59 |
| mental disorders | 296.22 | Major depressive disorder | 200712 | 92217 | 1.14 (1.13,1.15) | 1.1E-205 | 1.05 (1.04,1.07) | 6.0E-18 | 1.08 (1.07,1.09) | 1.6E-47 |
| mental disorders | 297 | Suicidal ideation or attempt | 285894 | 14832 | 1.16 (1.14,1.18) | 1.9E-64 | 1.1 (1.08,1.12) | 1.5E-26 | 1.11 (1.09,1.13) | 1.3E-30 |
| mental disorders | 297.1 | Suicidal ideation | 291719 | 12317 | 1.16 (1.14,1.18) | 1.5E-53 | 1.1 (1.08,1.12) | 1.6E-21 | 1.1 (1.08,1.13) | 6.0E-25 |
| mental disorders | 300.1 | Anxiety disorder | 206217 | 79142 | 1.12 (1.11,1.13) | 6.4E-137 | 1.06 (1.05,1.07) | 2.1E-29 | 1.06 (1.05,1.07) | 2.5E-33 |

| | | | | | | | | | | |
|---|---|---|---|---|---|---|---|---|---|---|
| mental disorders | 300.11 | Generalized anxiety disorder | 278052 | 21853 | 1.1 (1.08,1.11) | 8.0E-37 | 1.04 (1.03,1.06) | 4.2E-08 | 1.04 (1.03,1.06) | 8.5E-09 |
| mental disorders | 300.12 | Agorophobia, social phobia, & panic dis | 292966 | 12188 | 1.12 (1.1,1.14) | 7.2E-33 | 1.07 (1.05,1.09) | 4.2E-12 | 1.07 (1.05,1.09) | 4.1E-12 |
| mental disorders | 300.4 | Dysthymic disorder | 274401 | 23556 | 1.12 (1.11,1.14) | 1.3E-63 | 1.06 (1.04,1.07) | 1.2E-14 | 1.07 (1.05,1.08) | 5.0E-19 |
| mental disorders | 300.9 | Posttraumatic stress disorder | 230809 | 69537 | 1.1 (1.09,1.11) | 6.2E-99 | 1.04 (1.03,1.05) | 8.7E-18 | 1.05 (1.04,1.06) | 2.7E-22 |
| mental disorders | 301 | Personality disorders | 290676 | 13909 | 1.18 (1.16,1.2) | 2.3E-75 | 1.12 (1.1,1.14) | 3.4E-35 | 1.13 (1.11,1.15) | 1.6E-40 |
| mental disorders | 301.2 | Antisocial/borderline personality disorder | 303440 | 5191 | 1.2 (1.16,1.23) | 4.1E-35 | 1.14 (1.11,1.17) | 4.7E-19 | 1.15 (1.11,1.18) | 6.1E-21 |
| mental disorders | 304 | Adjustment reaction | 238220 | 47151 | 1.08 (1.07,1.09) | 4.5E-46 | 1.02 (1.01,1.03) | 2.3E-05 | 1.03 (1.02,1.04) | 5.8E-09 |
| mental disorders | 306 | Other mental disorder | 257665 | 23904 | 1.1 (1.08,1.11) | 4.5E-41 | 1.05 (1.03,1.06) | 6.7E-11 | 1.05 (1.04,1.07) | 3.2E-13 |
| mental disorders | 316 | Substance addiction and disorders | 261155 | 36576 | 1.12 (1.11,1.14) | 2.8E-87 | 1.07 (1.06,1.09) | 3.8E-31 | 1.08 (1.07,1.09) | 2.5E-36 |
| mental disorders | 317.1 | Alcoholism | 255939 | 39754 | 1.09 (1.08,1.1) | 3.5E-52 | 1.05 (1.04,1.06) | 1.2E-16 | 1.05 (1.04,1.07) | 1.6E-20 |
| mental disorders | 318 | Tobacco use disorder | 175890 | 103781 | 1.09 (1.08,1.1) | 4.8E-97 | 1.06 (1.05,1.07) | 1.2E-49 | 1.06 (1.05,1.07) | 2.1E-50 |
| musculoskeletal | 716 | Other arthropathies | 247336 | 33669 | 1.06 (1.05,1.08) | 3.5E-26 | 1.04 (1.03,1.05) | 1.5E-11 | 1.04 (1.02,1.05) | 1.1E-09 |
| musculoskeletal | 721 | Spondylosis and allied disorders | 238955 | 44356 | 1.07 (1.06,1.08) | 4.9E-37 | 1.04 (1.03,1.05) | 1.4E-13 | 1.03 (1.02,1.04) | 4.0E-10 |
| musculoskeletal | 721.1 | Spondylosis without myelopathy | 242860 | 40805 | 1.07 (1.06,1.08) | 7.5E-34 | 1.04 (1.03,1.05) | 2.3E-12 | 1.03 (1.02,1.04) | 3.0E-09 |
| musculoskeletal | 722 | Intervertebral disc disorders | 228596 | 55337 | 1.07 (1.06,1.08) | 2.5E-44 | 1.04 (1.03,1.05) | 3.4E-17 | 1.04 (1.03,1.05) | 1.2E-12 |
| musculoskeletal | 722.6 | Degeneration of intervertebral disc | 242906 | 42226 | 1.06 (1.05,1.07) | 8.6E-31 | 1.04 (1.03,1.05) | 2.3E-11 | 1.03 (1.02,1.04) | 3.1E-08 |
| musculoskeletal | 740 | Osteoarthrosis | 146222 | 131442 | 1.06 (1.05,1.06) | 4.1E-46 | 1.03 (1.03,1.04) | 1.1E-17 | 1.03 (1.02,1.04) | 3.4E-14 |
| musculoskeletal | 740.9 | Osteoarthrosis NOS | 179236 | 99593 | 1.07 (1.06,1.07) | 8.8E-56 | 1.04 (1.03,1.05) | 5.8E-24 | 1.04 (1.03,1.05) | 3.3E-20 |
| musculoskeletal | 745 | Pain in joint | 89094 | 187207 | 1.07 (1.06,1.07) | 2.2E-52 | 1.04 (1.03,1.04) | 7.1E-17 | 1.03 (1.02,1.04) | 2.8E-14 |
| neurological | 327 | Sleep disorders | 191386 | 81129 | 1.08 (1.07,1.09) | 9.4E-70 | 1.03 (1.03,1.04) | 7.2E-14 | 1.03 (1.02,1.04) | 1.5E-12 |
| neurological | 327.3 | Sleep apnea | 195189 | 101623 | 1.04 (1.04,1.05) | 6.0E-28 | 1.02 (1.01,1.03) | 2.2E-05 | 1.02 (1.01,1.02) | 3.5E-05 |
| neurological | 327.4 | Insomnia | 230277 | 51649 | 1.07 (1.06,1.08) | 1.4E-44 | 1.03 (1.02,1.04) | 4.2E-08 | 1.03 (1.02,1.04) | 1.4E-07 |
| neurological | 338 | Pain | 225658 | 47039 | 1.09 (1.08,1.1) | 4.6E-66 | 1.05 (1.04,1.07) | 6.5E-24 | 1.05 (1.04,1.06) | 4.2E-21 |
| neurological | 338.2 | Chronic pain | 248178 | 34632 | 1.1 (1.09,1.11) | 1.8E-57 | 1.06 (1.05,1.07) | 4.7E-21 | 1.05 (1.04,1.07) | 1.7E-18 |
| neurological | 339 | Other headache syndromes | 237942 | 42426 | 1.09 (1.08,1.1) | 1.1E-57 | 1.05 (1.04,1.07) | 2.6E-21 | 1.05 (1.04,1.06) | 5.3E-18 |
| neurological | 350 | Abnormal movement | 220347 | 57430 | 1.07 (1.06,1.08) | 5.0E-43 | 1.03 (1.02,1.04) | 2.1E-09 | 1.03 (1.02,1.04) | 1.3E-09 |
| neurological | 350.2 | Abnormality of gait | 238406 | 44292 | 1.07 (1.06,1.08) | 5.7E-37 | 1.03 (1.02,1.04) | 3.8E-09 | 1.03 (1.02,1.04) | 2.8E-09 |
| neurological | 351 | Other peripheral nerve disorders | 227740 | 57720 | 1.05 (1.05,1.06) | 7.5E-30 | 1.03 (1.02,1.04) | 6.5E-09 | 1.02 (1.01,1.03) | 1.6E-05 |
| neurological | 355.1 | Chronic pain syndrome | 283361 | 17187 | 1.11 (1.09,1.12) | 7.5E-37 | 1.06 (1.05,1.08) | 1.3E-14 | 1.06 (1.04,1.08) | 1.3E-12 |
| respiratory | 465 | Acute upper respiratory infections | 204993 | 53253 | 1.07 (1.06,1.08) | 5.1E-41 | 1.04 (1.03,1.05) | 3.9E-14 | 1.04 (1.03,1.05) | 1.5E-13 |
| respiratory | 480 | Pneumonia | 260383 | 31337 | 1.08 (1.07,1.09) | 2.5E-35 | 1.05 (1.04,1.06) | 1.0E-14 | 1.05 (1.03,1.06) | 5.8E-14 |
| respiratory | 496 | Chronic airway obstruction | 215720 | 72353 | 1.1 (1.09,1.11) | 2.0E-108 | 1.08 (1.07,1.09) | 1.8E-61 | 1.08 (1.07,1.09) | 1.2E-59 |
| respiratory | 496.2 | Chronic bronchitis | 276924 | 19677 | 1.12 (1.1,1.13) | 8.0E-49 | 1.09 (1.07,1.1) | 2.7E-27 | 1.08 (1.07,1.1) | 5.7E-27 |
| respiratory | 496.21 | Obstructive chronic bronchitis | 282188 | 17154 | 1.12 (1.1,1.13) | 2.2E-42 | 1.08 (1.07,1.1) | 2.3E-23 | 1.08 (1.07,1.1) | 4.6E-23 |
| respiratory | 497 | Bronchitis | 268282 | 14750 | 1.09 (1.08,1.11) | 3.9E-26 | 1.06 (1.04,1.08) | 3.9E-12 | 1.06 (1.04,1.08) | 3.6E-12 |
| respiratory | 509 | Respiratory failure, insufficiency, arrest | 275454 | 22821 | 1.09 (1.07,1.1) | 5.5E-32 | 1.06 (1.04,1.07) | 1.2E-15 | 1.06 (1.04,1.07) | 1.2E-14 |
| respiratory | 509.1 | Respiratory failure | 284540 | 17095 | 1.09 (1.08,1.11) | 1.6E-28 | 1.06 (1.05,1.08) | 6.2E-15 | 1.06 (1.05,1.08) | 4.3E-14 |
| respiratory | 512 | Other symptoms of respiratory system | 139662 | 120187 | 1.08 (1.07,1.09) | 3.6E-85 | 1.05 (1.04,1.06) | 5.2E-31 | 1.05 (1.04,1.06) | 9.6E-29 |
| respiratory | 512.7 | Shortness of breath | 209689 | 60907 | 1.08 (1.07,1.09) | 6.3E-60 | 1.05 (1.04,1.06) | 1.7E-24 | 1.05 (1.04,1.06) | 1.5E-22 |
| respiratory | 512.8 | Cough | 212486 | 48953 | 1.06 (1.05,1.07) | 2.7E-27 | 1.03 (1.02,1.04) | 1.8E-08 | 1.03 (1.02,1.04) | 4.2E-08 |
| respiratory | 512.9 | Other dyspnea | 219490 | 48646 | 1.07 (1.06,1.08) | 1.5E-37 | 1.04 (1.03,1.05) | 6.4E-14 | 1.04 (1.03,1.05) | 6.5E-13 |
| sense organs | 366 | Cataract | 116658 | 166416 | 1.05 (1.05,1.06) | 1.7E-37 | 1.03 (1.02,1.04) | 1.1E-13 | 1.03 (1.02,1.04) | 3.0E-12 |
| sense organs | 366.2 | Senile cataract | 139596 | 142445 | 1.04 (1.04,1.05) | 1.2E-27 | 1.02 (1.02,1.03) | 1.7E-09 | 1.02 (1.01,1.03) | 1.4E-08 |
| sense organs | 367 | Blindness and low vision | 86906 | 200705 | 1.05 (1.04,1.06) | 1.6E-35 | 1.03 (1.02,1.03) | 1.4E-09 | 1.02 (1.02,1.03) | 1.7E-08 |
| sense organs | 386.9 | Dizziness and giddiness | 231860 | 43001 | 1.08 (1.07,1.09) | 3.0E-44 | 1.04 (1.03,1.06) | 1.3E-15 | 1.04 (1.03,1.06) | 1.2E-15 |
| symptoms | 760 | Back pain | 128684 | 149486 | 1.09 (1.08,1.1) | 1.5E-105 | 1.06 (1.05,1.07) | 9.5E-44 | 1.05 (1.04,1.06) | 3.1E-37 |
| symptoms | 761 | Cervicalgia | 226921 | 56206 | 1.08 (1.07,1.09) | 3.8E-52 | 1.05 (1.04,1.06) | 1.2E-20 | 1.04 (1.03,1.05) | 4.2E-17 |
| symptoms | 785 | Abdominal pain | 206085 | 63494 | 1.09 (1.08,1.1) | 2.2E-82 | 1.06 (1.05,1.07) | 4.7E-37 | 1.06 (1.05,1.07) | 1.8E-34 |

**Supplemental Table 6. Significant findings ($p<10^{-5}$) in PheWAS of SCZ PRS in AA participants.**

| Group | Phecode | Description | Cont | Case | Base | | Diagnosis-adjusted | | Medication-adjusted | |
|---|---|---|---|---|---|---|---|---|---|---|
| | | | | | OR (95% CI) | *p*-value | OR (95% CI) | *p*-value | OR (95% CI) | *p*-value |
| dermatologic | 700 | Corns and callosities | 68194 | 9528 | 1.07 (1.04,1.1) | 2.8E-06 | 1.06 (1.03,1.09) | 9.2E-05 | 1.06 (1.03,1.09) | 1.5E-04 |
| digestive | 521 | Diseases of hard tissues of teeth | 48819 | 27048 | 1.06 (1.04,1.09) | 3.8E-10 | 1.04 (1.02,1.06) | 1.4E-04 | 1.04 (1.01,1.06) | 6.4E-04 |
| digestive | 521.1 | Dental caries | 49444 | 26480 | 1.06 (1.04,1.09) | 7.3E-10 | 1.04 (1.02,1.06) | 2.1E-04 | 1.04 (1.01,1.06) | 9.2E-04 |
| digestive | 523 | Gingival and periodontal diseases | 53599 | 22773 | 1.06 (1.04,1.08) | 3.8E-08 | 1.03 (1.01,1.06) | 1.5E-03 | 1.03 (1.01,1.05) | 3.4E-03 |
| digestive | 525 | Other diseases of the teeth | 47747 | 27265 | 1.08 (1.06,1.1) | 4.6E-14 | 1.05 (1.03,1.08) | 4.5E-07 | 1.05 (1.03,1.07) | 4.5E-06 |
| digestive | 525.1 | Loss of teeth or edentulism | 62014 | 16377 | 1.06 (1.03,1.08) | 9.7E-07 | 1.03 (1.01,1.06) | 4.3E-03 | 1.03 (1,1.05) | 1.9E-02 |
| infectious diseases | 110 | Dermatophytosis / Dermatomycosis | 45593 | 27229 | 1.05 (1.03,1.07) | 9.9E-07 | 1.04 (1.02,1.06) | 4.2E-04 | 1.04 (1.02,1.06) | 5.1E-04 |
| infectious diseases | 110.1 | Dermatophytosis | 46880 | 26148 | 1.05 (1.03,1.07) | 2.2E-06 | 1.04 (1.02,1.06) | 6.7E-04 | 1.04 (1.01,1.06) | 8.6E-04 |
| mental disorders | 292.6 | Hallucinations | 81498 | 518 | 1.3 (1.16,1.45) | 3.2E-06 | 1.26 (1.12,1.4) | 5.2E-05 | 1.21 (1.08,1.35) | 9.1E-04 |
| mental disorders | 296 | Mood disorders | 33552 | 44186 | 1.09 (1.06,1.11) | 8.3E-17 | 0.84 (0.78,0.9) | 2.1E-06 | 1.05 (1.03,1.07) | 2.1E-06 |
| mental disorders | 296.1 | Bipolar | 75000 | 5921 | 1.11 (1.07,1.15) | 2.2E-09 | 1.08 (1.04,1.12) | 5.2E-05 | 1.04 (1,1.08) | 4.2E-02 |
| mental disorders | 296.2 | Depression | 35260 | 42196 | 1.08 (1.05,1.1) | 9.2E-14 | 0.9 (0.86,0.95) | 6.8E-05 | 1.04 (1.02,1.07) | 3.8E-05 |
| mental disorders | 296.22 | Major depressive disorder | 44614 | 32643 | 1.05 (1.03,1.07) | 1.4E-07 | 0.97 (0.94,1) | 3.7E-02 | 1.02 (1,1.04) | 7.0E-02 |
| mental disorders | 297 | Suicidal ideation or attempt | 72688 | 6386 | 1.1 (1.06,1.14) | 6.7E-08 | 1.06 (1.02,1.1) | 1.3E-03 | 1.03 (1,1.07) | 7.8E-02 |
| mental disorders | 297.1 | Suicidal ideation | 74668 | 5503 | 1.1 (1.06,1.14) | 6.6E-07 | 1.06 (1.02,1.1) | 4.0E-03 | 1.03 (0.99,1.07) | 1.6E-01 |
| mental disorders | 300 | Anxiety disorders | 36301 | 40111 | 1.06 (1.04,1.08) | 2.5E-08 | 1 (0.98,1.03) | 7.9E-01 | 1.02 (1,1.05) | 2.2E-02 |
| mental disorders | 300.1 | Anxiety disorder | 51938 | 21915 | 1.06 (1.04,1.08) | 6.1E-08 | 1.02 (1,1.05) | 5.2E-02 | 1.03 (1.01,1.06) | 5.2E-03 |
| mental disorders | 301 | Personality disorders | 75463 | 4935 | 1.1 (1.05,1.14) | 2.2E-06 | 1.06 (1.02,1.1) | 3.4E-03 | 1.03 (0.99,1.07) | 1.7E-01 |
| mental disorders | 304 | Adjustment reaction | 55082 | 18851 | 1.05 (1.03,1.08) | 2.7E-06 | 1.02 (1,1.04) | 8.2E-02 | 1.03 (1.01,1.06) | 2.9E-03 |
| mental disorders | 316 | Substance addiction and disorders | 55813 | 22948 | 1.08 (1.06,1.1) | 1.7E-13 | 1.05 (1.03,1.07) | 6.2E-06 | 1.04 (1.02,1.07) | 8.4E-05 |
| mental disorders | 317.1 | Alcoholism | 58054 | 19278 | 1.07 (1.04,1.09) | 5.9E-09 | 1.04 (1.01,1.06) | 1.6E-03 | 1.03 (1.01,1.06) | 4.0E-03 |
| respiratory | 465 | Acute upper respiratory infections | 46098 | 21217 | 1.07 (1.05,1.09) | 1.7E-09 | 1.05 (1.03,1.07) | 1.5E-05 | 1.05 (1.03,1.07) | 2.8E-05 |

**Supplemental Table 7. Significant findings ($p$<$10^{-5}$) in PheWAS of BIP PRS in AA participants.**

| Group | Phecode | Description | Cont | Case | Base | | Diagnosis-adjusted | | Medication-adjusted | |
|---|---|---|---|---|---|---|---|---|---|---|
| | | | | | OR (95% CI) | *p*-value | OR (95% CI) | *p*-value | OR (95% CI) | *p*-value |
| digestive | 521 | Diseases of hard tissues of teeth | 48819 | 27048 | 1.01 (1,1.01) | 9.2E-11 | 1.01 (1,1.01) | 1.7E-06 | 1.01 (1,1.01) | 3.6E-08 |
| digestive | 521.1 | Dental caries | 49444 | 26480 | 1.01 (1,1.01) | 1.2E-09 | 1 (1,1.01) | 1.1E-05 | 1.01 (1,1.01) | 3.2E-07 |
| digestive | 523 | Gingival and periodontal diseases | 53599 | 22773 | 1.01 (1,1.01) | 2.2E-07 | 1 (1,1.01) | 3.8E-04 | 1 (1,1.01) | 2.3E-05 |
| digestive | 525 | Other diseases of the teeth | 47747 | 27265 | 1.01 (1,1.01) | 1.0E-09 | 1 (1,1.01) | 2.6E-05 | 1.01 (1,1.01) | 2.8E-07 |
| digestive | 525.1 | Loss of teeth or edentulism | 62014 | 16377 | 1.01 (1,1.01) | 3.9E-06 | 1 (1,1.01) | 2.1E-03 | 1 (1,1.01) | 1.4E-04 |
| infectious diseases | 110 | Dermatophytosis / Dermatomycosis | 45593 | 27229 | 1.01 (1,1.01) | 5.8E-07 | 1 (1,1.01) | 5.7E-05 | 1 (1,1.01) | 9.9E-06 |
| infectious diseases | 110.1 | Dermatophytosis | 46880 | 26148 | 1.01 (1,1.01) | 7.6E-07 | 1 (1,1.01) | 6.3E-05 | 1 (1,1.01) | 1.4E-05 |
| mental disorders | 296 | Mood disorders | 33552 | 44186 | 1.01 (1,1.01) | 4.6E-09 | 0.99 (0.98,1) | 3.3E-03 | 1 (1,1.01) | 5.2E-06 |
| mental disorders | 296.1 | Bipolar | 75000 | 5921 | 1.01 (1.01,1.02) | 8.5E-11 | 1.01 (1.01,1.01) | 2.3E-07 | 1.01 (1.01,1.01) | 1.9E-06 |
| mental disorders | 296.2 | Depression | 35260 | 42196 | 1.01 (1,1.01) | 3.3E-07 | 0.99 (0.99,1) | 1.4E-03 | 1 (1,1.01) | 6.5E-05 |
| mental disorders | 297 | Suicidal ideation or attempt | 72688 | 6386 | 1.01 (1.01,1.01) | 8.5E-08 | 1.01 (1,1.01) | 8.0E-05 | 1.01 (1,1.01) | 3.4E-05 |
| mental disorders | 297.1 | Suicidal ideation | 74668 | 5503 | 1.01 (1.01,1.01) | 2.7E-06 | 1.01 (1,1.01) | 7.4E-04 | 1.01 (1,1.01) | 3.9E-04 |
| mental disorders | 300 | Anxiety disorders | 36301 | 40111 | 1 (1,1.01) | 4.0E-06 | 1 (1,1) | 5.0E-01 | 1 (1,1.01) | 8.4E-04 |
| mental disorders | 301 | Personality disorders | 75463 | 4935 | 1.01 (1.01,1.01) | 5.2E-07 | 1.01 (1,1.01) | 1.5E-04 | 1.01 (1,1.01) | 9.3E-05 |
| mental disorders | 306 | Other mental disorder | 62819 | 9537 | 1.01 (1,1.01) | 1.7E-06 | 1.01 (1,1.01) | 1.1E-03 | 1.01 (1,1.01) | 1.8E-04 |
| mental disorders | 316 | Substance addiction and disorders | 55813 | 22948 | 1.01 (1,1.01) | 7.3E-09 | 1 (1,1.01) | 1.6E-04 | 1.01 (1,1.01) | 1.7E-06 |
| mental disorders | 317.1 | Alcoholism | 58054 | 19278 | 1.01 (1,1.01) | 6.1E-07 | 1 (1,1.01) | 1.3E-03 | 1 (1,1.01) | 5.1E-05 |
| mental disorders | 318 | Tobacco use disorder | 43079 | 33056 | 1.01 (1,1.01) | 4.1E-07 | 1 (1,1.01) | 1.7E-04 | 1 (1,1.01) | 7.2E-06 |
| respiratory | 465 | Acute upper respiratory infections | 46098 | 21217 | 1.01 (1,1.01) | 1.8E-06 | 1 (1,1.01) | 4.2E-04 | 1 (1,1.01) | 5.4E-05 |
| respiratory | 512.8 | Cough | 54075 | 14679 | 1.01 (1,1.01) | 9.2E-06 | 1 (1,1.01) | 3.9E-04 | 1.01 (1,1.01) | 8.3E-05 |

**Supplemental Table 8. Significant findings ($p$<$10^{-5}$) in PheWAS of DEP PRS in AA participants.** Displayed results are for PRS constructed from SNPs with $p$<$10^{-5}$.

| Group | Phecode | Description | Cont | Case | Base | | Diagnosis-adjusted | | Medication-adjusted | |
|---|---|---|---|---|---|---|---|---|---|---|
| | | | | | OR (95% CI) | *p*-value | OR (95% CI) | *p*-value | OR (95% CI) | *p*-value |
| mental disorders | 296 | Mood disorders | 33552 | 44186 | 1.04 (1.02,1.05) | 3.8E-07 | 0.97 (0.91,1.02) | 2.5E-01 | 1.03 (1.01,1.06) | 5.3E-04 |
| mental disorders | 296.2 | Depression | 35260 | 42196 | 1.04 (1.02,1.05) | 1.4E-06 | 0.99 (0.95,1.02) | 4.9E-01 | 1.03 (1.01,1.05) | 2.0E-03 |
| mental disorders | 300 | Anxiety disorders | 36301 | 40111 | 1.04 (1.02,1.05) | 2.6E-06 | 1.02 (1,1.04) | 7.5E-02 | 1.03 (1.01,1.05) | 1.8E-03 |
| mental disorders | 300.1 | Anxiety disorder | 51938 | 21915 | 1.04 (1.02,1.06) | 3.6E-06 | 1.02 (1,1.04) | 1.4E-02 | 1.03 (1.01,1.05) | 5.9E-04 |
| mental disorders | 300.9 | Posttraumatic stress disorder | 52155 | 26603 | 1.03 (1.02,1.05) | 7.7E-06 | 1.02 (1,1.04) | 1.8E-02 | 1.03 (1.01,1.05) | 1.2E-03 |
| mental disorders | 306 | Other mental disorder | 62819 | 9537 | 1.05 (1.03,1.08) | 4.7E-06 | 1.04 (1.02,1.06) | 5.4E-04 | 1.04 (1.02,1.07) | 1.2E-04 |
| respiratory | 512 | Other respiratory symptoms | 34852 | 33303 | 1.04 (1.02,1.05) | 4.4E-06 | 1.03 (1.01,1.05) | 2.5E-04 | 1.03 (1.02,1.05) | 6.4E-05 |

### Supplemental Table 9. Significant findings ($p<10^{-25}$) in PheWAS of SCZ-specific PRS in EA participants.

| Group | Phecode | Description | Cont | Case | Base OR (95% CI) | Base *p*-value | Diagnosis-adjusted OR (95% CI) | Diagnosis-adjusted *p*-value | Medication-adjusted OR (95% CI) | Medication-adjusted *p*-value |
|---|---|---|---|---|---|---|---|---|---|---|
| circulatory system | 411 | Ischemic Heart Disease | 187667 | 100858 | 0.98 (0.97,0.98) | 1.9E-26 | 0.98 (0.97,0.98) | 5.5E-23 | 0.98 (0.97,0.98) | 8.0E-29 |
| digestive | 530 | Diseases of esophagus | 153615 | 131627 | 0.98 (0.97,0.98) | 1.9E-32 | 0.98 (0.97,0.98) | 2.3E-26 | 0.97 (0.97,0.98) | 1.9E-36 |
| digestive | 530.1 | Esophagitis, GERD and related diseases | 156612 | 128556 | 0.98 (0.97,0.98) | 1.1E-34 | 0.98 (0.97,0.98) | 2.0E-28 | 0.97 (0.97,0.98) | 1.0E-38 |
| digestive | 530.11 | GERD | 162469 | 122039 | 0.98 (0.97,0.98) | 2.3E-35 | 0.98 (0.97,0.98) | 5.2E-29 | 0.97 (0.97,0.98) | 2.6E-39 |
| mental disorders | 296.22 | Major depressive disorder | 200712 | 92217 | 0.98 (0.97,0.98) | 4.2E-29 | 0.97 (0.97,0.98) | 4.7E-18 | 0.97 (0.96,0.97) | 7.4E-41 |
| musculoskeletal | 721 | Spondylosis and allied disorders | 238955 | 44356 | 0.97 (0.96,0.97) | 1.4E-29 | 0.97 (0.97,0.98) | 3.0E-25 | 0.97 (0.96,0.97) | 2.0E-32 |
| musculoskeletal | 721.1 | Spondylosis without myelopathy | 242860 | 40805 | 0.97 (0.96,0.97) | 1.6E-30 | 0.97 (0.96,0.98) | 4.3E-26 | 0.97 (0.96,0.97) | 2.4E-33 |
| musculoskeletal | 722 | Intervertebral disc disorders | 228596 | 55337 | 0.97 (0.96,0.97) | 1.1E-41 | 0.97 (0.96,0.97) | 1.3E-36 | 0.96 (0.96,0.97) | 2.4E-45 |
| musculoskeletal | 722.6 | Degeneration of intervertebral disc | 242906 | 42226 | 0.96 (0.96,0.97) | 1.7E-37 | 0.97 (0.96,0.97) | 4.2E-33 | 0.96 (0.96,0.97) | 1.8E-40 |
| musculoskeletal | 726 | Peripheral enthesopathies and allied syndromes | 196831 | 75403 | 0.97 (0.96,0.97) | 6.3E-44 | 0.97 (0.97,0.97) | 9.6E-39 | 0.97 (0.96,0.97) | 9.3E-47 |
| musculoskeletal | 740 | Osteoarthrosis | 146222 | 131442 | 0.97 (0.97,0.98) | 8.0E-44 | 0.97 (0.97,0.98) | 3.1E-38 | 0.97 (0.97,0.97) | 4.3E-47 |
| musculoskeletal | 740.1 | Osteoarthritis; localized | 205924 | 74421 | 0.98 (0.97,0.98) | 2.4E-27 | 0.98 (0.97,0.98) | 2.1E-24 | 0.98 (0.97,0.98) | 1.6E-28 |
| musculoskeletal | 740.9 | Osteoarthrosis NOS | 179236 | 99593 | 0.97 (0.97,0.98) | 2.5E-37 | 0.97 (0.97,0.98) | 3.9E-32 | 0.97 (0.97,0.98) | 4.5E-41 |
| musculoskeletal | 745 | Pain in joint | 89094 | 187207 | 0.98 (0.97,0.98) | 7.8E-29 | 0.98 (0.97,0.98) | 4.4E-23 | 0.97 (0.97,0.98) | 2.5E-32 |
| neurological | 327.3 | Sleep apnea | 195189 | 101623 | 0.97 (0.96,0.97) | 6.9E-65 | 0.97 (0.96,0.97) | 2.0E-57 | 0.96 (0.96,0.97) | 1.9E-68 |
| neurological | 327.32 | Obstructive sleep apnea | 213928 | 82508 | 0.97 (0.96,0.97) | 2.4E-54 | 0.97 (0.96,0.97) | 3.0E-48 | 0.97 (0.96,0.97) | 6.4E-57 |
| neurological | 351 | Other peripheral nerve disorders | 227740 | 57720 | 0.97 (0.96,0.97) | 1.0E-43 | 0.97 (0.96,0.97) | 1.5E-38 | 0.96 (0.96,0.97) | 2.2E-47 |
| sense organs | 389 | Hearing loss | 121761 | 165684 | 0.97 (0.97,0.97) | 5.4E-45 | 0.97 (0.97,0.98) | 3.8E-41 | 0.97 (0.97,0.97) | 2.5E-46 |
| sense organs | 389.1 | Sensorineural hearing loss | 143787 | 132938 | 0.97 (0.96,0.97) | 7.4E-55 | 0.97 (0.96,0.97) | 6.8E-51 | 0.97 (0.96,0.97) | 1.5E-56 |
| symptoms | 760 | Back pain | 128684 | 149486 | 0.98 (0.97,0.98) | 2.0E-31 | 0.98 (0.97,0.98) | 1.8E-25 | 0.97 (0.97,0.98) | 2.5E-36 |

### Supplemental Table 10. Significant findings ($p<10^{-5}$) in PheWAS of SCZ-specific PRS in AA participants.

| Group | Phecode | Description | Cont | Case | Base OR (95% CI) | Base *p*-value | Diagnosis-adjusted OR (95% CI) | Diagnosis-adjusted *p*-value | Medication-adjusted OR (95% CI) | Medication-adjusted *p*-value |
|---|---|---|---|---|---|---|---|---|---|---|
| dermatologic | 687.4 | Disturbance of skin sensation | 62795 | 8986 | 0.98 (0.97,0.99) | 3.6E-06 | 0.98 (0.97,0.99) | 3.9E-06 | 0.98 (0.97,0.99) | 2.4E-06 |
| neurological | 351 | Other peripheral nerve disorders | 59576 | 15819 | 0.98 (0.98,0.99) | 2.5E-06 | 0.98 (0.98,0.99) | 1.9E-06 | 0.98 (0.98,0.99) | 9.1E-07 |

### Supplemental Table 11. Significant findings ($p<10^{-25}$) in PheWAS of BIP-specific PRS in EA participants.

| Group | Phecode | Description | Cont | Case | Base OR (95% CI) | Base *p*-value | Diagnosis-adjusted OR (95% CI) | Diagnosis-adjusted *p*-value | Medication-adjusted OR (95% CI) | Medication-adjusted *p*-value |
|---|---|---|---|---|---|---|---|---|---|---|
| circulatory system | 418 | Nonspecific chest pain | 178521 | 91397 | 0.98 (0.98,0.99) | 1.9E-26 | 0.99 (0.98,0.99) | 1.2E-16 | 0.98 (0.98,0.99) | 5.3E-25 |
| mental disorders | 296 | Mood disorders | 161370 | 132442 | 0.98 (0.98,0.99) | 9.6E-28 | 0.99 (0.98,1.01) | 4.6E-01 | 0.98 (0.98,0.99) | 1.6E-26 |
| mental disorders | 296.2 | Depression | 167868 | 124718 | 0.98 (0.98,0.98) | 2.6E-35 | 0.98 (0.97,0.99) | 7.9E-07 | 0.98 (0.98,0.98) | 2.9E-33 |
| mental disorders | 296.22 | Major depressive disorder | 200712 | 92217 | 0.98 (0.98,0.99) | 4.6E-26 | 0.99 (0.99,1) | 3.4E-04 | 0.98 (0.98,0.99) | 9.0E-24 |
| mental disorders | 300 | Anxiety disorders | 166912 | 123000 | 0.98 (0.98,0.98) | 2.7E-36 | 0.98 (0.98,0.99) | 1.2E-14 | 0.98 (0.98,0.98) | 1.3E-34 |
| symptoms | 785 | Abdominal pain | 206085 | 63494 | 0.98 (0.98,0.98) | 1.3E-26 | 0.98 (0.98,0.99) | 2.8E-18 | 0.98 (0.98,0.98) | 1.0E-25 |

**Supplemental Table 12. Significant findings ($p<10^{-25}$) in PheWAS of DEP-specific PRS in EA participants.**

| | | | | | Base | | Diagnosis-adjusted | | Medication-adjusted | |
|---|---|---|---|---|---|---|---|---|---|---|
| Group | Phecode | Description | Cont | Case | OR (95% CI) | *p*-value | OR (95% CI) | *p*-value | OR (95% CI) | *p*-value |
| circulatory system | 401 | Hypertension | 69818 | 225389 | 1.04 (1.04,1.05) | 1.5E-31 | 1.04 (1.03,1.04) | 4.6E-22 | 1.03 (1.02,1.04) | 7.7E-18 |
| circulatory system | 401.1 | Essential hypertension | 70815 | 224179 | 1.04 (1.04,1.05) | 1.7E-31 | 1.04 (1.03,1.04) | 4.3E-22 | 1.03 (1.02,1.04) | 7.2E-18 |
| circulatory system | 411 | Ischemic Heart Disease | 187667 | 100858 | 1.06 (1.05,1.06) | 3.4E-64 | 1.05 (1.04,1.06) | 3.2E-50 | 1.05 (1.04,1.05) | 3.5E-45 |
| circulatory system | 411.3 | Angina pectoris | 265965 | 26054 | 1.06 (1.05,1.07) | 4.2E-31 | 1.05 (1.04,1.07) | 1.1E-23 | 1.05 (1.04,1.06) | 2.2E-21 |
| circulatory system | 411.4 | Coronary atherosclerosis | 205059 | 88022 | 1.05 (1.05,1.06) | 5.0E-56 | 1.05 (1.04,1.06) | 1.6E-44 | 1.05 (1.04,1.05) | 3.9E-40 |
| circulatory system | 411.8 | Other chronic ischemic heart disease, unspecified | 242159 | 53680 | 1.05 (1.04,1.06) | 1.0E-37 | 1.05 (1.04,1.05) | 3.2E-30 | 1.04 (1.04,1.05) | 5.7E-27 |
| circulatory system | 418 | Nonspecific chest pain | 178521 | 91397 | 1.05 (1.04,1.06) | 6.7E-52 | 1.04 (1.03,1.05) | 1.3E-30 | 1.03 (1.03,1.04) | 3.6E-24 |
| digestive | 530 | Diseases of esophagus | 153615 | 131627 | 1.06 (1.05,1.07) | 4.8E-83 | 1.05 (1.05,1.06) | 9.5E-60 | 1.05 (1.04,1.05) | 2.5E-51 |
| digestive | 530.1 | Esophagitis, GERD and related diseases | 156612 | 128556 | 1.06 (1.05,1.07) | 7.5E-86 | 1.05 (1.05,1.06) | 2.6E-62 | 1.05 (1.04,1.06) | 1.1E-53 |
| digestive | 530.11 | GERD | 162469 | 122039 | 1.06 (1.05,1.07) | 3.0E-85 | 1.05 (1.05,1.06) | 3.7E-62 | 1.05 (1.04,1.06) | 1.2E-53 |
| digestive | 561 | Symptoms involving digestive system | 233209 | 41830 | 1.05 (1.04,1.06) | 1.2E-28 | 1.04 (1.03,1.04) | 5.9E-16 | 1.03 (1.02,1.04) | 2.4E-13 |
| endocrine/metabolic | 250 | Diabetes mellitus | 189084 | 108545 | 1.04 (1.04,1.05) | 3.3E-42 | 1.04 (1.03,1.04) | 6.7E-33 | 1.03 (1.03,1.04) | 9.8E-29 |
| endocrine/metabolic | 250.2 | Type 2 diabetes | 190540 | 107822 | 1.04 (1.04,1.05) | 5.8E-42 | 1.04 (1.03,1.04) | 9.4E-33 | 1.03 (1.03,1.04) | 1.1E-28 |
| endocrine/metabolic | 250.24 | Type 2 diabetes with neurological manifestations | 259077 | 40530 | 1.06 (1.05,1.07) | 2.4E-40 | 1.05 (1.04,1.06) | 4.3E-30 | 1.05 (1.04,1.05) | 4.1E-25 |
| endocrine/metabolic | 272 | Disorders of lipoid metabolism | 60613 | 234106 | 1.04 (1.03,1.05) | 1.1E-27 | 1.04 (1.03,1.04) | 1.9E-20 | 1.03 (1.02,1.04) | 4.5E-17 |
| endocrine/metabolic | 272.1 | Hyperlipidemia | 60815 | 233903 | 1.04 (1.03,1.05) | 5.8E-28 | 1.04 (1.03,1.04) | 1.1E-20 | 1.03 (1.02,1.04) | 2.9E-17 |
| endocrine/metabolic | 278 | Overweight, obesity and other hyperalimentation | 151774 | 133275 | 1.04 (1.03,1.04) | 1.1E-35 | 1.03 (1.03,1.04) | 3.8E-25 | 1.03 (1.02,1.04) | 9.3E-23 |
| endocrine/metabolic | 278.1 | Obesity | 169074 | 119822 | 1.04 (1.04,1.05) | 1.7E-41 | 1.04 (1.03,1.04) | 4.2E-30 | 1.03 (1.03,1.04) | 2.0E-27 |
| endocrine/metabolic | 278.11 | Morbid obesity | 265927 | 32713 | 1.05 (1.04,1.06) | 4.7E-27 | 1.04 (1.03,1.05) | 2.9E-20 | 1.04 (1.03,1.05) | 7.6E-19 |
| injuries & poisonings | 840 | Sprains and strains | 238860 | 36387 | 1.05 (1.04,1.06) | 4.0E-28 | 1.04 (1.03,1.05) | 1.9E-19 | 1.04 (1.03,1.05) | 2.5E-16 |
| mental disorders | 296 | Mood disorders | 161370 | 132442 | 1.06 (1.05,1.06) | 1.8E-73 | 1.14 (1.1,1.18) | 6.3E-16 | 1.02 (1.01,1.03) | 2.2E-08 |
| mental disorders | 296.2 | Depression | 167868 | 124718 | 1.07 (1.06,1.07) | 6.5E-96 | 1.12 (1.1,1.14) | 5.8E-38 | 1.04 (1.03,1.05) | 1.5E-19 |
| mental disorders | 296.22 | Major depressive disorder | 200712 | 92217 | 1.07 (1.06,1.08) | 4.6E-90 | 1.06 (1.05,1.07) | 1.4E-29 | 1.04 (1.03,1.05) | 3.2E-22 |
| mental disorders | 300 | Anxiety disorders | 166912 | 123000 | 1.05 (1.04,1.05) | 5.9E-47 | 1.02 (1.02,1.03) | 3.1E-09 | 1.01 (1.01,1.02) | 3.0E-04 |
| mental disorders | 300.1 | Anxiety disorder | 206217 | 79142 | 1.04 (1.03,1.05) | 3.5E-27 | 1.01 (1.01,1.02) | 1.3E-04 | 1.01 (1,1.01) | 6.9E-02 |
| mental disorders | 300.9 | Posttraumatic stress disorder | 230809 | 69537 | 1.04 (1.04,1.05) | 1.6E-31 | 1.02 (1.01,1.03) | 1.6E-08 | 1.02 (1.01,1.02) | 2.8E-05 |
| musculoskeletal | 716 | Other arthropathies | 247336 | 33669 | 1.05 (1.04,1.06) | 4.3E-28 | 1.04 (1.03,1.05) | 4.6E-20 | 1.04 (1.03,1.05) | 6.3E-17 |
| musculoskeletal | 716.9 | Arthropathy NOS | 258687 | 26329 | 1.06 (1.05,1.07) | 6.8E-28 | 1.05 (1.04,1.06) | 3.5E-20 | 1.05 (1.03,1.06) | 2.7E-17 |
| musculoskeletal | 721 | Spondylosis and allied disorders | 238955 | 44356 | 1.06 (1.05,1.07) | 4.5E-46 | 1.05 (1.04,1.06) | 4.0E-32 | 1.04 (1.04,1.05) | 1.8E-25 |
| musculoskeletal | 721.1 | Spondylosis without myelopathy | 242860 | 40805 | 1.06 (1.05,1.07) | 8.2E-45 | 1.05 (1.04,1.06) | 2.4E-31 | 1.05 (1.04,1.06) | 4.7E-25 |
| musculoskeletal | 722 | Intervertebral disc disorders | 228596 | 55337 | 1.06 (1.05,1.07) | 4.7E-56 | 1.05 (1.04,1.06) | 1.7E-40 | 1.05 (1.04,1.05) | 4.3E-32 |
| musculoskeletal | 722.6 | Degeneration of intervertebral disc | 242906 | 42226 | 1.06 (1.05,1.07) | 8.3E-44 | 1.05 (1.04,1.06) | 8.4E-32 | 1.05 (1.04,1.05) | 1.9E-25 |
| musculoskeletal | 726 | Peripheral enthesopathies and allied syndromes | 196831 | 75403 | 1.06 (1.05,1.06) | 9.4E-60 | 1.05 (1.04,1.06) | 1.1E-44 | 1.05 (1.04,1.05) | 1.0E-37 |
| musculoskeletal | 727 | Other disorders of synovium, tendon, and bursa | 245069 | 38207 | 1.06 (1.05,1.06) | 1.2E-34 | 1.05 (1.04,1.06) | 2.4E-26 | 1.04 (1.04,1.05) | 1.4E-22 |
| musculoskeletal | 740 | Osteoarthrosis | 146222 | 131442 | 1.06 (1.05,1.07) | 1.4E-82 | 1.05 (1.05,1.06) | 2.5E-62 | 1.05 (1.04,1.06) | 4.4E-53 |
| musculoskeletal | 740.1 | Osteoarthritis; localized | 205924 | 74421 | 1.05 (1.04,1.05) | 2.2E-42 | 1.04 (1.03,1.05) | 8.0E-33 | 1.04 (1.03,1.05) | 3.3E-28 |
| musculoskeletal | 740.11 | Osteoarthrosis, localized, primary | 238007 | 48589 | 1.05 (1.04,1.06) | 9.2E-33 | 1.04 (1.03,1.05) | 2.9E-26 | 1.04 (1.03,1.05) | 5.1E-23 |
| musculoskeletal | 740.9 | Osteoarthrosis NOS | 179236 | 99593 | 1.06 (1.05,1.06) | 4.6E-67 | 1.05 (1.04,1.06) | 4.6E-49 | 1.04 (1.04,1.05) | 4.0E-41 |
| musculoskeletal | 745 | Pain in joint | 89094 | 187207 | 1.05 (1.05,1.06) | 3.6E-55 | 1.04 (1.04,1.05) | 2.7E-36 | 1.04 (1.03,1.05) | 1.0E-28 |

| | | | | | | | | | | |
|---|---|---|---|---|---|---|---|---|---|---|
| neurological | 327 | Sleep disorders | 191386 | 81129 | 1.05 (1.04,1.06) | 7.1E-48 | 1.04 (1.03,1.04) | 2.2E-22 | 1.03 (1.02,1.04) | 9.7E-16 |
| neurological | 327.3 | Sleep apnea | 195189 | 101623 | 1.06 (1.05,1.06) | 4.6E-69 | 1.05 (1.04,1.05) | 1.7E-47 | 1.04 (1.04,1.05) | 1.2E-41 |
| neurological | 327.32 | Obstructive sleep apnea | 213928 | 82508 | 1.05 (1.05,1.06) | 3.0E-54 | 1.04 (1.04,1.05) | 2.2E-37 | 1.04 (1.03,1.05) | 4.7E-32 |
| neurological | 338 | Pain | 225658 | 47039 | 1.06 (1.05,1.07) | 2.6E-47 | 1.05 (1.04,1.06) | 1.5E-28 | 1.04 (1.03,1.05) | 8.6E-22 |
| neurological | 338.2 | Chronic pain | 248178 | 34632 | 1.06 (1.05,1.07) | 2.8E-40 | 1.05 (1.04,1.06) | 3.2E-24 | 1.04 (1.03,1.05) | 1.6E-18 |
| neurological | 339 | Other headache syndromes | 237942 | 42426 | 1.07 (1.06,1.08) | 2.2E-50 | 1.05 (1.05,1.06) | 6.7E-33 | 1.05 (1.04,1.06) | 7.0E-26 |
| neurological | 340 | Migraine | 280876 | 21151 | 1.08 (1.07,1.09) | 4.9E-36 | 1.07 (1.05,1.08) | 3.2E-27 | 1.06 (1.05,1.07) | 2.0E-22 |
| neurological | 350 | Abnormal movement | 220347 | 57430 | 1.04 (1.04,1.05) | 1.9E-29 | 1.03 (1.02,1.04) | 2.7E-14 | 1.03 (1.02,1.03) | 1.6E-11 |
| neurological | 351 | Other peripheral nerve disorders | 227740 | 57720 | 1.07 (1.06,1.07) | 2.5E-65 | 1.06 (1.05,1.06) | 1.7E-48 | 1.05 (1.04,1.06) | 6.5E-39 |
| neurological | 355.1 | Chronic pain syndrome | 283361 | 17187 | 1.08 (1.06,1.09) | 2.3E-30 | 1.06 (1.05,1.07) | 9.4E-20 | 1.05 (1.04,1.07) | 1.6E-15 |
| respiratory | 496 | Chronic airway obstruction | 215720 | 72353 | 1.06 (1.05,1.07) | 1.6E-60 | 1.05 (1.04,1.06) | 2.5E-43 | 1.05 (1.04,1.05) | 2.5E-38 |
| respiratory | 496.2 | Chronic bronchitis | 276924 | 19677 | 1.07 (1.05,1.08) | 1.3E-26 | 1.05 (1.04,1.07) | 1.0E-18 | 1.05 (1.04,1.06) | 8.1E-17 |
| respiratory | 512 | Other symptoms of respiratory system | 139662 | 120187 | 1.05 (1.05,1.06) | 1.2E-58 | 1.04 (1.03,1.05) | 4.8E-36 | 1.04 (1.03,1.04) | 1.0E-28 |
| respiratory | 512.7 | Shortness of breath | 209689 | 60907 | 1.05 (1.05,1.06) | 4.6E-44 | 1.04 (1.03,1.05) | 2.1E-28 | 1.04 (1.03,1.05) | 6.6E-24 |
| respiratory | 512.9 | Other dyspnea | 219490 | 48646 | 1.05 (1.05,1.06) | 1.5E-39 | 1.04 (1.04,1.05) | 3.0E-27 | 1.04 (1.03,1.05) | 4.7E-23 |
| sense organs | 386.9 | Dizziness and giddiness (Light-headedness and vertigo) | 231860 | 43001 | 1.05 (1.05,1.06) | 7.0E-36 | 1.04 (1.03,1.05) | 1.7E-22 | 1.04 (1.03,1.05) | 1.7E-19 |
| sense organs | 389 | Hearing loss | 121761 | 165684 | 1.04 (1.04,1.05) | 1.5E-40 | 1.04 (1.03,1.04) | 1.2E-31 | 1.04 (1.03,1.04) | 5.5E-29 |
| sense organs | 389.1 | Sensorineural hearing loss | 143787 | 132938 | 1.05 (1.04,1.06) | 1.7E-48 | 1.04 (1.04,1.05) | 2.5E-39 | 1.04 (1.04,1.05) | 4.4E-36 |
| symptoms | 760 | Back pain | 128684 | 149486 | 1.06 (1.05,1.07) | 5.6E-81 | 1.05 (1.04,1.06) | 3.3E-54 | 1.04 (1.04,1.05) | 2.4E-42 |
| symptoms | 761 | Cervicalgia | 226921 | 56206 | 1.06 (1.05,1.06) | 1.2E-46 | 1.05 (1.04,1.05) | 8.6E-31 | 1.04 (1.03,1.05) | 2.1E-24 |
| symptoms | 763 | Thoracic or lumbosacral neuritis or radiculitis, unspecified | 263827 | 29935 | 1.06 (1.05,1.07) | 1.1E-31 | 1.05 (1.04,1.06) | 2.9E-22 | 1.04 (1.03,1.05) | 7.6E-17 |
| symptoms | 765 | Cervical radiculitis | 282740 | 16245 | 1.07 (1.06,1.08) | 8.7E-26 | 1.06 (1.05,1.07) | 1.8E-19 | 1.05 (1.04,1.07) | 7.4E-16 |
| symptoms | 770 | Myalgia and myositis unspecified | 273265 | 19134 | 1.07 (1.06,1.08) | 7.6E-29 | 1.06 (1.04,1.07) | 1.1E-19 | 1.05 (1.04,1.06) | 4.9E-16 |
| symptoms | 785 | Abdominal pain | 206085 | 63494 | 1.05 (1.05,1.06) | 2.2E-45 | 1.04 (1.03,1.05) | 5.5E-28 | 1.04 (1.03,1.05) | 1.9E-22 |

**Supplemental Table 13. Significant findings ($p<10^{-5}$) in PheWAS of DEP-specific PRS in AA participants.**

| | | | | | Base | | Diagnosis-adjusted | | Medication-adjusted | |
|---|---|---|---|---|---|---|---|---|---|---|
| Group | Phecode | Description | Cont | Case | OR (95% CI) | *p*-value | OR (95% CI) | *p*-value | OR (95% CI) | *p*-value |
| neurological | 339 | Other headache syndromes | 54393 | 17396 | 1.04 (1.03,1.05) | 9.2E-09 | 1.04 (1.02,1.05) | 1.8E-08 | 1.04 (1.03,1.05) | 1.1E-08 |

### Supplemental Table 14. Significant findings ($p<10^{-25}$) in PheWAS of common factor PRS in EA participants.

| Group | Phecode | Description | Cont | Case | Base | | Diagnosis-adjusted | | Medication-adjusted | |
|---|---|---|---|---|---|---|---|---|---|---|
| | | | | | OR (95% CI) | *p*-value | OR (95% CI) | *p*-value | OR (95% CI) | *p*-value |
| circulatory system | 401 | Hypertension | 69818 | 225389 | 1.03 (1.02,1.03) | 2.2E-31 | 1.01 (1.01,1.02) | 2.1E-07 | 1.02 (1.01,1.02) | 2.2E-15 |
| circulatory system | 401.1 | Essential hypertension | 70815 | 224179 | 1.03 (1.02,1.03) | 2.6E-30 | 1.01 (1.01,1.02) | 4.6E-07 | 1.02 (1.01,1.02) | 7.6E-15 |
| circulatory system | 411 | Ischemic Heart Disease | 187667 | 100858 | 1.03 (1.03,1.04) | 5.4E-51 | 1.02 (1.01,1.02) | 4.8E-19 | 1.02 (1.02,1.03) | 1.3E-30 |
| circulatory system | 411.1 | Unstable angina | 287650 | 11889 | 1.06 (1.05,1.07) | 8.0E-32 | 1.04 (1.03,1.05) | 1.2E-14 | 1.04 (1.04,1.05) | 5.3E-21 |
| circulatory system | 411.3 | Angina pectoris | 265965 | 26054 | 1.04 (1.03,1.05) | 1.6E-34 | 1.02 (1.02,1.03) | 7.8E-13 | 1.03 (1.03,1.04) | 5.4E-22 |
| circulatory system | 411.4 | Coronary atherosclerosis | 205059 | 88022 | 1.03 (1.02,1.03) | 3.3E-33 | 1.01 (1.01,1.02) | 2.4E-11 | 1.02 (1.02,1.02) | 2.8E-20 |
| circulatory system | 418 | Nonspecific chest pain | 178521 | 91397 | 1.06 (1.05,1.06) | 3.0E-174 | 1.04 (1.03,1.04) | 3.0E-64 | 1.04 (1.04,1.05) | 9.4E-99 |
| circulatory system | 427 | Cardiac dysrhythmias | 168977 | 104053 | 1.03 (1.02,1.03) | 1.9E-39 | 1.01 (1.01,1.02) | 5.8E-10 | 1.02 (1.01,1.02) | 5.0E-16 |
| circulatory system | 428 | Congestive heart failure; nonhypertensive | 254758 | 41526 | 1.03 (1.02,1.04) | 2.6E-28 | 1.01 (1.01,1.02) | 7.8E-08 | 1.02 (1.01,1.03) | 3.2E-13 |
| circulatory system | 433 | Cerebrovascular disease | 239598 | 49889 | 1.03 (1.03,1.04) | 6.2E-34 | 1.01 (1.01,1.02) | 3.1E-08 | 1.02 (1.01,1.03) | 1.7E-15 |
| circulatory system | 455 | Hemorrhoids | 222924 | 42032 | 1.03 (1.02,1.03) | 1.9E-27 | 1.02 (1.01,1.02) | 3.4E-10 | 1.02 (1.02,1.03) | 2.3E-15 |
| circulatory system | 458 | Hypotension | 256763 | 27983 | 1.04 (1.03,1.05) | 7.7E-35 | 1.01 (1.01,1.02) | 3.2E-05 | 1.02 (1.01,1.02) | 3.0E-08 |
| dermatologic | 681 | Superficial cellulitis and abscess | 224657 | 52705 | 1.04 (1.03,1.04) | 1.7E-54 | 1.02 (1.01,1.02) | 1.8E-14 | 1.02 (1.02,1.03) | 2.2E-20 |
| dermatologic | 687.1 | Rash and other nonspecific skin eruption | 236181 | 30351 | 1.04 (1.03,1.05) | 2.3E-37 | 1.02 (1.02,1.03) | 8.9E-14 | 1.03 (1.02,1.03) | 1.5E-19 |
| digestive | 521 | Diseases of hard tissues of teeth | 233809 | 60055 | 1.05 (1.05,1.06) | 3.6E-118 | 1.03 (1.02,1.03) | 1.0E-30 | 1.03 (1.03,1.04) | 3.8E-43 |
| digestive | 521.1 | Dental caries | 236323 | 58432 | 1.05 (1.05,1.06) | 3.6E-116 | 1.03 (1.02,1.03) | 2.3E-30 | 1.03 (1.03,1.04) | 2.1E-42 |
| digestive | 522 | Diseases of pulp and periapical tissues | 284618 | 14361 | 1.06 (1.05,1.07) | 1.1E-45 | 1.04 (1.03,1.04) | 9.5E-16 | 1.04 (1.03,1.05) | 2.3E-18 |
| digestive | 522.1 | Pulpitis and necrosis of tooth pulp | 294641 | 8437 | 1.07 (1.06,1.08) | 3.7E-37 | 1.04 (1.03,1.06) | 5.1E-15 | 1.05 (1.04,1.06) | 1.1E-16 |
| digestive | 523 | Gingival and periodontal diseases | 246246 | 49998 | 1.05 (1.05,1.06) | 4.4E-95 | 1.03 (1.02,1.03) | 5.1E-24 | 1.03 (1.03,1.04) | 1.3E-33 |
| digestive | 523.1 | Gingivitis | 281815 | 17735 | 1.05 (1.05,1.06) | 5.9E-41 | 1.03 (1.02,1.04) | 4.4E-12 | 1.03 (1.02,1.04) | 3.0E-16 |
| digestive | 523.3 | Periodontitis (acute or chronic) | 271618 | 25654 | 1.06 (1.05,1.06) | 1.5E-64 | 1.03 (1.02,1.04) | 5.2E-19 | 1.03 (1.03,1.04) | 3.2E-25 |
| digestive | 523.31 | Acute periodontitis | 282062 | 17450 | 1.05 (1.05,1.06) | 3.9E-41 | 1.03 (1.02,1.03) | 3.6E-11 | 1.03 (1.02,1.04) | 3.5E-15 |
| digestive | 523.32 | Chronic periodontitis | 287968 | 14089 | 1.06 (1.05,1.06) | 5.4E-36 | 1.03 (1.02,1.04) | 2.6E-11 | 1.03 (1.03,1.04) | 3.5E-15 |
| digestive | 525 | Other diseases of the teeth and supporting structures | 230287 | 60680 | 1.06 (1.05,1.06) | 3.4E-135 | 1.03 (1.03,1.03) | 6.7E-36 | 1.04 (1.03,1.04) | 3.7E-48 |
| digestive | 525.1 | Loss of teeth or edentulism | 261829 | 38073 | 1.05 (1.05,1.06) | 7.4E-85 | 1.03 (1.02,1.03) | 4.1E-20 | 1.03 (1.03,1.04) | 1.0E-27 |
| digestive | 530 | Diseases of esophagus | 153615 | 131627 | 1.04 (1.04,1.04) | 2.3E-102 | 1.02 (1.02,1.03) | 3.2E-31 | 1.03 (1.03,1.03) | 4.7E-54 |
| digestive | 530.1 | Esophagitis, GERD and related diseases | 156612 | 128556 | 1.04 (1.04,1.05) | 3.8E-102 | 1.02 (1.02,1.03) | 1.6E-31 | 1.03 (1.03,1.03) | 4.4E-54 |
| digestive | 530.11 | GERD | 162469 | 122039 | 1.04 (1.04,1.05) | 5.1E-101 | 1.02 (1.02,1.03) | 1.6E-31 | 1.03 (1.03,1.03) | 1.9E-53 |
| digestive | 532 | Dysphagia | 252740 | 40103 | 1.04 (1.03,1.04) | 1.4E-44 | 1.02 (1.01,1.02) | 3.4E-10 | 1.02 (1.01,1.03) | 3.3E-13 |
| digestive | 550 | Abdominal hernia | 237547 | 50050 | 1.03 (1.02,1.03) | 6.0E-29 | 1.02 (1.01,1.02) | 2.3E-10 | 1.02 (1.01,1.02) | 6.1E-16 |
| digestive | 561 | Symptoms involving digestive system | 233209 | 41830 | 1.05 (1.04,1.05) | 4.7E-66 | 1.02 (1.02,1.03) | 4.7E-16 | 1.03 (1.02,1.03) | 9.3E-26 |
| digestive | 578 | Gastrointestinal hemorrhage | 250313 | 35111 | 1.04 (1.03,1.04) | 9.4E-40 | 1.02 (1.01,1.03) | 4.4E-13 | 1.02 (1.02,1.03) | 5.6E-17 |
| endocrine/metabolic | 276 | Disorders of fluid, electrolyte, and acid-base balance | 221908 | 54151 | 1.04 (1.03,1.04) | 2.1E-59 | 1.02 (1.01,1.02) | 1.2E-13 | 1.02 (1.01,1.02) | 9.6E-15 |
| endocrine/metabolic | 276.1 | Electrolyte imbalance | 242364 | 39794 | 1.03 (1.03,1.04) | 7.2E-33 | 1.01 (1.01,1.02) | 2.4E-07 | 1.01 (1.01,1.02) | 3.6E-07 |
| endocrine/metabolic | 276.5 | Hypovolemia | 270652 | 16482 | 1.05 (1.04,1.05) | 1.2E-29 | 1.02 (1.01,1.03) | 1.5E-06 | 1.02 (1.01,1.03) | 4.6E-07 |
| genitourinary | 591 | Urinary tract infection | 252052 | 33029 | 1.04 (1.04,1.05) | 2.2E-42 | 1.02 (1.02,1.03) | 1.2E-12 | 1.02 (1.02,1.03) | 4.0E-16 |
| genitourinary | 599 | Other symptoms of urinary system | 215307 | 59968 | 1.04 (1.04,1.04) | 1.6E-63 | 1.02 (1.02,1.03) | 1.7E-17 | 1.03 (1.02,1.03) | 3.6E-32 |
| genitourinary | 600 | Hyperplasia of prostate | 189050 | 97473 | 1.03 (1.03,1.04) | 8.4E-48 | 1.02 (1.01,1.02) | 1.6E-16 | 1.02 (1.02,1.03) | 7.6E-28 |
| genitourinary | 601 | Inflammatory diseases of prostate | 281612 | 14536 | 1.05 (1.04,1.06) | 1.6E-31 | 1.03 (1.03,1.04) | 1.2E-15 | 1.04 (1.03,1.05) | 1.9E-20 |
| genitourinary | 605 | Erectile dysfunction [ED] | 209855 | 76830 | 1.02 (1.02,1.03) | 5.0E-30 | 1.01 (1.01,1.02) | 1.6E-07 | 1.02 (1.01,1.02) | 2.6E-18 |
| infectious diseases | 70.3 | Viral hepatitis C | 294978 | 13935 | 1.07 (1.06,1.08) | 1.8E-49 | 1.05 (1.04,1.05) | 3.7E-24 | 1.05 (1.04,1.05) | 4.1E-24 |
| infectious diseases | 110 | Dermatophytosis / Dermatomycosis | 206920 | 69951 | 1.03 (1.02,1.03) | 2.9E-37 | 1.01 (1.01,1.02) | 8.0E-08 | 1.02 (1.01,1.02) | 2.9E-16 |
| infectious diseases | 110.1 | Dermatophytosis | 209524 | 68178 | 1.03 (1.02,1.03) | 3.1E-36 | 1.01 (1.01,1.02) | 1.7E-07 | 1.02 (1.01,1.02) | 8.9E-16 |

| | | | | | | | | | | |
|---|---|---|---|---|---|---|---|---|---|---|
| infectious diseases | 110.11 | Dermatophytosis of nail | 235188 | 52596 | 1.03 (1.02,1.03) | 2.4E-26 | 1.01 (1,1.01) | 3.4E-05 | 1.02 (1.01,1.02) | 1.8E-11 |
| injuries & poisonings | 916 | Contusion | 261799 | 18348 | 1.06 (1.05,1.07) | 7.5E-51 | 1.03 (1.02,1.04) | 7.8E-17 | 1.04 (1.03,1.05) | 2.5E-21 |
| injuries & poisonings | 969 | Poisoning by psychotropic agents | 306220 | 1944 | 1.14 (1.11,1.17) | 4.0E-30 | 1.1 (1.07,1.12) | 4.0E-16 | 1.07 (1.05,1.1) | 1.5E-09 |
| mental disorders | 290.1 | Dementias | 291550 | 14058 | 1.05 (1.04,1.06) | 1.2E-29 | 1.02 (1.01,1.03) | 2.4E-05 | 1.02 (1.01,1.03) | 4.5E-07 |
| mental disorders | 290.3 | Other persistent mental disorders due to conditions classified elsewhere | 288225 | 14474 | 1.06 (1.05,1.06) | 1.7E-37 | 1.02 (1.01,1.03) | 2.6E-05 | 1.02 (1.01,1.03) | 9.6E-08 |
| mental disorders | 292 | Neurological disorders | 246745 | 40721 | 1.05 (1.05,1.06) | 9.6E-88 | 1.02 (1.02,1.03) | 1.8E-16 | 1.03 (1.02,1.03) | 6.5E-24 |
| mental disorders | 292.4 | Altered mental status | 288556 | 10297 | 1.07 (1.06,1.08) | 6.4E-44 | 1.04 (1.03,1.05) | 2.4E-13 | 1.03 (1.02,1.04) | 8.5E-09 |
| mental disorders | 296 | Mood disorders | 161370 | 132442 | 1.11 (1.1,1.11) | 0.0E+00 | 1.04 (1.02,1.06) | 1.1E-04 | 1.09 (1.08,1.09) | 0.0E+00 |
| mental disorders | 296.1 | Bipolar | 285708 | 20399 | 1.16 (1.15,1.17) | 0.0E+00 | 1.11 (1.1,1.12) | 3.2E-166 | 1.1 (1.09,1.11) | 5.6E-121 |
| mental disorders | 296.2 | Depression | 167868 | 124718 | 1.1 (1.09,1.1) | 0.0E+00 | 1 (0.99,1.01) | 5.6E-01 | 1.08 (1.07,1.08) | 1.5E-290 |
| mental disorders | 296.22 | Major depressive disorder | 200712 | 92217 | 1.09 (1.09,1.1) | 0.0E+00 | 1.02 (1.02,1.03) | 4.1E-16 | 1.07 (1.06,1.07) | 1.2E-196 |
| mental disorders | 297 | Suicidal ideation or attempt | 285894 | 14832 | 1.13 (1.12,1.14) | 7.6E-166 | 1.08 (1.07,1.09) | 2.4E-73 | 1.07 (1.06,1.08) | 5.0E-53 |
| mental disorders | 297.1 | Suicidal ideation | 291719 | 12317 | 1.13 (1.12,1.14) | 1.6E-149 | 1.09 (1.08,1.1) | 3.5E-67 | 1.07 (1.06,1.09) | 6.2E-47 |
| mental disorders | 297.2 | Suicide or self-inflicted injury | 304207 | 3948 | 1.12 (1.11,1.14) | 1.3E-47 | 1.08 (1.07,1.1) | 2.1E-22 | 1.06 (1.04,1.08) | 2.3E-12 |
| mental disorders | 300 | Anxiety disorders | 166912 | 123000 | 1.09 (1.09,1.1) | 0.0E+00 | 1.05 (1.04,1.05) | 7.2E-80 | 1.07 (1.07,1.08) | 4.7E-236 |
| mental disorders | 300.1 | Anxiety disorder | 206217 | 79142 | 1.09 (1.09,1.1) | 0.0E+00 | 1.05 (1.05,1.06) | 1.7E-102 | 1.07 (1.07,1.08) | 2.0E-201 |
| mental disorders | 300.11 | Generalized anxiety disorder | 278052 | 21853 | 1.08 (1.07,1.09) | 4.1E-99 | 1.04 (1.03,1.05) | 3.4E-26 | 1.05 (1.04,1.06) | 1.1E-43 |
| mental disorders | 300.12 | Agorophobia, social phobia, & panic dis | 292966 | 12188 | 1.08 (1.07,1.09) | 2.1E-67 | 1.05 (1.04,1.06) | 8.3E-23 | 1.05 (1.04,1.06) | 2.5E-26 |
| mental disorders | 300.3 | Obsessive-compulsive disorders | 306240 | 3142 | 1.11 (1.09,1.13) | 2.2E-32 | 1.07 (1.05,1.09) | 9.2E-15 | 1.07 (1.05,1.09) | 5.4E-14 |
| mental disorders | 300.4 | Dysthymic disorder | 274401 | 23556 | 1.08 (1.07,1.09) | 2.9E-117 | 1.03 (1.03,1.04) | 6.8E-22 | 1.05 (1.05,1.06) | 5.3E-51 |
| mental disorders | 300.9 | Posttraumatic stress disorder | 230809 | 69537 | 1.06 (1.06,1.07) | 1.4E-166 | 1.02 (1.02,1.03) | 6.9E-18 | 1.04 (1.03,1.04) | 6.3E-55 |
| mental disorders | 301 | Personality disorders | 290676 | 13909 | 1.14 (1.12,1.15) | 2.2E-177 | 1.09 (1.08,1.1) | 8.0E-85 | 1.08 (1.07,1.09) | 1.6E-65 |
| mental disorders | 301.2 | Antisocial/borderline personality disorder | 303440 | 5191 | 1.15 (1.14,1.17) | 9.3E-85 | 1.11 (1.09,1.13) | 1.5E-46 | 1.09 (1.07,1.1) | 3.1E-29 |
| mental disorders | 303 | Psychogenic and somatoform disorders | 297603 | 7203 | 1.08 (1.06,1.09) | 1.8E-34 | 1.04 (1.03,1.05) | 2.1E-10 | 1.05 (1.03,1.06) | 1.3E-13 |
| mental disorders | 303.4 | Somatoform disorder | 300051 | 6434 | 1.08 (1.06,1.09) | 2.2E-31 | 1.04 (1.03,1.05) | 1.7E-09 | 1.05 (1.03,1.06) | 6.7E-13 |
| mental disorders | 304 | Adjustment reaction | 238220 | 47151 | 1.06 (1.05,1.06) | 2.5E-108 | 1.02 (1.01,1.03) | 1.6E-13 | 1.04 (1.04,1.05) | 5.7E-55 |
| mental disorders | 306 | Other mental disorder | 257665 | 23904 | 1.07 (1.06,1.08) | 2.2E-89 | 1.03 (1.03,1.04) | 9.8E-23 | 1.04 (1.04,1.05) | 5.2E-33 |
| mental disorders | 316 | Substance addiction and disorders | 261155 | 36576 | 1.1 (1.09,1.1) | 2.2E-215 | 1.06 (1.05,1.07) | 1.6E-82 | 1.06 (1.06,1.07) | 6.2E-83 |
| mental disorders | 317.1 | Alcoholism | 255939 | 39754 | 1.07 (1.06,1.07) | 1.5E-125 | 1.04 (1.03,1.05) | 1.7E-42 | 1.04 (1.04,1.05) | 1.3E-48 |
| mental disorders | 318 | Tobacco use disorder | 175890 | 103781 | 1.05 (1.05,1.06) | 3.5E-141 | 1.03 (1.03,1.04) | 1.4E-59 | 1.04 (1.03,1.04) | 2.8E-72 |
| musculoskeletal | 716 | Other arthropathies | 247336 | 33669 | 1.03 (1.03,1.04) | 3.7E-27 | 1.02 (1.01,1.02) | 1.9E-07 | 1.02 (1.02,1.03) | 6.4E-14 |
| musculoskeletal | 720 | Spinal stenosis | 265640 | 30340 | 1.04 (1.03,1.04) | 1.2E-32 | 1.02 (1.01,1.02) | 6.8E-09 | 1.03 (1.02,1.03) | 1.0E-18 |
| musculoskeletal | 721 | Spondylosis and allied disorders | 238955 | 44356 | 1.04 (1.04,1.05) | 7.1E-55 | 1.02 (1.01,1.03) | 1.2E-14 | 1.03 (1.02,1.04) | 1.0E-30 |
| musculoskeletal | 721.1 | Spondylosis without myelopathy | 242860 | 40805 | 1.04 (1.03,1.05) | 7.7E-49 | 1.02 (1.01,1.02) | 1.3E-12 | 1.03 (1.02,1.03) | 7.4E-27 |
| musculoskeletal | 722 | Intervertebral disc disorders | 228596 | 55337 | 1.04 (1.04,1.04) | 7.4E-62 | 1.02 (1.02,1.03) | 3.5E-17 | 1.03 (1.02,1.03) | 6.3E-34 |
| musculoskeletal | 722.6 | Degeneration of intervertebral disc | 242906 | 42226 | 1.04 (1.03,1.04) | 1.6E-48 | 1.02 (1.01,1.03) | 5.1E-14 | 1.03 (1.02,1.03) | 1.1E-26 |
| musculoskeletal | 740 | Osteoarthrosis | 146222 | 131442 | 1.03 (1.02,1.03) | 3.6E-39 | 1.01 (1,1.01) | 3.6E-06 | 1.02 (1.01,1.02) | 1.2E-19 |
| musculoskeletal | 740.9 | Osteoarthrosis NOS | 179236 | 99593 | 1.03 (1.03,1.03) | 2.9E-51 | 1.01 (1.01,1.02) | 1.5E-11 | 1.02 (1.02,1.03) | 5.5E-26 |
| musculoskeletal | 745 | Pain in joint | 89094 | 187207 | 1.03 (1.03,1.04) | 7.6E-61 | 1.01 (1.01,1.02) | 5.2E-10 | 1.02 (1.02,1.03) | 1.7E-31 |
| neurological | 327 | Sleep disorders | 191386 | 81129 | 1.05 (1.05,1.05) | 2.0E-116 | 1.02 (1.01,1.02) | 1.5E-15 | 1.03 (1.03,1.04) | 8.9E-48 |
| neurological | 327.3 | Sleep apnea | 195189 | 101623 | 1.02 (1.02,1.03) | 1.8E-30 | 1 (1,1.01) | 1.3E-01 | 1.02 (1.01,1.02) | 1.5E-15 |
| neurological | 327.4 | Insomnia | 230277 | 51649 | 1.05 (1.05,1.06) | 1.1E-96 | 1.02 (1.02,1.03) | 6.2E-17 | 1.03 (1.03,1.04) | 1.8E-37 |
| neurological | 338 | Pain | 225658 | 47039 | 1.06 (1.05,1.06) | 8.4E-104 | 1.03 (1.02,1.03) | 3.5E-28 | 1.04 (1.03,1.04) | 1.8E-46 |
| neurological | 338.2 | Chronic pain | 248178 | 34632 | 1.06 (1.06,1.07) | 3.6E-99 | 1.03 (1.03,1.04) | 3.2E-30 | 1.04 (1.04,1.05) | 2.3E-47 |
| neurological | 339 | Other headache syndromes | 237942 | 42426 | 1.05 (1.04,1.05) | 8.1E-67 | 1.02 (1.02,1.03) | 8.6E-14 | 1.03 (1.02,1.03) | 9.2E-25 |
| neurological | 350 | Abnormal movement | 220347 | 57430 | 1.04 (1.04,1.05) | 2.1E-68 | 1.02 (1.01,1.02) | 3.9E-10 | 1.02 (1.02,1.03) | 2.6E-21 |
| neurological | 350.2 | Abnormality of gait | 238406 | 44292 | 1.04 (1.04,1.05) | 6.7E-53 | 1.01 (1.01,1.02) | 2.2E-08 | 1.02 (1.02,1.03) | 3.6E-17 |

| | | | | | | | | | |
|---|---|---|---|---|---|---|---|---|---|
| neurological | 351 | Other peripheral nerve disorders | 227740 | 57720 | 1.03 (1.02,1.03) | 2.3E-36 | 1.01 (1.01,1.02) | 6.4E-06 | 1.02 (1.02,1.02) | 1.2E-17 |
| neurological | 355.1 | Chronic pain syndrome | 283361 | 17187 | 1.07 (1.06,1.08) | 8.8E-64 | 1.04 (1.03,1.05) | 8.4E-21 | 1.05 (1.04,1.06) | 1.7E-32 |
| neurological | 356 | Hereditary and idiopathic peripheral neuropathy | 264091 | 26984 | 1.03 (1.03,1.04) | 3.8E-26 | 1.01 (1.01,1.02) | 2.2E-05 | 1.02 (1.02,1.03) | 1.8E-12 |
| respiratory | 464 | Acute sinusitis | 264174 | 17840 | 1.06 (1.06,1.07) | 1.2E-57 | 1.05 (1.04,1.05) | 3.8E-30 | 1.05 (1.05,1.06) | 4.7E-40 |
| respiratory | 465 | Acute upper respiratory infections of multiple or unspecified sites | 204993 | 53253 | 1.06 (1.05,1.06) | 9.3E-121 | 1.04 (1.03,1.04) | 3.2E-49 | 1.04 (1.04,1.05) | 4.6E-67 |
| respiratory | 465.2 | Acute pharyngitis | 276813 | 9789 | 1.06 (1.05,1.08) | 2.9E-33 | 1.05 (1.03,1.06) | 1.8E-17 | 1.05 (1.04,1.06) | 1.4E-20 |
| respiratory | 475 | Chronic sinusitis | 257693 | 26585 | 1.04 (1.04,1.05) | 4.7E-42 | 1.03 (1.02,1.03) | 1.3E-17 | 1.04 (1.03,1.04) | 1.0E-26 |
| respiratory | 476 | Allergic rhinitis | 215075 | 63297 | 1.03 (1.02,1.03) | 1.9E-36 | 1.02 (1.01,1.02) | 4.3E-14 | 1.02 (1.02,1.03) | 5.2E-25 |
| respiratory | 480 | Pneumonia | 260383 | 31337 | 1.04 (1.03,1.05) | 3.4E-39 | 1.02 (1.01,1.02) | 4.0E-10 | 1.02 (1.01,1.03) | 2.0E-11 |
| respiratory | 496 | Chronic airway obstruction | 215720 | 72353 | 1.06 (1.05,1.06) | 9.2E-137 | 1.04 (1.03,1.04) | 3.9E-62 | 1.04 (1.04,1.05) | 2.8E-79 |
| respiratory | 496.2 | Chronic bronchitis | 276924 | 19677 | 1.06 (1.05,1.07) | 4.3E-59 | 1.04 (1.03,1.05) | 2.6E-26 | 1.04 (1.04,1.05) | 3.9E-31 |
| respiratory | 496.21 | Obstructive chronic bronchitis | 282188 | 17154 | 1.06 (1.05,1.07) | 2.0E-50 | 1.04 (1.03,1.05) | 8.0E-22 | 1.04 (1.03,1.05) | 8.8E-26 |
| respiratory | 497 | Bronchitis | 268282 | 14750 | 1.07 (1.06,1.08) | 7.7E-60 | 1.05 (1.04,1.06) | 9.3E-29 | 1.05 (1.04,1.06) | 2.0E-34 |
| respiratory | 509 | Respiratory failure, insufficiency, arrest | 275454 | 22821 | 1.04 (1.03,1.05) | 1.2E-28 | 1.02 (1.01,1.03) | 2.3E-08 | 1.02 (1.01,1.03) | 1.5E-07 |
| respiratory | 512 | Other symptoms of respiratory system | 139662 | 120187 | 1.04 (1.04,1.05) | 1.6E-104 | 1.02 (1.02,1.02) | 4.8E-24 | 1.03 (1.03,1.03) | 3.0E-49 |
| respiratory | 512.7 | Shortness of breath | 209689 | 60907 | 1.04 (1.04,1.05) | 3.7E-70 | 1.02 (1.02,1.03) | 3.5E-18 | 1.03 (1.02,1.03) | 7.1E-33 |
| respiratory | 512.8 | Cough | 212486 | 48953 | 1.03 (1.03,1.04) | 2.6E-35 | 1.01 (1.01,1.02) | 4.7E-07 | 1.02 (1.01,1.02) | 1.1E-14 |
| respiratory | 512.9 | Other dyspnea | 219490 | 48646 | 1.03 (1.02,1.03) | 9.6E-32 | 1.01 (1,1.01) | 1.5E-04 | 1.02 (1.01,1.02) | 1.1E-12 |
| sense organs | 366 | Cataract | 116658 | 166416 | 1.03 (1.02,1.03) | 2.1E-37 | 1.01 (1.01,1.01) | 1.4E-06 | 1.02 (1.01,1.02) | 2.0E-18 |
| sense organs | 366.2 | Senile cataract | 139596 | 142445 | 1.02 (1.02,1.03) | 5.4E-28 | 1.01 (1,1.01) | 1.5E-04 | 1.01 (1.01,1.02) | 3.0E-13 |
| sense organs | 367 | Disorders of refraction and accommodation; blindness and low vision | 86906 | 200705 | 1.03 (1.03,1.04) | 5.2E-58 | 1.01 (1.01,1.02) | 3.5E-11 | 1.02 (1.02,1.03) | 4.4E-27 |
| sense organs | 375.1 | Dry eyes | 216725 | 58461 | 1.03 (1.02,1.03) | 9.9E-29 | 1.01 (1.01,1.02) | 3.5E-06 | 1.02 (1.01,1.02) | 1.7E-14 |
| sense organs | 386.9 | Dizziness and giddiness (Light-headedness and vertigo) | 231860 | 43001 | 1.04 (1.04,1.05) | 2.2E-56 | 1.02 (1.01,1.02) | 1.7E-12 | 1.03 (1.02,1.03) | 9.3E-27 |
| symptoms | 760 | Back pain | 128684 | 149486 | 1.05 (1.05,1.05) | 2.3E-146 | 1.03 (1.02,1.03) | 7.0E-46 | 1.04 (1.04,1.04) | 2.5E-86 |
| symptoms | 761 | Cervicalgia | 226921 | 56206 | 1.05 (1.05,1.06) | 3.9E-96 | 1.03 (1.02,1.03) | 1.9E-33 | 1.04 (1.03,1.04) | 4.2E-56 |
| symptoms | 763 | Thoracic or lumbosacral neuritis or radiculitis, unspecified | 263827 | 29935 | 1.04 (1.03,1.05) | 2.5E-38 | 1.02 (1.01,1.03) | 1.2E-11 | 1.03 (1.02,1.04) | 9.6E-22 |
| symptoms | 770 | Myalgia and myositis unspecified | 273265 | 19134 | 1.06 (1.05,1.06) | 5.4E-48 | 1.03 (1.02,1.04) | 1.3E-16 | 1.04 (1.03,1.05) | 3.2E-25 |
| symptoms | 772.3 | Muscle weakness | 265118 | 25069 | 1.04 (1.03,1.05) | 6.0E-35 | 1.02 (1.01,1.02) | 5.1E-07 | 1.02 (1.01,1.03) | 2.4E-10 |
| symptoms | 785 | Abdominal pain | 206085 | 63494 | 1.05 (1.05,1.06) | 4.1E-120 | 1.03 (1.03,1.04) | 4.0E-42 | 1.04 (1.03,1.04) | 1.7E-60 |
| symptoms | 788 | Syncope and collapse | 267138 | 25048 | 1.04 (1.03,1.04) | 7.3E-28 | 1.01 (1.01,1.02) | 1.7E-04 | 1.02 (1.01,1.03) | 2.6E-08 |
| symptoms | 789 | Nausea and vomiting | 262919 | 21357 | 1.06 (1.05,1.07) | 9.3E-56 | 1.03 (1.02,1.04) | 7.8E-18 | 1.03 (1.02,1.04) | 1.6E-18 |
| symptoms | 798 | Malaise and fatigue | 223258 | 47019 | 1.04 (1.04,1.05) | 7.8E-65 | 1.02 (1.01,1.02) | 1.7E-11 | 1.03 (1.02,1.03) | 2.2E-26 |

### Supplemental Table 15. Significant findings ($p<10^{-5}$) in PheWAS of common factor PRS in AA participants.

| Group | Phecode | Description | Cont | Case | Base | | Diagnosis-adjusted | | Medication-adjusted | |
|---|---|---|---|---|---|---|---|---|---|---|
| | | | | | OR (95% CI) | *p*-value | OR (95% CI) | *p*-value | OR (95% CI) | *p*-value |
| circulatory system | 418 | Nonspecific chest pain | 41653 | 29692 | 1.03 (1.02,1.04) | 3.5E-15 | 1.02 (1.01,1.03) | 1.4E-07 | 1.02 (1.02,1.03) | 1.4E-09 |
| dermatologic | 681 | Superficial cellulitis and abscess | 59215 | 13907 | 1.03 (1.02,1.03) | 7.4E-08 | 1.02 (1.01,1.03) | 1.1E-04 | 1.02 (1.01,1.03) | 1.0E-04 |
| digestive | 521 | Diseases of hard tissues of teeth | 48819 | 27048 | 1.03 (1.03,1.04) | 6.1E-18 | 1.02 (1.01,1.03) | 8.7E-08 | 1.02 (1.01,1.03) | 5.2E-09 |
| digestive | 521.1 | Dental caries | 49444 | 26480 | 1.03 (1.02,1.04) | 5.9E-17 | 1.02 (1.01,1.03) | 3.1E-07 | 1.02 (1.01,1.03) | 2.1E-08 |
| digestive | 523 | Gingival and periodontal diseases | 53599 | 22773 | 1.03 (1.02,1.04) | 3.9E-14 | 1.02 (1.01,1.03) | 8.6E-06 | 1.02 (1.01,1.03) | 9.4E-07 |
| digestive | 523.3 | Periodontitis (acute or chronic) | 63670 | 12964 | 1.03 (1.02,1.04) | 9.2E-09 | 1.02 (1.01,1.03) | 8.0E-04 | 1.02 (1.01,1.03) | 2.4E-04 |
| digestive | 523.31 | Acute periodontitis | 69118 | 8566 | 1.03 (1.01,1.04) | 3.8E-06 | 1.01 (1,1.03) | 8.8E-03 | 1.02 (1.01,1.03) | 3.4E-03 |
| digestive | 525 | Other diseases of the teeth | 47747 | 27265 | 1.03 (1.02,1.04) | 5.8E-17 | 1.02 (1.01,1.03) | 8.8E-07 | 1.02 (1.01,1.03) | 1.1E-07 |
| digestive | 525.1 | Loss of teeth or edentulism | 62014 | 16377 | 1.03 (1.02,1.04) | 7.4E-13 | 1.02 (1.01,1.03) | 1.2E-05 | 1.02 (1.01,1.03) | 6.1E-06 |
| digestive | 526 | Diseases of the jaws | 76125 | 2427 | 1.05 (1.03,1.07) | 1.6E-06 | 1.04 (1.02,1.06) | 1.2E-04 | 1.04 (1.02,1.06) | 6.1E-05 |
| digestive | 530 | Diseases of esophagus | 42841 | 32106 | 1.02 (1.02,1.03) | 3.5E-10 | 1.01 (1.01,1.02) | 1.1E-04 | 1.02 (1.01,1.02) | 7.1E-06 |
| digestive | 530.1 | Esophagitis, GERD and related | 43610 | 31338 | 1.02 (1.02,1.03) | 1.6E-10 | 1.01 (1.01,1.02) | 6.2E-05 | 1.02 (1.01,1.02) | 3.7E-06 |
| digestive | 530.11 | GERD | 44780 | 30147 | 1.02 (1.02,1.03) | 3.5E-10 | 1.01 (1.01,1.02) | 9.4E-05 | 1.02 (1.01,1.02) | 6.3E-06 |
| digestive | 561 | Symptoms involving digestive system | 61913 | 10387 | 1.03 (1.02,1.04) | 4.7E-09 | 1.02 (1.01,1.03) | 5.2E-05 | 1.02 (1.01,1.03) | 1.6E-05 |
| infectious diseases | 110 | Dermatophytosis / Dermatomycosis | 45593 | 27229 | 1.02 (1.02,1.03) | 1.5E-09 | 1.02 (1.01,1.02) | 3.1E-05 | 1.02 (1.01,1.03) | 4.0E-06 |
| infectious diseases | 110.1 | Dermatophytosis | 46880 | 26148 | 1.02 (1.02,1.03) | 1.6E-09 | 1.02 (1.01,1.02) | 2.3E-05 | 1.02 (1.01,1.03) | 3.9E-06 |
| infectious diseases | 110.11 | Dermatophytosis of nail | 55927 | 19673 | 1.02 (1.01,1.03) | 6.5E-06 | 1.01 (1,1.02) | 3.8E-03 | 1.01 (1.01,1.02) | 1.2E-03 |
| injuries & poisonings | 916 | Contusion | 70114 | 4461 | 1.04 (1.02,1.05) | 2.5E-06 | 1.03 (1.01,1.04) | 6.8E-04 | 1.03 (1.01,1.04) | 3.9E-04 |
| mental disorders | 296 | Mood disorders | 33552 | 44186 | 1.05 (1.04,1.06) | 3.6E-37 | 1 (0.97,1.03) | 8.8E-01 | 1.04 (1.03,1.05) | 9.6E-20 |
| mental disorders | 296.1 | Bipolar | 75000 | 5921 | 1.06 (1.05,1.08) | 2.1E-20 | 1.05 (1.03,1.06) | 4.2E-11 | 1.04 (1.02,1.05) | 5.9E-07 |
| mental disorders | 296.2 | Depression | 35260 | 42196 | 1.05 (1.04,1.05) | 2.4E-33 | 0.99 (0.98,1.01) | 5.4E-01 | 1.03 (1.03,1.04) | 5.9E-18 |
| mental disorders | 296.22 | Major depressive disorder | 44614 | 32643 | 1.04 (1.03,1.04) | 9.7E-22 | 1 (0.99,1.01) | 7.6E-01 | 1.02 (1.02,1.03) | 8.8E-10 |
| mental disorders | 297 | Suicidal ideation or attempt | 72688 | 6386 | 1.05 (1.04,1.07) | 1.6E-15 | 1.04 (1.02,1.05) | 7.8E-08 | 1.03 (1.02,1.05) | 6.7E-06 |
| mental disorders | 297.1 | Suicidal ideation | 74668 | 5503 | 1.05 (1.04,1.07) | 2.3E-14 | 1.04 (1.02,1.05) | 2.2E-07 | 1.03 (1.02,1.05) | 2.0E-05 |
| mental disorders | 300 | Anxiety disorders | 36301 | 40111 | 1.04 (1.03,1.05) | 4.5E-26 | 1.02 (1.01,1.03) | 2.8E-04 | 1.03 (1.02,1.04) | 1.0E-12 |
| mental disorders | 300.1 | Anxiety disorder | 51938 | 21915 | 1.04 (1.03,1.05) | 9.6E-23 | 1.02 (1.01,1.03) | 3.0E-07 | 1.03 (1.02,1.04) | 8.5E-13 |
| mental disorders | 300.11 | Generalized anxiety disorder | 74900 | 4671 | 1.03 (1.02,1.05) | 4.8E-06 | 1.02 (1,1.03) | 1.3E-02 | 1.02 (1.01,1.04) | 1.5E-03 |
| mental disorders | 300.9 | Posttraumatic stress disorder | 52155 | 26603 | 1.02 (1.02,1.03) | 7.4E-10 | 1 (1,1.01) | 3.2E-01 | 1.01 (1,1.02) | 5.3E-03 |
| mental disorders | 301 | Personality disorders | 75463 | 4935 | 1.07 (1.06,1.09) | 1.3E-20 | 1.05 (1.04,1.07) | 3.1E-12 | 1.05 (1.03,1.06) | 2.3E-09 |
| mental disorders | 301.2 | Antisocial/borderline personality dis. | 79773 | 2083 | 1.09 (1.07,1.11) | 5.2E-15 | 1.07 (1.05,1.1) | 2.6E-10 | 1.06 (1.04,1.09) | 8.0E-08 |
| mental disorders | 304 | Adjustment reaction | 55082 | 18851 | 1.02 (1.01,1.03) | 3.7E-08 | 1.01 (1,1.02) | 1.1E-01 | 1.02 (1.01,1.03) | 1.2E-04 |
| mental disorders | 306 | Other mental disorder | 62819 | 9537 | 1.04 (1.03,1.05) | 5.9E-12 | 1.02 (1.01,1.03) | 4.0E-05 | 1.02 (1.01,1.04) | 3.2E-05 |
| mental disorders | 316 | Substance addiction and disorders | 55813 | 22948 | 1.03 (1.02,1.04) | 1.2E-14 | 1.02 (1.01,1.03) | 6.1E-05 | 1.02 (1.01,1.03) | 2.6E-05 |
| mental disorders | 317.1 | Alcoholism | 58054 | 19278 | 1.03 (1.02,1.04) | 1.8E-10 | 1.01 (1,1.02) | 2.2E-03 | 1.01 (1.01,1.02) | 1.0E-03 |
| mental disorders | 318 | Tobacco use disorder | 43079 | 33056 | 1.03 (1.02,1.04) | 3.4E-14 | 1.02 (1.01,1.03) | 4.7E-07 | 1.02 (1.01,1.03) | 1.2E-07 |
| musculoskeletal | 735.2 | Acquired toe deformities | 69294 | 8813 | 1.03 (1.01,1.04) | 4.4E-06 | 1.02 (1.01,1.03) | 6.8E-04 | 1.02 (1.01,1.03) | 1.7E-04 |
| musculoskeletal | 741.3 | Difficulty in walking | 71434 | 6164 | 1.03 (1.02,1.05) | 1.7E-06 | 1.02 (1.01,1.04) | 4.0E-04 | 1.03 (1.01,1.04) | 1.6E-04 |
| neurological | 327 | Sleep disorders | 47798 | 23542 | 1.02 (1.01,1.03) | 1.7E-06 | 1.01 (1,1.01) | 1.9E-01 | 1.01 (1,1.02) | 7.4E-03 |
| neurological | 327.4 | Insomnia | 58442 | 15340 | 1.02 (1.01,1.03) | 1.8E-06 | 1.01 (1,1.02) | 8.0E-02 | 1.01 (1,1.02) | 5.3E-03 |
| neurological | 338 | Pain | 56011 | 15018 | 1.03 (1.02,1.04) | 5.1E-09 | 1.02 (1.01,1.03) | 7.2E-04 | 1.02 (1.01,1.03) | 6.1E-05 |
| neurological | 338.2 | Chronic pain | 62731 | 11206 | 1.03 (1.01,1.04) | 8.0E-07 | 1.01 (1,1.02) | 7.0E-03 | 1.02 (1.01,1.03) | 9.5E-04 |
| neurological | 339 | Other headache syndromes | 54393 | 17396 | 1.03 (1.02,1.04) | 1.2E-09 | 1.02 (1.01,1.03) | 4.0E-04 | 1.02 (1.01,1.03) | 1.4E-05 |
| respiratory | 465 | Acute upper respiratory infections | 46098 | 21217 | 1.04 (1.03,1.04) | 2.9E-17 | 1.03 (1.02,1.03) | 1.3E-09 | 1.03 (1.02,1.04) | 3.2E-11 |
| respiratory | 496 | Chronic airway obstruction | 61946 | 14621 | 1.03 (1.02,1.04) | 9.8E-11 | 1.02 (1.01,1.03) | 1.8E-06 | 1.02 (1.01,1.03) | 1.5E-06 |

| | | | | | | | | | | |
|---|---|---|---|---|---|---|---|---|---|---|
| respiratory | 496.2 | Chronic bronchitis | 75733 | 3835 | 1.04 (1.02,1.06) | 3.3E-06 | 1.03 (1.01,1.05) | 1.8E-04 | 1.03 (1.01,1.05) | 1.6E-04 |
| respiratory | 497 | Bronchitis | 69829 | 4681 | 1.04 (1.03,1.06) | 4.8E-09 | 1.04 (1.02,1.05) | 3.9E-06 | 1.04 (1.02,1.05) | 2.0E-06 |
| respiratory | 512 | Other respiratory symptoms | 34852 | 33303 | 1.03 (1.02,1.04) | 1.6E-14 | 1.02 (1.01,1.03) | 1.2E-07 | 1.02 (1.02,1.03) | 2.3E-09 |
| respiratory | 512.7 | Shortness of breath | 56849 | 15224 | 1.03 (1.02,1.03) | 3.6E-08 | 1.02 (1.01,1.03) | 1.7E-04 | 1.02 (1.01,1.03) | 3.7E-05 |
| respiratory | 512.8 | Cough | 54075 | 14679 | 1.03 (1.02,1.04) | 2.3E-10 | 1.02 (1.01,1.03) | 5.1E-06 | 1.02 (1.01,1.03) | 3.6E-07 |
| sense organs | 367.1 | Myopia | 58235 | 13637 | 1.02 (1.01,1.03) | 3.1E-06 | 1.02 (1.01,1.03) | 4.1E-04 | 1.02 (1.01,1.03) | 3.1E-04 |
| sense organs | 386.9 | Dizziness and giddiness | 62691 | 10111 | 1.03 (1.02,1.04) | 4.5E-07 | 1.02 (1.01,1.03) | 6.9E-04 | 1.02 (1.01,1.03) | 7.4E-05 |
| symptoms | 760 | Back pain | 26406 | 47752 | 1.03 (1.02,1.04) | 1.5E-13 | 1.02 (1.01,1.03) | 2.0E-06 | 1.02 (1.02,1.03) | 3.1E-09 |
| symptoms | 770 | Myalgia and myositis unspecified | 70525 | 5917 | 1.04 (1.02,1.05) | 2.1E-07 | 1.02 (1.01,1.04) | 3.5E-04 | 1.03 (1.01,1.04) | 4.8E-05 |
| symptoms | 785 | Abdominal pain | 49712 | 20812 | 1.03 (1.02,1.04) | 9.9E-12 | 1.02 (1.01,1.03) | 4.2E-06 | 1.02 (1.01,1.03) | 4.0E-07 |
| symptoms | 789 | Nausea and vomiting | 67505 | 6884 | 1.03 (1.02,1.04) | 1.9E-06 | 1.02 (1.01,1.03) | 1.6E-03 | 1.02 (1.01,1.03) | 1.5E-03 |
| symptoms | 798 | Malaise and fatigue | 61002 | 11331 | 1.03 (1.02,1.04) | 1.3E-08 | 1.02 (1.01,1.03) | 8.4E-05 | 1.02 (1.01,1.03) | 1.5E-05 |

**Supplemental Figure 1. Comparative performance of SCZ PRS based on varying GWAS training sets.**

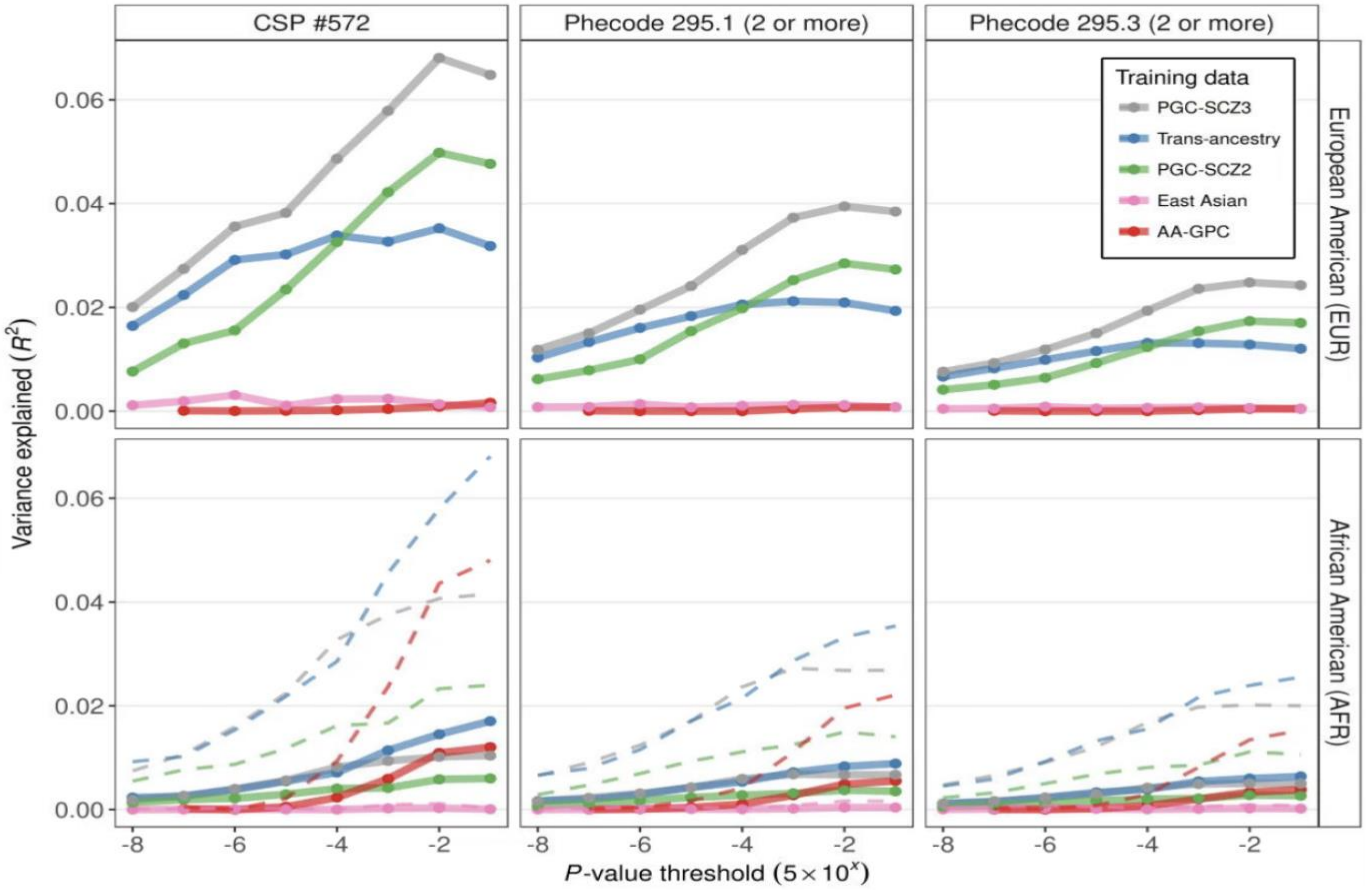

**Supplemental Figure 2. Odds ratios for top deciles of SCZ PRS in MVP.** Odds ratios for top neuropsychiatric PRS deciles compared to the bottom 90%.

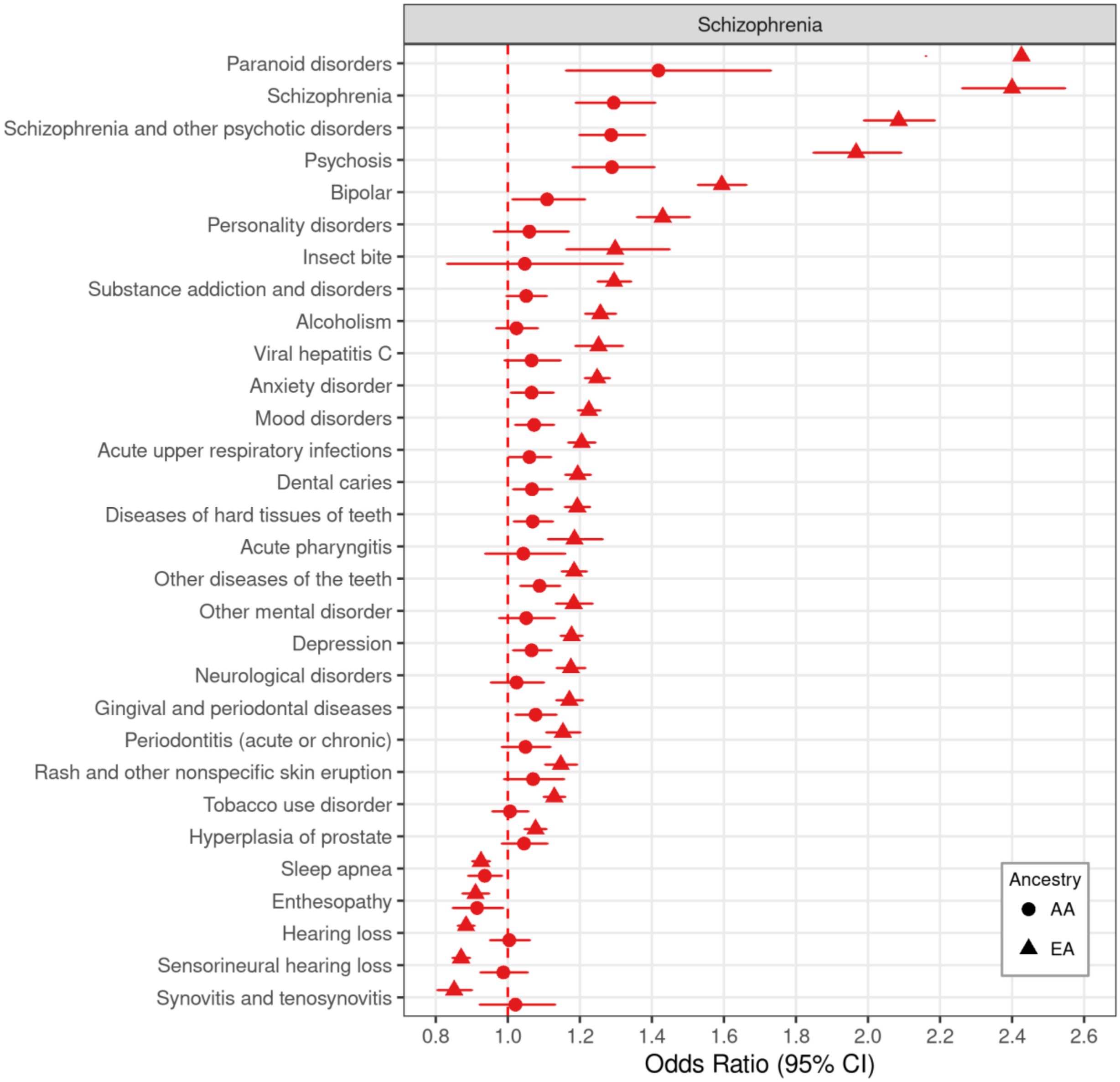

**Supplemental Figure 3. Odds ratios for top deciles of BIP PRS in MVP.** Odds ratios for top neuropsychiatric PRS deciles compared to the bottom 90%.

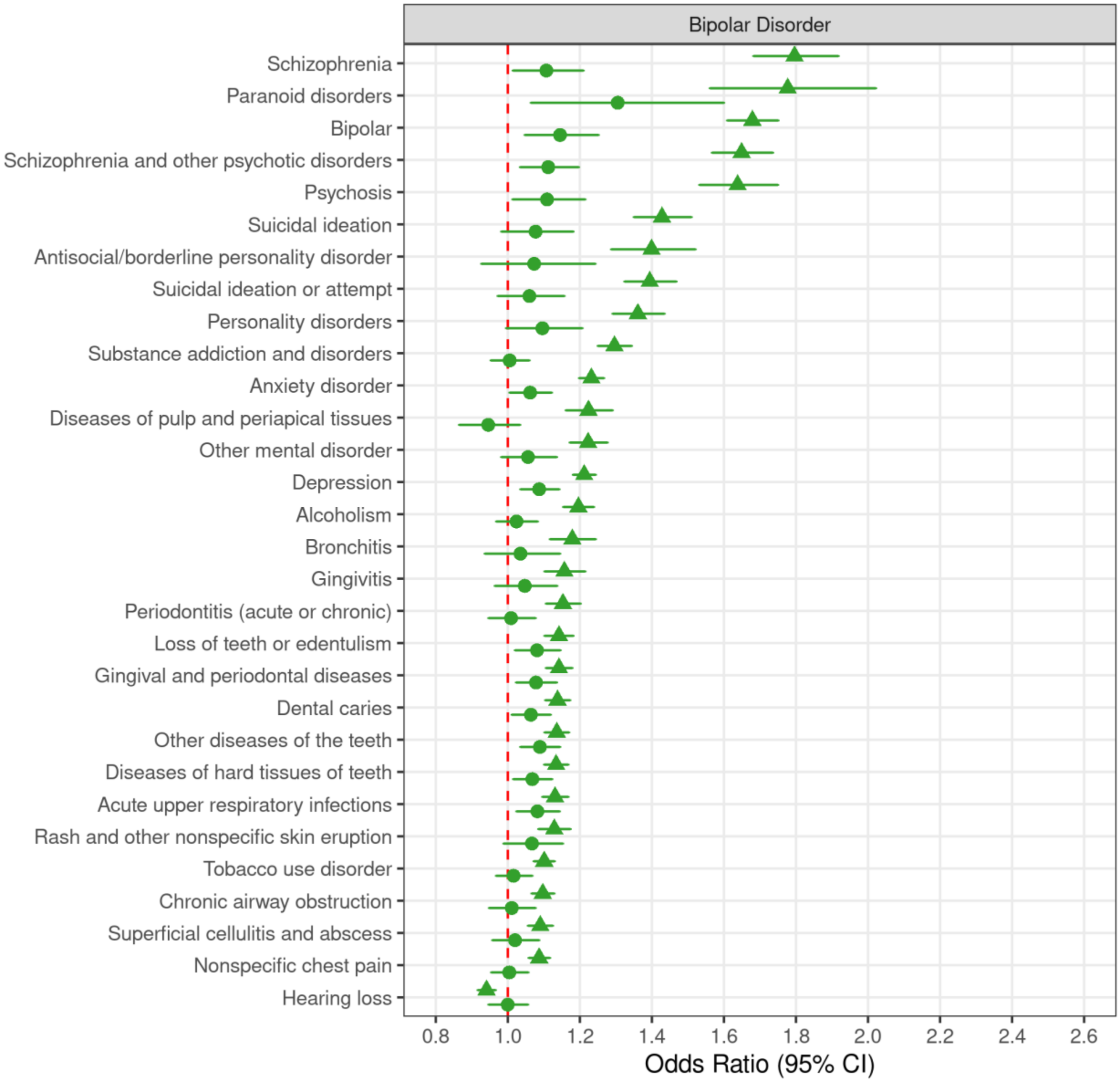

**Supplemental Figure 4. Odds ratios for top deciles of MDD PRS in MVP.** Odds ratios for top neuropsychiatric PRS deciles compared to the bottom 90%.

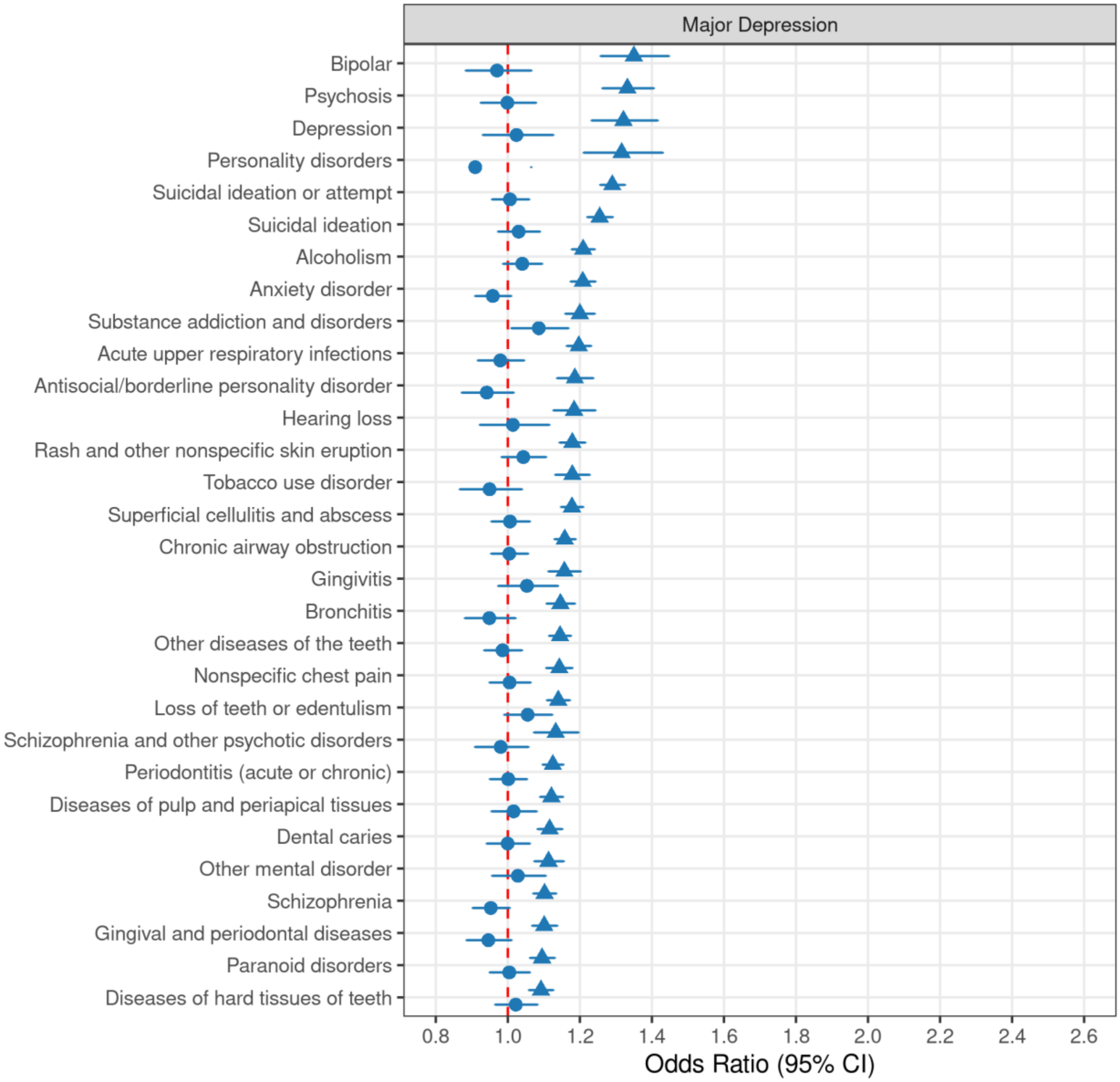

**Supplemental Figure 5. Relative enrichments of SCZ and BIP PheWAS results across disease categories.** For each disease category, ranked *p*-values for SCZ are plotted against ranked p-values for BIP. Displayed results are based on EA participants.

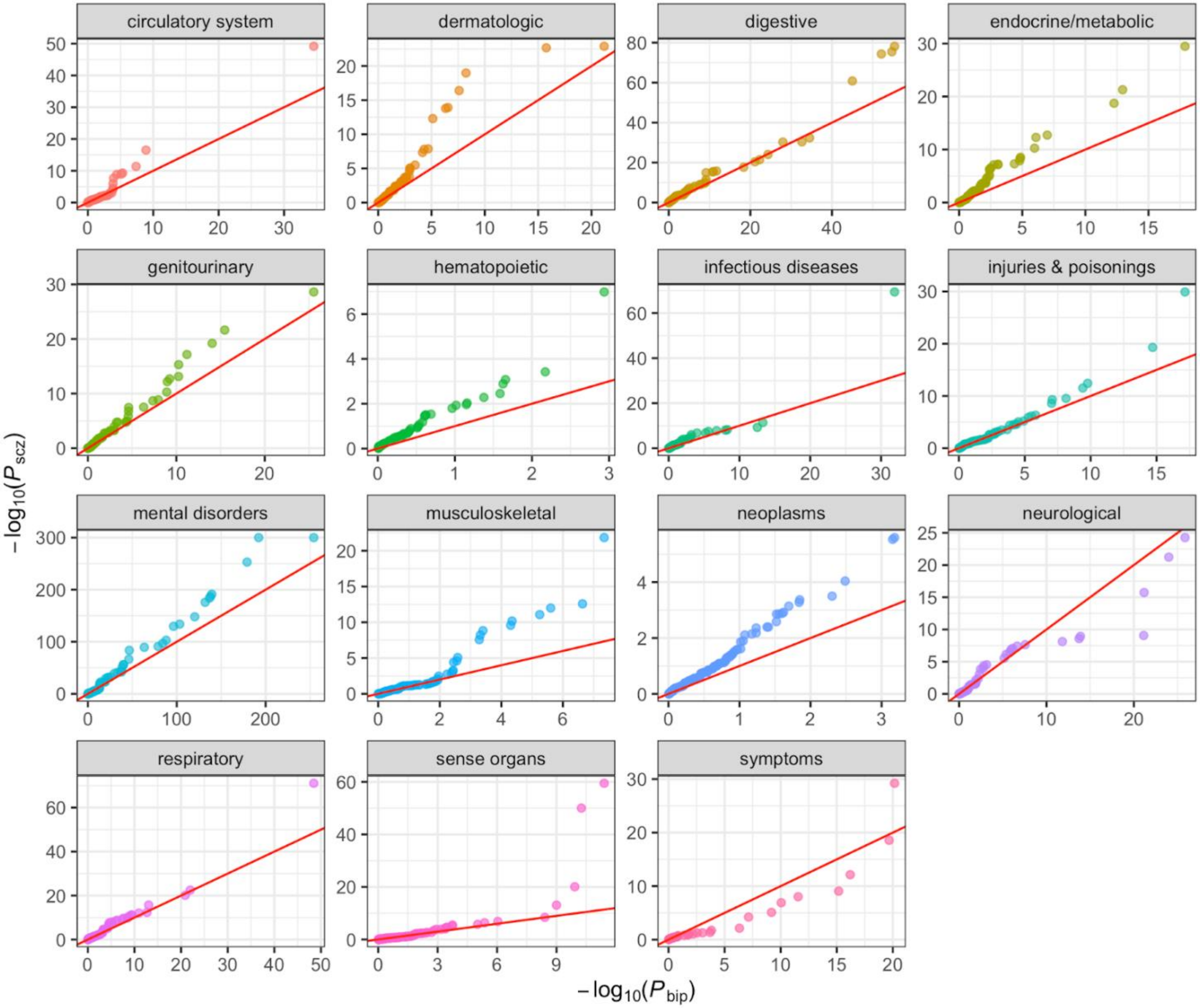

**Supplemental Figure 6. Relative enrichments of SCZ and DEP PheWAS results across disease categories.** For each disease category, ranked *p*-values for SCZ are plotted against ranked p-values for DEP. Displayed results are based on EA participants.

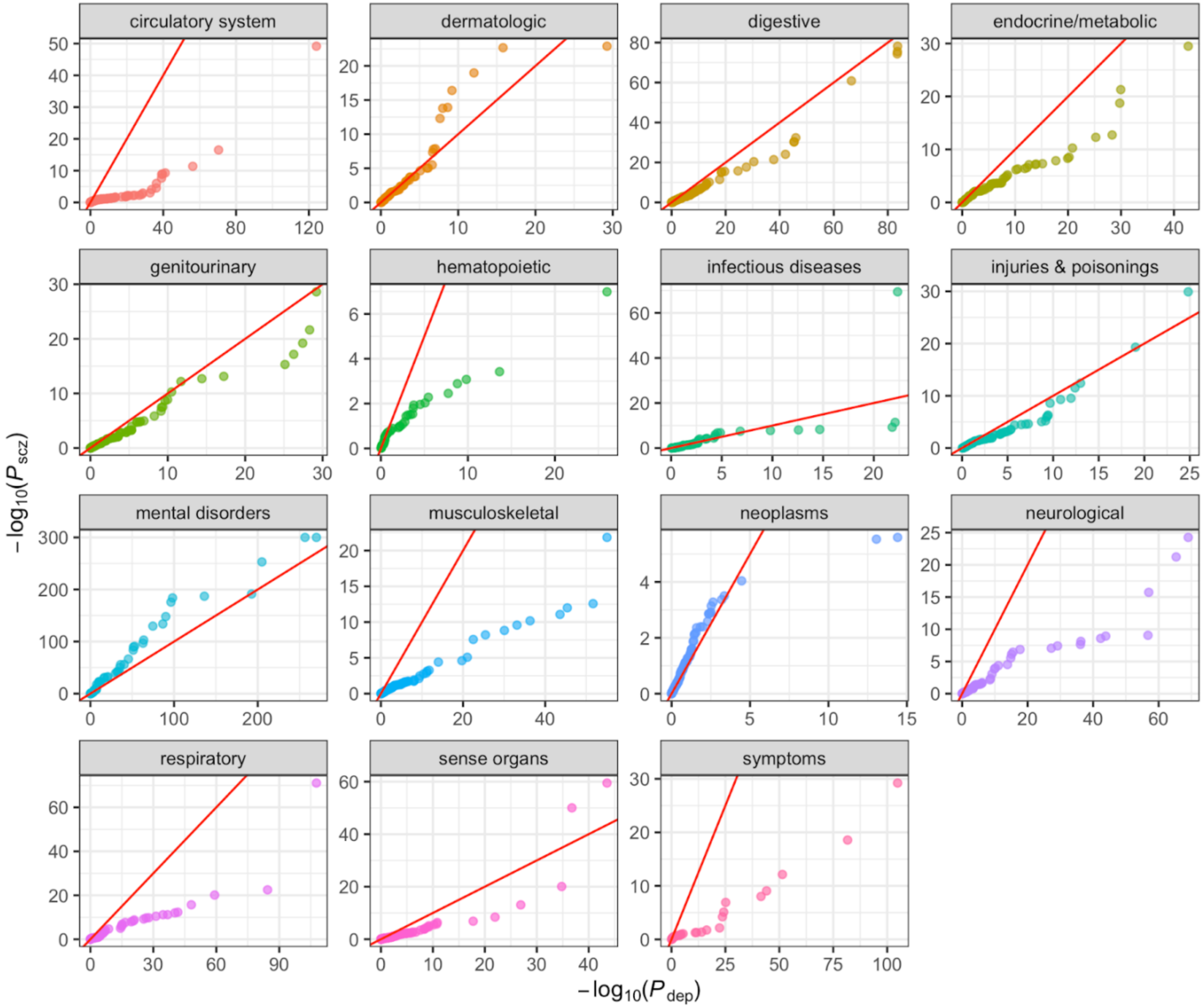

**Supplemental Figure 7. Relative enrichments of BIP and DEP PheWAS results across disease categories.** For each disease category, ranked *p*-values for BIP are plotted against ranked p-values for DEP. Displayed results are based on EA participants.

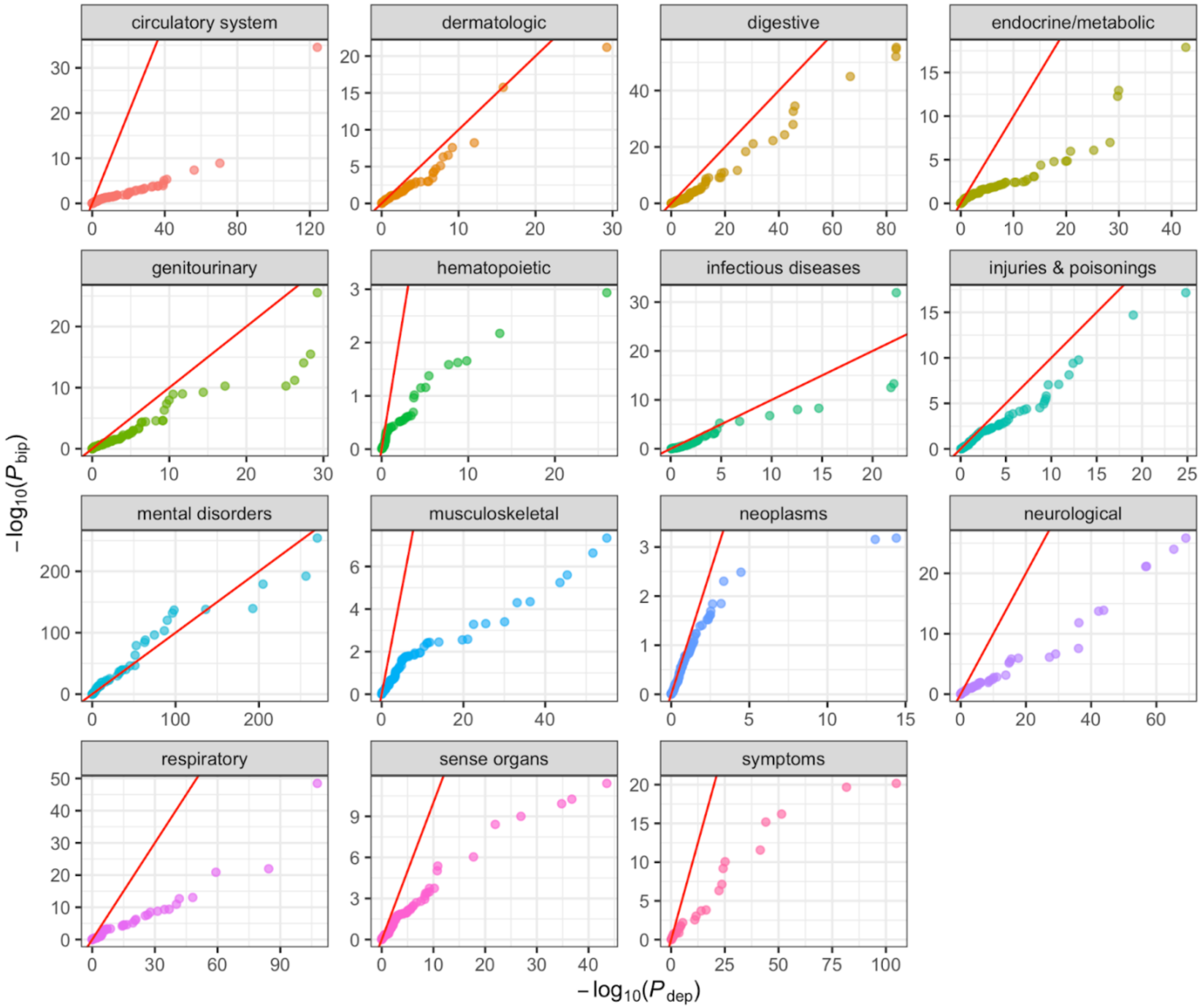

### SUPPLEMENTARY ACKNOWLEDGEMENTS

The CSP #572 study team

**Planning Committee:** M. Aslan, M. Antonelli, M. de Asis, M. S. Bauer, M. Brophy, J. Concato, F. Cunningham, R. Freedman, M. Gaziano, T. Gleason, P. D. Harvey, G. Huang, J. Kelsoe, T. Kosten, T. Lehner, J. B. Lohr, S. R. Marder, P. Miller, T.J. O'Leary, T. Patterson, P. Peduzzi, R. Przygodzki, L. Siever, P. Sklar, S. Strakowski, H. Zhao.

**Executive Committee:** M. Brophy, J. Concato, A.H. Fanous, W. Farwell, M. Gaziano, P.D. Harvey, T. Kosten, A. Malhotra, S. Mane, P. Palacios, P. Sklar, L. Siever, H. Zhao, T.B. Bigdeli.

**Study Chairs' Offices:** VA Healthcare System, Bronx, NY, included L. Siever (Study Co- Chair), M. Corsey, L. Zaluda. VA Healthcare System, Miami, FL, included P.D. Harvey (Study Co- Chair), J. Johnson/ M Sueiro.

**CSP Epidemiology Centers:** VA Clinical Epidemiology Research Center (CERC), VA Connecticut Healthcare System, West Haven, CT, included J. Concato (Director, Methodological Co-Principal Proponent), M. Aslan, D. Cavaliere, V. Jeanpaul, A. Maffucci, L. Mancini; the Massachusetts Veterans Epidemiology Research and Information Center (MAVERIC), VA Boston Healthcare System, Jamaica Plain, MA, included M. Gaziano (Director, Methodological Co-Principal Proponent), J. Deen, G. Muldoon, S. Whitbourne.

**Study Sites:** *Albuquerque*: J. Canive, L. Adamson, L. Calais, G. Fuldauer, R. Kushner, G. Toney, M. Lackey, A. Mank, N. Mahdavi, G. Villarreal. *Atlanta*: E. C. Muly, F. Amin, M. Dent, J. Wold. *Baltimore*: B. Fischer, A. Elliott, C. Felix, G. Gill. *Birmingham*: P. E. Parker, C. Logan, J. McAlpine. *Brockton*: L.E. DeLisi, S. G Reece. *Charleston*: M.B. Hammer, D. Agbor-Tabie, W. Goodson. *Cincinnati*: M. Aslam, M. Grainger, Neil Richtand, Alexander Rybalsky. *Houston*: R. Al Jurdi, E. Boeckman, T. Natividad, D. Smith, M. Stewart, S. Torres, Z. Zhao. *Indianapolis*: A. Mayeda, A. Green, J. Hofstetter, S. Ngombu, M. K. Scott, A. Strasburger, J. Sumner. *Little Rock*: G. Paschall, J. Mucciarelli, R. Owen, S. Theus, D. Tompkins. *Long Beach*: S.G. Potkin, C. Reist, M. Novin, S. Khalaghizadeh. *Miami*: R. Douyon, J. Johnson, N. Kumar, B. Martinez. *Minneapolis*: S.R. Sponheim, T.L. Bender, H.L. Lucas, A.M. Lyon, M.P. Marggraf, L.H. Sorensen, C.R. Surerus. *Montrose*: C. Sison, J. Amato, D.R. Johnson, N. Pagan-Howard. *New York Harbor*: L.A. Adler, S. Alperin, T. Leon. *Northampton*: K.M. Mattocks, N. Araeva, J.C. Sullivan. *Palo Alto*: T. Suppes, K. Bratcher, L. Drag, E.G. Fischer, L. Fujitani, S. Gill, D. Grimm, J. Hoblyn, T. Nguyen, E. Nikolaev, L. Shere, R. Relova, A. Vicencio, M. Yip. *Philadelphia*: I. Hurford, S. Acheampong, G. Carfagno. *Pittsburgh*: G.L. Haas, C. Appelt, E. Brown, B. Chakraborty, E. Kelly, G. Klima, S. Steinhauer. *Salisbury*: R.A. Hurley, R. Belle, D. Eknoyan, K. Johnson, J. Lamotte. *San Diego*: E. Granholm, K. Bradshaw, J. Holden, R. H. Jones, T. Le, I.G. Molina, M. Peyton, I. Ruiz, L. Sally. *Tacoma*: A. Tapp, S. Devroy, V. Jain, N. Kilzieh, L. Maus, K. Miller, H. Pope, A. Wood. *Temple*: E. Meyer, P. Givens, P. B. Hicks, S. Justice, K. McNair, J.L. Pena, D.F. Tharp. *Tuscaloosa*: L. Davis, M. Ban, L. Cheatum, P. Darr, W. Grayson, J. Munford, D. Smith, B. Whitfield, E. Wilson. *Washington DC*: A.H. Fanous, S.E. Melnikoff, B.L Schwartz, M.A. Tureson. *West Haven*: D. D'Souza, K. Forselius, M. Ranganathan, L. Rispoli.

**Albuquerque, NM, CSP Coordinating Center (for monitoring)**: M. Sather (Director), C. Colling, C. Haakenson, D. Krueger.

**VA Office of Research and Development**: T. O'Leary (Chief Research and Development Officer Emeritus), G. Huang (Director, Cooperative Studies Program), T. Gleason (Director, Clinical Science Research and Development Service), R. Przygodzki (Associate Director for Genomic Medicine, and Acting Director of Biomedical Laboratory Research and Development Service), S. Muralidhar (Senior Scientific Program Manager Genomic Medicine Program, Biomedical and Clinical R&D Services).

**Million Veteran Program: Consortium Acknowledgement for Manuscripts**

**MVP Executive Committee**

- Co-Chair: J. Michael Gaziano, M.D., M.P.H.
- Co-Chair: Rachel Ramoni, D.M.D., Sc.D.
- Jim Breeling, M.D. (ex-officio)
- Kyong-Mi Chang, M.D.
- Grant Huang, Ph.D.
- Sumitra Muralidhar, Ph.D.
- Christopher J. O'Donnell, M.D., M.P.H.
- Philip S. Tsao, Ph.D.

**MVP Program Office**

- Sumitra Muralidhar, Ph.D.
- Jennifer Moser, Ph.D.

**MVP Recruitment/Enrollment**

- Recruitment/Enrollment Director/Deputy Director, Boston
  - Stacey B. Whitbourne, Ph.D.; Jessica V. Brewer, M.P.H.
- MVP Coordinating Centers
  - o Clinical Epidemiology Research Center (CERC), West Haven – John Concato, M.D., M.P.H.
  - o Cooperative Studies Program Clinical Research Pharmacy Coordinating Center, Albuquerque - Stuart Warren, J.D., Pharm D.; Dean P. Argyres, M.S.
  - o Genomics Coordinating Center, Palo Alto – Philip S. Tsao, Ph.D.
  - o Massachusetts Veterans Epidemiology Research Information Center (MAVERIC), Boston - J. Michael Gaziano, M.D., M.P.H.
  - o MVP Information Center, Canandaigua – Brady Stephens, M.S.
- Core Biorepository, Boston – Mary T. Brophy M.D., M.P.H.; Donald E. Humphries, Ph.D.
- MVP Informatics, Boston – Nhan Do, M.D.; Shahpoor Shayan
- Data Operations/Analytics, Boston – Xuan-Mai T. Nguyen, Ph.D.

**MVP Science**

- Genomics - Christopher J. O'Donnell, M.D., M.P.H.; Saiju Pyarajan Ph.D.; Philip S. Tsao, Ph.D.
- Phenomics - Kelly Cho, M.P.H, Ph.D.
- Data and Computational Sciences – Saiju Pyarajan, Ph.D.
- Statistical Genetics – Elizabeth Hauser, Ph.D.; Yan Sun, Ph.D.; Hongyu Zhao, Ph.D.

**MVP Local Site Investigators**

- Atlanta VA Medical Center (Peter Wilson) –
- Bay Pines VA Healthcare System (Rachel McArdle)
- Birmingham VA Medical Center (Louis Dellitalia)
- Cincinnati VA Medical Center (John Harley)
- Clement J. Zablocki VA Medical Center (Jeffrey Whittle)
- Durham VA Medical Center (Jean Beckham)
- Edith Nourse Rogers Memorial Veterans Hospital (John Wells)
- Edward Hines, Jr. VA Medical Center (Salvador Gutierrez)
- Fayetteville VA Medical Center (Gretchen Gibson)
- VA Health Care Upstate New York (Laurence Kaminsky)
- New Mexico VA Health Care System (Gerardo Villareal)
- VA Boston Healthcare System (Scott Kinlay)
- VA Western New York Healthcare System (Junzhe Xu)

- Ralph H. Johnson VA Medical Center (Mark Hamner)
- Wm. Jennings Bryan Dorn VA Medical Center (Kathlyn Sue Haddock)
- VA North Texas Health Care System (Sujata Bhushan)
- Hampton VA Medical Center (Pran Iruvanti)
- Hunter Holmes McGuire VA Medical Center (Michael Godschalk)
- Iowa City VA Health Care System (Zuhair Ballas)
- Jack C. Montgomery VA Medical Center (Malcolm Buford)
- James A. Haley Veterans' Hospital (Stephen Mastorides)
- Louisville VA Medical Center (Jon Klein)
- Manchester VA Medical Center (Nora Ratcliffe)
- Miami VA Health Care System (Hermes Florez)
- Michael E. DeBakey VA Medical Center (Alan Swann)
- Minneapolis VA Health Care System (Maureen Murdoch)
- N. FL/S. GA Veterans Health System (Peruvemba Sriram)
- Northport VA Medical Center (Shing Shing Yeh)
- Overton Brooks VA Medical Center (Ronald Washburn)
- Philadelphia VA Medical Center (Darshana Jhala)
- Phoenix VA Health Care System (Samuel Aguayo)
- Portland VA Medical Center (David Cohen)
- Providence VA Medical Center (Satish Sharma)
- Richard Roudebush VA Medical Center (John Callaghan)
- Salem VA Medical Center (Kris Ann Oursler)
- San Francisco VA Health Care System (Mary Whooley)
- South Texas Veterans Health Care System (Sunil Ahuja)
- Southeast Louisiana Veterans Health Care System (Amparo Gutierrez)
- Southern Arizona VA Health Care System (Ronald Schifman)
- Sioux Falls VA Health Care System (Jennifer Greco)
- St. Louis VA Health Care System (Michael Rauchman)
- Syracuse VA Medical Center (Richard Servatius)
- VA Eastern Kansas Health Care System (Mary Oehlert)
- VA Greater Los Angeles Health Care System (Agnes Wallbom)
- VA Loma Linda Healthcare System (Ronald Fernando)
- VA Long Beach Healthcare System (Timothy Morgan)
- VA Maine Healthcare System (Todd Stapley)
- VA New York Harbor Healthcare System (Scott Sherman)
- VA Pacific Islands Health Care System (Gwenevere Anderson)
- VA Palo Alto Health Care System (Philip Tsao)
- VA Pittsburgh Health Care System (Elif Sonel)
- VA Puget Sound Health Care System (Edward Boyko)
- VA Salt Lake City Health Care System (Laurence Meyer)
- VA San Diego Healthcare System (Samir Gupta)
- VA Southern Nevada Healthcare System (Joseph Fayad)
- VA Tennessee Valley Healthcare System (Adriana Hung)
- Washington DC VA Medical Center (Jack Lichy)
- W.G. (Bill) Hefner VA Medical Center (Robin Hurley)
- White River Junction VA Medical Center (Brooks Robey)
- William S. Middleton Memorial Veterans Hospital (Robert Striker)
